# CHANGES IN LIFE EXPECTANCY BETWEEN 2019 AND 2021 IN THE UNITED STATES AND 21 PEER COUNTRIES

**DOI:** 10.1101/2022.04.05.22273393

**Authors:** Ryan K. Masters, Laudan Y. Aron, Steven H. Woolf

## Abstract

**BACKGROUND:** Prior studies reported large decreases in US life expectancy during 2020 as a result of the COVID-19 pandemic, disproportionately affecting Hispanic and Black populations and vastly exceeding the average change in life expectancy in other high-income countries. Life expectancy estimates for 2021 have not been reported. This study estimated changes in life expectancy during 2019-2021 in the US population, in US racial/ethnic groups, and in 21 peer countries. The study compared outcomes across five US racial/ethnic groups and is the first to estimate changes in life expectancy during the pandemic in non-Hispanic American Indian/Alaska Native and Asian populations.

**METHODS:** US and peer country death data for 2019-2021 were obtained from the National Center for Health Statistics, the Human Mortality Database, and overseas statistical agencies. The 21 peer countries included Australia, Austria, Belgium, Canada, Denmark, England and Wales, Finland, France, Germany, Israel, Italy, Netherlands, New Zealand, Northern Ireland, Norway, Portugal, Scotland, South Korea, Spain, Sweden, and Switzerland. Life expectancy was calculated for 2019 and 2020 and estimated for 2021 using a previously validated modeling method.

**RESULTS:** US life expectancy decreased from 78.85 years in 2019 to 76.98 years in 2020 and 76.44 years in 2021, a net loss of 2.41 years. In contrast, peer countries averaged a smaller decrease in life expectancy between 2019 and 2020 (0.55 years) and a 0.26-year *increase* between 2020 and 2021, widening the gap in life expectancy between the United States and peer countries to more than five years. The decrease in US life expectancy was highly racialized: whereas the largest decreases in 2020 occurred among non-Hispanic (NH) American Indian/Alaska Native, Hispanic, NH Black, and NH Asian populations, in 2021 the largest decreases occurred in the NH White population.

**DISCUSSION:** The US mortality experience during 2020 and 2021 was more severe than in peer countries, deepening a US disadvantage in health and survival that has been building for decades. Over the two-year period between 2019 and 2021, US NH American Indian/Alaska Native, Hispanic, and NH Black populations experienced the largest losses in life expectancy, reflecting the ongoing legacy of systemic racism as well as inadequacies in the US handling of the pandemic. The crossover in racialized outcomes between 2020 and 2021, in which the NH White population experienced the largest decreases, likely has multiple explanations.

## INTRODUCTION

In 2020, the United States (US) experienced a much larger decline in life expectancy than did other high-income countries, with disproportionately large losses in its Hispanic and non-Hispanic (NH) Black populations.^1^ Although the introduction and availability of effective vaccines were expected to curb US mortality rates in 2021, slow vaccine uptake and the spread of the Delta variant produced large surges in mortality. This study used official mortality data for 2019 and 2020 and provisional mortality data for 2021 to estimate changes in life expectancy in the US population, in US racial/ethnic groups, and in 21 peer countries. The study compared outcomes across five US racial/ethnic groups and is the first to include separate estimates for NH American Indian/Alaska Native (AI/AN) and NH Asian populations.

## METHODS

Methods used previously to estimate life expectancy for 2020 based on provisional death counts^1^ were replicated for 2021. Official and provisional mortality data for US populations were obtained from the National Center for Health Statistics.^2,3^ Data for 21 other high-income advanced democratic countries were obtained from the Human Mortality Database (HMD), direct sources, and the HMD Short-term Mortality Fluctuation series.^4^ Data for all countries were stratified by sex, and US data were further stratified by race and ethnicity for Hispanic and NH Asian, NH AI/AN, NH Black, and NH White populations (see supplementary material for details). A credible range (CR) for life expectancy in 2021 was calculated by simulating life tables from estimated age-specific mortality rates and allowing for 10% random error.

## RESULTS

US life expectancy decreased by 0.54 (CR = 0.36-0.71) years between 2020 and 2021, adding to an historic 1.87-year reduction in 2020 (**Figure 1**). Changes in US life expectancy during 2020 and 2021 were highly racialized but followed a distinctly different pattern. In 2020, the largest decreases in life expectancy occurred in NH AI/AN (4.48 [CR = 4.27-4.69] years), Hispanic (3.72 [CR = 3.55-3.90] years), NH Black (3.20 [CR = 3.01-3.39] years), and NH Asian (1.83 [CR = 1.67-1.99] years) populations, and the smallest decrease occurred in the NH White population (1.38 [CR = 1.20-1.54] years).

**Figure 1.**
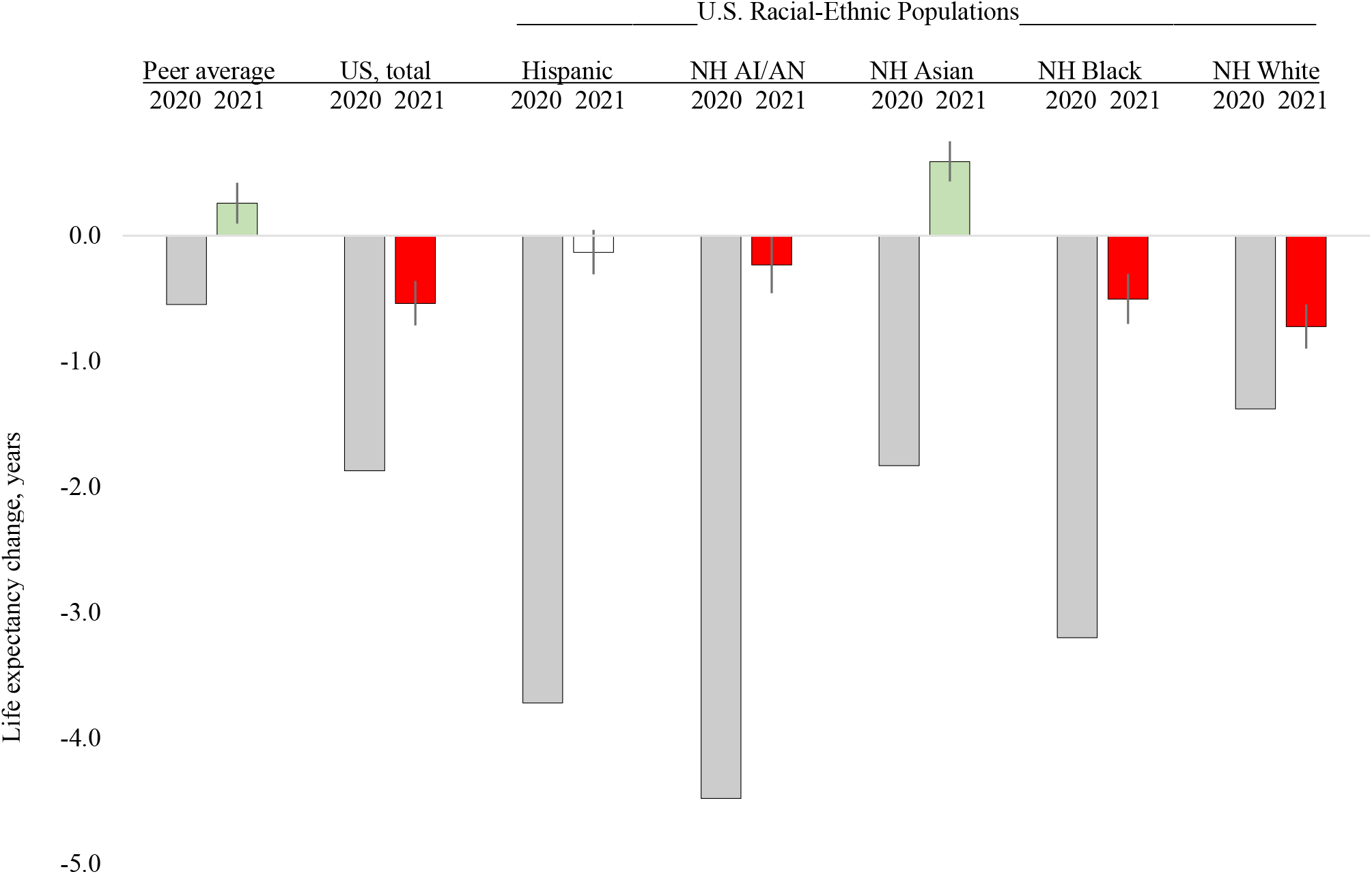
Changes in US life expectancy and average changes in life expectancy among 21 peer countries, 2019-2020 and 2020-2021. AI/AN = American Indian/Alaska Native; NH = Non-Hispanic. Vertical bars for 2021 estimates depict the credible range based on 10% random error in mortality risks. Separate figures for females and males are in the supplement.

In 2021, by contrast, decreases in life expectancy were larger in the NH White population (0.72 [CR = 0.55-0.90 years] years) than in NH Black (0.50 [CR = 0.30-0.70] years) and NH AI/AN (0.23 [CR = 0.01-0.46] years) populations. Life expectancy did not change significantly in Hispanic populations (CR = -0.31 to 0.05 years) and increased by 0.59 (CR = 0.43-0.75) years in the NH Asian population. (**Table 1**).

**Table 1.** Life expectancy in United States and 21 peer countries: 2019, 2020, and 2021.

|  | Life expectancy (years), and confidence range (CR) for 2021 estimates |  |  |  |  |  |  |  |  |
| --- | --- | --- | --- | --- | --- | --- | --- | --- | --- |
|  | Total population |  |  | Female population |  |  | Male population |  |  |
|  | 2019 | 2020 | 2021 (CR) | 2019 | 2020 | 2021 (CR) | 2019 | 2020 | 2021 (CR) |
| **United States** | **78.86** | **76.98** | **76.44 (76.27-76.62)** | **81.39** | **79.88** | **79.49 (79.32-79.66)** | **76.31** | **74.18** | **73.51 (73.33-73.69)** |
| Hispanic | 81.85 | 78.13 | 78.00 (77.82-78.18) | 84.43 | 81.60 | 81.46 (81.30-81.63) | 79.07 | 74.73 | 74.60 (74.41-74.78) |
| NH AI/AN | 71.75 | 67.27 | 67.04 (66.81-67.26) | 74.95 | 70.92 | 70.43 (70.22-70.65) | 68.64 | 63.90 | 63.85 (63.62-64.09) |
| NH Asian | 85.56 | 83.73 | 84.32 (84.16-84.48) | 87.43 | 86.01 | 86.52 (86.37-86.68) | 83.46 | 81.25 | 81.90 (81.74-82.06) |
| NH Black | 74.78 | 71.58 | 71.08 (70.88-71.28) | 78.1- | 75.39 | 74.99 (74.80-75.18) | 71.32 | 67.79 | 67.20 (67.00-67.41) |
| NH White | 78.78 | 77.40 | 76.68 (76.50-76.85) | 81.26 | 80.10 | 79.59 (79.42-79.76) | 76.33 | 74.80 | 73.91 (73.74-74.09) |
| **Peer Average** | **82.22** | **81.67** | **81.93 (81.77-82.09)** | **84.37** | **83.94** | **84.20 (84.04-84.36)** | **80.02** | **79.40** | **79.69 (79.53-79.86)** |
| Australia | 83.32 | 83.57 | 83.55 (83.39-83.71) | 85.37 | 85.62 | 85.49 (85.34-85.65) | 81.28 | 81.51 | 81.60 (81.44-81.77) |
| Austria | 81.91 | 81.04 | 81.28 (81.12-81.44) | 84.20 | 83.47 | 83.78 (83.63-83.94) | 79.54 | 78.57 | 78.74 (78.58-78.91) |
| Belgium | 81.84 | 80.79 | 81.77 (81.61-81.94) | 84.00 | 83.06 | 84.17 (84.01-84.33) | 79.59 | 78.53 | 79.36 (79.19-79.52) |
| Canada | 82.37 | 81.39 | 81.65 (81.48-81.82) | 84.43 | 83.67 | 84.04 (83.88-84.20) | 80.28 | 79.12 | 79.26 (79.10-79.43) |
| Denmark | 81.43 | 81.55 | 81.58 (81.42-81.74) | 83.43 | 83.51 | 83.45 (83.29-83.61) | 79.44 | 79.58 | 79.71 (79.55-79.88) |
| England/Wales | 81.71 | 80.43 | 80.78 (80.62-80.95) | 83.53 | 82.43 | 82.79 (82.63-82.95) | 79.85 | 78.43 | 78.78 (78.61-78.95) |
| Finland | 81.91 | 81.87 | 81.97 (81.81-82.13) | 84.53 | 84.66 | 84.91 (84.75-85.08) | 79.18 | 79.08 | 79.48 (79.31-79.65) |
| France | 82.75 | 82.06 | 82.61 (82.45-82.77) | 85.62 | 85.04 | 85.56 (85.41-85.72) | 79.75 | 79.01 | 79.55 (79.38-79.71) |
| Germany | 81.16 | 80.77 | 80.67 (80.50-80.83) | 83.67 | 83.37 | 83.32 (83.16-83.48) | 78.94 | 78.48 | 78.34 (78.17-78.51) |
| Israel | 82.51 | 82.12 | 81.85 (81.68-82.01) | 84.51 | 84.36 | 84.06 (83.90-84.22) | 80.67 | 80.05 | 79.82 (79.65-79.99) |
| Italy | 83.35 | 82.17 | 82.64 (82.49-82.81) | 85.40 | 84.43 | 84.82 (84.66-84.98) | 81.14 | 79.85 | 80.37 (80.21-80.54) |
| Netherlands | 82.05 | 81.25 | 81.35 (81.19-81.51) | 83.56 | 82.94 | 82.98 (82.83-83.14) | 80.46 | 79.55 | 79.70 (79.54-79.86) |
| New Zealand | 81.65 | 82.36 | 81.99 (81.83-82.15) | 83.49 | 84.16 | 83.75 (83.59-83.91) | 79.96 | 80.70 | 80.37 (80.20-80.53) |
| Northern Ireland | 80.92 | 79.83 | 79.99 (79.82-80.15) | 82.75 | 81.72 | 81.91 (81.75-82.07) | 79.00 | 77.91 | 78.05 (77.88-78.21) |
| Norway | 82.96 | 83.20 | 83.32 (83.16-83.48) | 84.70 | 84.89 | 84.88 (84.73-85.04) | 81.18 | 81.48 | 81.73 (81.57-81.89) |
| Portugal | 81.71 | 81.07 | 81.29 (81.13-81.45) | 84.56 | 84.00 | 84.24 (84.08-84.39) | 78.64 | 77.96 | 78.16 (78.00-78.33) |
| Scotland | 79.29 | 78.29 | 78.43 (78.27-78.61) | 81.26 | 80.56 | 80.49 (80.33-80.66) | 77.28 | 76.02 | 76.36 (76.19-76.53) |
| South Korea | 83.29 | 83.48 | 83.51 (83.35-83.67) | 86.30 | 86.47 | 86.51 (86.35-86.67) | 80.27 | 80.49 | 80.54 (80.38-80.71) |
| Spain | 83.56 | 82.30 | 83.16 (83.00-83.33) | 86.18 | 85.02 | 86.01 (85.85-86.17) | 80.82 | 79.55 | 80.38 (80.21-80.55) |
| Sweden | 83.06 | 82.43 | 83.22 (83.06-83.38) | 84.73 | 84.29 | 85.03 (84.87-85.18) | 81.35 | 80.60 | 81.40 (81.24-81.56) |
| Switzerland | 83.79 | 83.06 | 83.85 (83.69-84.01) | 85.57 | 85.08 | 85.92 (85.77-86.08) | 81.89 | 81.00 | 81.84 (81.68-82.00) |

**Note:** AI/AN = American Indian/Alaska Native; CR = credible range; NH = Non-Hispanic. Red shading indicates a decrease in life expectancy and green shading indicates an increase; significant decreases and increases in 2021 are determined by the CR, with no color applied to changes in 2021 that lacked significance (i.e., when 2020 values fell within the CR for 2021).

Examining the two years together, the overall decline in US life expectancy over two years between 2019 (78.85 years) and 2021 (76.44 [CR = 76.27-76.62] years) was substantial, a decrease of 2.41 (CR = 2.23-2.58) years. Losses were much larger in NH AI/AN, Hispanic, and NH Black populations (4.71 [CR = 4.49-4.94] years, 3.85 [CR = 3.67-4.03] years, and 3.70 [CR = 3.50-3.90, respectively) than in the NH White population (2.10 [CR = 1.93-2.28] years).

Compared to the United States, peer countries experienced smaller declines in life expectancy; on average, life expectancy in peer countries decreased by 0.55 years between 2019 and 2020 and then increased by 0.26 (CR = 0.10-0.42) years between 2020 and 2021 (**Figure 2**). As a result, the US-peer life expectancy gap grew from 3.37 years in 2019 to 4.69 years in 2020 and 5.49 (CR = 5.15-5.82) years in 2021.

**Figure 2.**
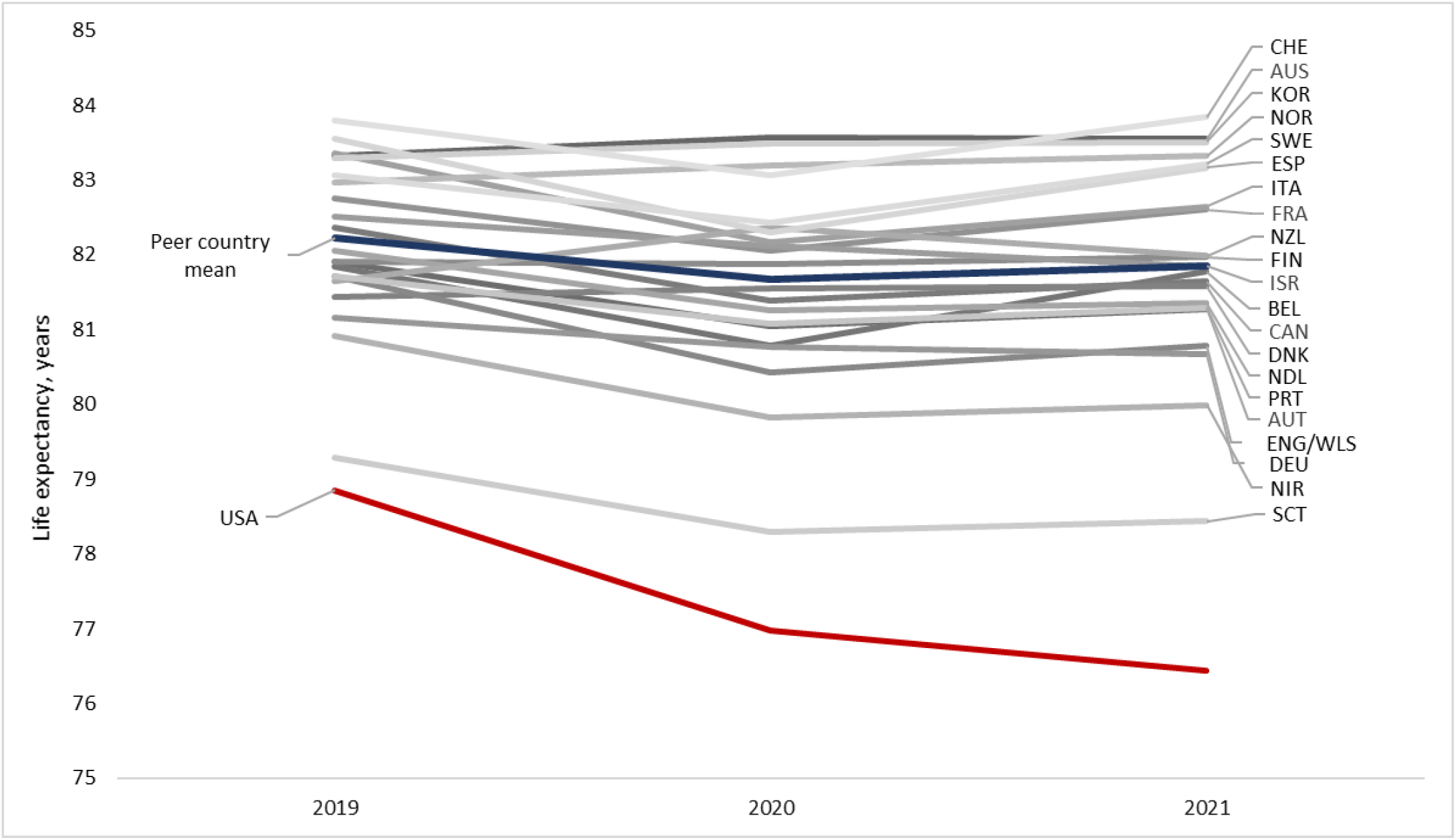
Life expectancy, 2019-2021, United States, 21 peer countries, and peer country average. Country codes and names: AUS (Australia), AUT (Austria), BEL (Belgium), CAN (Canada), CHE (Switzerland), DEU (Germany), DNK (Denmark), ENG/WLS (England and Wales), ESP (Spain), FIN (Finland), FRA (France), ISR (Israel), ITA (Italy), KOR (South Korea), NDL (Netherlands), NIR (Northern Ireland), NOR (Norway), NZL (New Zealand), PRT (Portugal), SCT (Scotland), SWE (Sweden), USA (United States). Life expectancy values for 2021 are estimated from provisional data; see table for credible ranges.

## DISCUSSION

Life expectancy is a summary measure of a population’s mortality experience. Comparing life expectancy changes between 2019 and 2021 reveals how the mortality consequences of the COVID-19 pandemic varied across countries. Compared to its peers, the US experienced much higher mortality rates and larger drops in life expectancy in both 2020 and 2021. Between 2019 and 2021, US life expectancy decreased by 2.41 (CR = 2.23-2.58) years, a decline not experienced since 1943, the deadliest year for Americans in World War II.^6^

Among peer countries, the largest loss in life expectancy between 2019 and 2021 was 0.93 (CR = 0.76-1.09) years in England and Wales (combined) and Northern Ireland (CR = 0.77-1.10), while four countries (Australia, New Zealand, Norway, and South Korea) gained life expectancy between 2019 and 2021. The gap between US life expectancy and the peer average rose to more than 5 years in 2021, further deepening a US disadvantage in health and survival that has been building for decades.^5^

Reasons for the surprising crossover in racialized US outcomes between 2020 and 2021—in which NH AI/AN, Hispanic, and NH Black populations saw large drops in life expectancy in 2020 but smaller losses in 2021 than those experienced by the NH White population—are not entirely clear and likely have multiple explanations.

Nonetheless, over the two-year period (2019-2021), NH AI/AN, Hispanic, and NH Black populations clearly experienced much larger losses in life expectancy than did the NH White population. These patterns reflect a long history of systemic racism and its attendant injustices and inadequacies in how the pandemic was managed in the United States.^7-9^ Although highly effective COVID-19 vaccines became available in 2021, their uptake was limited by public skepticism and inadequacies in distribution and access.

Limitations of this analysis include the reliance on provisional data for 2021 and extrapolation of population counts based on data from 2017-2020, cross-country variation in reporting of deaths, and the exclusion of some high-income countries (e.g., Japan).

## Data Availability

All data produced in the present work are contained in the manuscript.

## ACKNOWLEDGMENTS

The authors report no conflicts of interest. Dr. Masters received support from the University of Colorado Population Center grant from the Eunice Kennedy Shriver Institute of Child Health and Human Development (CUPC project 2P2CHD066613-06)..Dr. Woolf received partial funding from grant UL1TR002649 from the National Center for Advancing Translational Sciences. There was no specific funding for this study. The above funders had no role in the design and conduct of the study.

