## supplementary material for "CHANGES IN LIFE EXPECTANCY BETWEEN 2019 AND 2021 IN THE UNITED STATES AND 21 PEER COUNTRIES"

### Supplementary Material on Methods

1. Data sources for counts of death among U.S. populations, 2019-2020 and 2021

2. Data source for U.S. population estimates, 2019-2021

3. Data source for official U.S. life tables, 2019

4. Calculation of life tables for U.S. populations, 2020 and 2021

5. Data sources for counts of death among other high-income countries, 2018-2020 and 2021

6. Data sources for peer country population estimates, 2018-2021

7. Data sources for life tables for peer country populations, 2018, 2019, and/or 2020

8. Calculation of life tables for peer country populations, 2019, 2020, and/or 2021

9. Validation of estimates of U.S. and peer country life expectancy, 2019 and 2020

10. Simulation of 2021 life tables and life expectancy estimates

11. Figures for female life expectancy and male life expectancy changes, 2019-2020 and 2020-2021

12. Examples of analytic scripts in Stata and Python for merging data, simulating life tables, and estimating median values and credible ranges of 2021 life expectancy estimates

[1] Department of Sociology, University of Colorado Boulder, Boulder, CO
[2] Health Policy Center, Urban Institute, Washington, DC
[3] Center on Society and Health, Virginia Commonwealth University, Richmond, VA
Correspondence: Steven H. Woolf, MD, MPH, Center on Society and Health, Virginia Commonwealth University School of Medicine, 830 East Main Street, Suite 5035, Richmond, VA 23298-0212.

Below, we describe the data sources and methods used to estimate 2021 life expectancy among populations in the United States and other high-income countries. We use common notations for life table calculations. For example, the number of deaths occurring in an age interval *x* + n is commonly indicated by ${}_{n}d_{x}$, where x is the beginning of the age interval and n is the width of the age interval (e.g., n=1 for age interval 0-1, n=4 for age interval 1-4, and n=5 for five-year age intervals 5-9, ..., 95-99). The n is often omitted from the notation such that deaths occurring in age interval *x*+n are simply represented as $d_x$. Below, we **bold** notations to indicate values obtained from an official source (e.g., National Center for Health Statistics [NCHS] for U.S. values in 2019) or a reported source (e.g., Human Mortality Database for values among other high-income countries in 2018, 2019, or 2020). Notations that are not bolded indicate a value obtained from provisional sources and/or a value that we estimated using our methods. For example:

| | |
|---|---|
| Official counts of death in age *x* in year *t*: | $\boldsymbol{d_{x_t}}$ |
| Provisional counts of death in age *x* in year *t*: | $d_{x_t}$ |
| Official estimates of mid-year population in age *x* in year *t*: | $\boldsymbol{L_{x_t}}$ |
| Provisional estimates of mid-year population in age *x* in year *t*: | $L_{x_t}$ |
| Official estimates of death rate in age x in year t: | $\boldsymbol{m_{x_t}}$ |
| Provisional estimates of death rate in age x in year t: | $m_{x_t}$ |

#### 1. Data source for counts of death among U.S. populations, 2019-2020 and 2021

Official counts of death among U.S. populations in 2019 and 2020 ($\boldsymbol{d_{x_t}}$) were obtained from the National Center for Health Statistics' (NCHS) Restricted-Use Vital Statistics Data.[1] Provisional counts of death among U.S. populations in 2021 ($d_{x_{2021}}$) were obtained from the May 25, 2022 data release of the NCHS provisional mortality file, *AH Excess Deaths by Sex, Age, and Race.*[2]

#### 2. Data source for U.S. population estimates, 2019-2021

Mid-year population estimates in 2019 and 2020 ($\boldsymbol{L_{x_t}}$) by age, sex, and race/ethnicity were obtained from the U.S. Census Bureau file, Vintage 2020 Bridged-Race Population Estimates.[3] July 1 population counts for each year in 2019-2020 were summed at ages 0-1, 1-4, 5-9, ..., 95-99, 100+ and for 2021 were summed at ages 0-14, 15-19, ..., 80-84, 85+ to match the age structure of death counts reported in provisional NCHS data. U.S. population counts on July 1, 2021 ($L_{x_{2021}}$) were estimated from the linear trends of age-specific mid-year populations between years 2017 and 2020.

#### 3. Data source for official U.S. life tables, 2019

United States life tables for 2019 were obtained from Arias and Xu (2022).[4]

#### 4. Calculation of life tables for U.S. populations, 2020-2021

The $m_{x_t}$ for U.S. male and female populations was calculated by merging official counts of death by age ($\boldsymbol{d_{x_t}}$) for 19 age groups (0-1, 1-4, 5-9, ..., 95-99, 100+) with mid-year population counts by age ($\boldsymbol{L_{x_t}}$)[2]. We also estimated $m_{x_{2021}}$ by merging provisional death counts by age for 14 age groups (0-14, 15-19, ..., 80-84, 85+), $d_{x_{2021}}$, with $L_{x_{2021}}$ estimates. Using $m_{x_t}$, we estimated the 2021:2019 and 2020:2019 age-specific mortality rate ratios ($RR_{2021}$, $RR_{2020}$). To estimate official $\boldsymbol{m_{x_{2020}}}$ and $\boldsymbol{m_{x_{2021}}}$ used to generate 2020 and 2021 U.S. life tables, we combined age-specific $RR_{2020}$ and $RR_{2021}$ with $\boldsymbol{m_{x_{2019}}}$ obtained from the official 2019 US life tables, as follows:[4]

$$\boldsymbol{m_{x_{2020}}} = \boldsymbol{m_{x_{2019}}} * RR_{2020} \qquad \boldsymbol{m_{x_{2021}}} = \boldsymbol{m_{x_{2019}}} * RR_{2021}$$

To generate 2020 and 2021 U.S. life tables, we used $\boldsymbol{m_{x_{2020}}}$ and $\boldsymbol{m_{x_{2021}}}$ and $\boldsymbol{a_{x_{2019}}}$ (i.e., average time lived by the deceased in age interval *x*) obtained from the official 2019 life table to estimate $\boldsymbol{q_{x_{2020}}}$ and $\boldsymbol{q_{x_{2021}}}$ (i.e., probability of death at age *x*). For example, for the 2021 life table:

$$\boldsymbol{q_{x_{2021}}} = {(\boldsymbol{m_{x_{2021}}} * n)}\Big/{\left(1 + \left[(n - a_{x_{2019}}) * \boldsymbol{m_{x_{2021}}}\right]\right)}$$ where $n$ is the width of the age interval.

**5. Data sources for counts of death among other high-income countries, 2018-2021**

Death counts in 2018, 2019, 2020, and 2021 by sex and by age in each peer country were obtained from the May 23, 2022 release of the Human Mortality Database (HMD) - Short-term Mortality Fluctuations original input data in standardized format files.[5] The age-specificity of death counts varied across peer countries. For example, death counts in England and Wales (combined) for some years were provided across seven age groups of varying intervals (0-1, 1-14 15-44, 45-64, 65-74, 75-84, 85+), while death counts in Norway for all years were provided across 21 five-year age intervals (0-4, 5-9, ..., 95-99, 100+). See the HMD metadata for additional details about each peer country's reports of 2020 and 2021 deaths.

**6. Data sources for peer country population estimates, 2018-2021**

Population counts in 2018, 2019, 2020, and/or 2021 by sex and age were obtained from each country's central statistical agency:

Austria
Australia
Belgium
Denmark
Finland
France
Germany
Israel
Italy
Netherlands
Norway
Portugal
Spain
Sweden
Switzerland
UK - England and Wales
UK - Northern Ireland

UK - Scotland

Death counts for Canada, New Zealand, and South Korea were reported for wider age groups than the age groups published by the HMD-STMF (i.e., 0-14, 15-64, 65-74, 75-84, 85+), and therefore the HMD-STFM was used as the source for these countries' year-specific mortality rates, $m_{x_{2021}}$. Because we use $m_{x_{2021}}$ from HMD-STMF, population estimates were not obtained for these three countries.

### 7. Data sources for life tables for peer country populations, 2018, 2019, and/or 2020

Life tables in 2019 and 2020 for Belgium, Denmark, Finland, Norway, Portugal, Spain, Sweden, and Switzerland were obtained from the HMD, as was life expectancy in 2019 for Austria, Australia, France, Italy, and Netherlands.[6] Life tables in 2018 for the UK constituent countries England and Wales (combined), Northern Ireland, and Scotland were obtained from the HMD. Life tables for Germany, Israel, South Korea, and New Zealand were obtained from their central statistical agencies because they were not available for 2019 or 2020 in the HMD.

### 8. Calculation of life tables for peer country populations, 2019, 2020, and/or 2021

For life expectancies not provided in the HMD or not provided by each country's statistical agency, we generated 2019, 2020, and/or 2021 life tables in four steps. First, we estimated age-specific death rates ($m_{x_t}$) for each country's male and female population in years 2018, 2019, 2020, and 2021 using the HMD-STMF original input data in standardized format files (numerator) and age-specific mid-year population estimates (denominator) provided by each country's central statistical agency. Second, we estimated the 2021:2020, 2021:2019, or 2021:2018 age-specific mortality rate ratios ($RR_{2021,STMF}$) for each country's total, male, and female populations using the estimated $m_{x_t}$. If needed, we also estimated the 2020:2019 or 2020:2018 and 2019:2018 rate ratios. Third, to estimate $\boldsymbol{m}_{\boldsymbol{x}_{\boldsymbol{2021}}}$ for the 22 age groups used to generate 2021 life tables for most countries, we multiplied the age-specific $RR_{2021,STMF}$ by the age-specific 2018, 2019, and/or 2020 $m_{x_t}$ reported in each country's HMD life table, as follows:

$$\boldsymbol{m}_{\boldsymbol{x}_{\boldsymbol{2021}}} = m_{x_t} * RR_{2021,STMF}$$

where *t* is the most recently available life table and x = 0-1, 1-4, 5-9, ..., 100-104, 105-109, 110+

For Germany, South Korea, and New Zealand, $\boldsymbol{m}_{\boldsymbol{x}_{\boldsymbol{2021}}}$ was estimated as:

$$\boldsymbol{m}_{\boldsymbol{x}_{\boldsymbol{2021}}} = m_{x_t} * RR_{2021,STMF}$$

where *t* is the most recently available life table and x = 0-1, 1-4, 5-9, ..., 90-94, 95-99, 100+

This same step was used to estimate $\boldsymbol{m}_{\boldsymbol{x}_{\boldsymbol{2019}}}$ and $\boldsymbol{m}_{\boldsymbol{x}_{\boldsymbol{2020}}}$ for peer countries for which 2019 life tables (e.g., Scotland) or 2020 life tables (e.g., Austria) were not available in either the HMD or direct sources.

Fourth, to generate 2021 life tables for peer countries, we used $\boldsymbol{m}_{\boldsymbol{x}_{\boldsymbol{2021}}}$ and each country's $\boldsymbol{a}_{\boldsymbol{x}_{\boldsymbol{t}}}$ (i.e., average time lived by the deceased in age interval x) from the most recently available life table to estimate $\boldsymbol{q}_{\boldsymbol{x}_{\boldsymbol{2021}}}$ (i.e., probability of death at age x). For example, for 2021 in Denmark:

$$q_{x_{2021}} = \left(m_{x_{2021}} * n\right) \Big/ \left(1 + \left[\left(n - a_{x_{2020}}\right) * m_{x_{2021}}\right]\right)$$ where $n$ is the width of the age interval.

Where needed, we also generated 2019 and 2020 life tables for peer countries using $m_{x_{2019}}$ or $m_{x_{2020}}$ and each country's $a_{x_t}$ from the most recently available life table to estimate $q_{x_{2021}}$. For example, for 2020 in Austria:

$$q_{x_{2020}} = \left(m_{x_{2020}} * n\right) \Big/ \left(1 + \left[\left(n - a_{x_{2019}}\right) * m_{x_{2020}}\right]\right)$$ where $n$ is the width of the age interval.

**9. Simulation of 2021 life tables and life expectancy estimates**

Due to the provisional nature of the data used to estimate each country's 2021 life expectancy, we generated distributions of 2021 life expectancies for each country by adding 10% random uncertainty to $q_{x_{2021}}$ and 10% random uncertainty to $a_{x_{2021}}$ and using Python version 3.10.2 to simulate 50,000 life tables.[7] We report the median ($P_{50}$) estimate for each life expectancy as well as the fifth ($P_5$) and ninety-fifth ($P_{95}$) percentiles as plausible ranges for 2021 life expectancies. This "credible range" was calculated for the total, female, and male populations of each country, and also by race-ethnicity for the United States.

**10. Validation of life expectancy estimates for U.S. (2019 and 2020) and peer countries (2020)**

In the table below, we contrast values of 2019 U.S. life expectancy reported in the HMD and the official estimated reported by NCHS[7] with our estimates of 2019 U.S. life expectancy derived from our method using 2018 life tables. We also contrast our estimates with 2020 U.S. life expectancy estimates reported by NCHS;[8] the HMD does not currently report 2020 life expectancy for the United States. For 2020 estimates, we include the credible range as described above.

| | **2019 Life Expectancy** | | | **2020 Life Expectancy** | | |
|---|---|---|---|---|---|---|
| | HMD | NCHS*[7] | Estimate | NCHS*[8] | Estimate | Credible Range** |
| **United States** | | | | | | |
| Total | 79.16 | 78.8 | 78.86 | 77.0 | 76.99 | (76.83-77.16) |
| Male | 76.59 | 76.3 | 76.32 | 74.2 | 74.19 | (74.02-74.36) |
| Female | 81.72 | 81.4 | 81.39 | 79.9 | 79.88 | (79.72-80.04) |

* NCHS reports life expectancy to one decimal place only.
** Credible range = life expectancy values corresponding to the fifth ($P_5$) and ninety-fifth ($P_{95}$) percentiles from distributions of 50,000 simulated life tables.

U.S. life expectancy in 2019 reported in the HMD exceeds official NCHS reports of 2019 U.S. life expectancy by about 0.3 years (female) or 0.4 years (total).[7] Our method produces estimates that are nearly identical to those reported by NCHS (female: 81.39 vs. 81.4 and male: 76.32 vs. 76.3) or that fall between the values reported by the HMD and NCHS (total: 78.86 vs. 78.8 and 79.16). For 2020, NCHS estimates match our estimates exactly and fall in the middle of our credible range.

In the table below, we contrast HMD-reported male and female life expectancy for Belgium, Finland, Spain, and Switzerland in 2020 with life expectancy estimates derived from our method. For South Korea, we contrast male and female life expectancy reported by Korean Statistical Information Services (KOSIS) with life expectancy estimates derived from our method. For each country examined, the values of 2020 life expectancy reported in the HMD fall within our estimated credible range.

| | **2020 Life Expectancy** | | |
|---|---|---|---|
| | HMD | Estimate | Credible Range* |
| **Belgium** | | | |
| Male | 78.53 | 78.43 | (78.28-78.59) |
| Female | 83.06 | 82.96 | (82.81-83.11) |
| **Finland** | | | |
| Male | 79.08 | 79.05 | (78.89-79.21) |
| Female | 84.66 | 84.56 | (84.42-84.72) |
| **South Korea** | | | |
| Male | 80.49 | 80.43 | (80.27-80.59) |
| Female | 86.47 | 86.34 | (86.19-86.49) |
| **Spain** | | | |
| Male | 79.56 | 79.39 | (79.23-79.56) |
| Female | 85.02 | 84.87 | (84.73-85.02) |
| **Switzerland** | | | |
| Male | 81.00 | 80.87 | (80.72-81.02) |
| Female | 85.08 | 84.97 | (84.83-85.12) |

* Credible range = life expectancy values corresponding to the fifth ($P_5$) and ninety-fifth ($P_{95}$) percentiles from distributions of 50,000 simulated life tables.

**Panel A: Female**

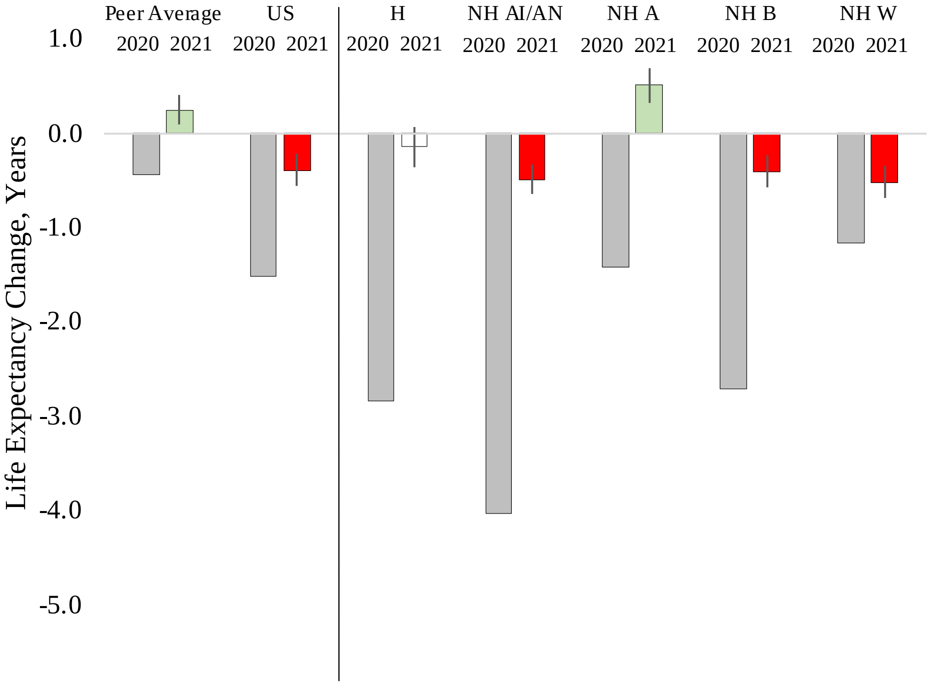


**Panel B: Male**

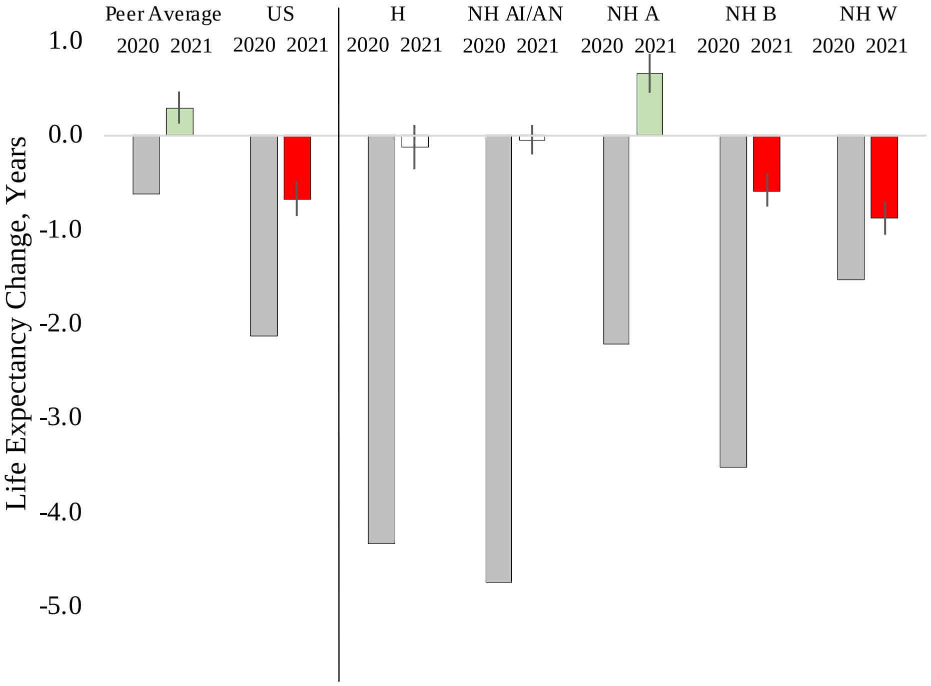


Notes: H = Hispanic, NH AI/AN = non-Hispanic American Indian/Alaska Native, NH A = non-Hispanic Asian, NH B = non-Hispanic Black, NH W = non-Hispanic White. 2019-2020 life expectancy change indicated by gray bar and 2020-2021 life expectancy change indicated by green bar (increase) or red bar (decrease). Vertical line indicates "credible range" of 2020-2021 life expectancy change.

**12. Examples of analytic scripts in Stata and Python for merging data, simulating life tables, and estimating median values and credible ranges of life expectancy estimates**

**Import HMD-STMF Death Counts: Austria, Belgium, United Kingdom as Examples**

```
*** Import HMDB Raw .txt Data ***

*** Peer Group 2021 LEs - HMD-STMF 5/23/22 Data

* Peer Group: 21 Countries: AUT, AUS, BEL, CAN, CHE, DEN, ESP, FIN, FRA, DEU,
KOR, ISR, ITA, NLD, NZL, NOR, POR, SWE, UK-ENG/W, UK-NIR, UK-SCO
* STMF RR: use for CAN, NZL, & KOR
* 2020 HMD LT = BEL, CHE, DEN, ESP, FIN, NOR, POR, SWE
* 2019 HMD LT = AUT, AUS, CAN, FRA, ITA, NLD (ISR & NZL Central Stats
Sources)
* 2018 HMD LT = DEU (Central Stats) UK-ENG/W, UK-NIR, UK-SCO
* 2020 & 2019 LT KOSIS = KOR
* 2021 LT generated from 2021:2020 RR (BEL, CHE, DEN, ESP, FIN, KOR, NOR,
POR, SWE), 2021:2019 RR (AUT, AUS, CAN, FRA, ITA, ISR, NLD, NZL), and from
2020:2018 (DEU, UK)

***************
*** Austria ***
***************

import delimited "/Users/ryan/Documents/Papers/LE
Trends_COVID/STMFinput_5_23_22/AUTstmf.csv", encoding(ISO-8859-1) clear

keep year week sex age deaths

keep if year == 2019 | year == 2020 | year == 2021

drop if age == "TOT"
drop if age == "UNK"

destring age, gen(age5)
drop age
rename age5 age

* Estimate Yearly Age-specific Death Count - Sum across the 52 weeks

sort sex year age

collapse (sum) deaths, by(sex year age)

gen nage = .
replace nage = 1 if age ==0
replace nage = 2 if age ==5
replace nage = 3 if age ==10
replace nage = 4 if age ==15
replace nage = 5 if age ==20
replace nage = 6 if age ==25
replace nage = 7 if age ==30
replace nage = 8 if age ==35
replace nage = 9 if age ==40
```

```
replace nage = 10 if age ==45
replace nage = 11 if age ==50
replace nage = 12 if age ==55
replace nage = 13 if age ==60
replace nage = 14 if age ==65
replace nage = 15 if age ==70
replace nage = 16 if age ==75
replace nage = 17 if age ==80
replace nage = 18 if age ==85
replace nage = 19 if age ==90

gen country = "Austria"

sort year sex nage

save "/Users/ryan/Documents/Papers/LE
Trends_COVID/STMFinput_5_23_22/austria_1921 deaths.dta", replace



***************
*** Belgium ***
***************

import delimited "/Users/ryan/Documents/Papers/LE
Trends_COVID/STMFinput_5_23_22/BELstmf.csv", encoding(ISO-8859-1) clear

keep year week sex age deaths

keep if year == 2020 | year == 2021

drop if age == "TOT"
drop if age == "UNK"

destring age, gen(age5)
drop age
rename age5 age

* Estimate Yearly Age-specific Death Count - Sum across the 52 weeks

sort sex year age

collapse (sum) deaths, by(sex year age)

gen nage = .
replace nage = 1 if age ==0
replace nage = 2 if age ==5
replace nage = 3 if age ==10
replace nage = 4 if age ==15
replace nage = 5 if age ==20
replace nage = 6 if age ==25
replace nage = 7 if age ==30
replace nage = 8 if age ==35
replace nage = 9 if age ==40
replace nage = 10 if age ==45
replace nage = 11 if age ==50
replace nage = 12 if age ==55
```

```
replace nage = 13 if age ==60
replace nage = 14 if age ==65
replace nage = 15 if age ==70
replace nage = 16 if age ==75
replace nage = 17 if age ==80
replace nage = 18 if age ==85
replace nage = 19 if age ==90

gen country = "Belgium"

sort year sex nage

save "/Users/ryan/Documents/Papers/LE
Trends_COVID/STMFinput_5_23_22/belgium_2021 deaths.dta", replace



***** United Kingdom: Northern Ireland, Scotland, England + Wales *****


***********************
*** Northern Ireland ***
***********************

****************************************
* Use full ages for 2018, 2019, and 2020 *
****************************************

import delimited "/Users/ryan/Documents/Papers/LE
Trends_COVID/STMFinput_5_23_22/GBR_NIRstmf.csv", encoding(ISO-8859-1) clear

keep year week sex age deaths

keep if year == 2018 | year == 2019 | year == 2020


drop if age == "TOT"
drop if age == "UNK"

destring age, gen(age5)
drop age
rename age5 age

collapse (sum) deaths, by(sex year age)

gen age20 = .
replace age20 = age
replace age20 = 0 if age20 == 1

sort sex year age20

collapse (sum) deaths, by(sex year age20)

rename age20 age

gen nage = .
replace nage = 1 if age ==0
```

```
replace nage = 2 if age ==5
replace nage = 3 if age ==10
replace nage = 4 if age ==15
replace nage = 5 if age ==20
replace nage = 6 if age ==25
replace nage = 7 if age ==30
replace nage = 8 if age ==35
replace nage = 9 if age ==40
replace nage = 10 if age ==45
replace nage = 11 if age ==50
replace nage = 12 if age ==55
replace nage = 13 if age ==60
replace nage = 14 if age ==65
replace nage = 15 if age ==70
replace nage = 16 if age ==75
replace nage = 17 if age ==80
replace nage = 18 if age ==85
replace nage = 19 if age ==90

gen country = "Northern Ireland - 2018-2020"

sort year sex nage

save "/Users/ryan/Documents/Papers/LE Trends_COVID/STMFinput_5_23_22/northern
ireland_1820 deaths.dta", replace


**************************************
* Use Trimmed Ages for 2018 and 2021 *
**************************************


import delimited "/Users/ryan/Documents/Papers/LE
Trends_COVID/STMFinput_5_23_22/GBR_NIRstmf.csv", encoding(ISO-8859-1) clear

keep year week sex age deaths

keep if year == 2018 | year == 2021

drop if age == "TOT"
drop if age == "UNK"

destring age, gen(age5)
drop age
rename age5 age

* Estimate Yearly Age-specific Death Count - Sum across the 52 weeks, but
2018 and 2021 have different age profiles

collapse (sum) deaths, by(sex year age)

gen age21 = .
replace age21 = age if year == 2021
replace age21 = 0 if age21 == 1
replace age21 = 0  if age < 15 & year == 2018
replace age21 = 15 if age >= 15 & age < 45 & year == 2018
replace age21 = 45 if age >= 45 & age < 65 & year == 2018
```

```
replace age21 = 65 if age >= 65 & age < 75 & year == 2018
replace age21 = 75 if age >= 75 & age < 85 & year == 2018
replace age21 = 85 if age >= 85 & year == 2018

sort sex year age21

collapse (sum) deaths, by(sex year age21)

rename age21 age

gen nage = .
replace nage = 1 if age ==0
replace nage = 2 if age ==15
replace nage = 3 if age ==45
replace nage = 4 if age ==65
replace nage = 5 if age ==75
replace nage = 6 if age ==85

gen country = "Northern Ireland"

sort year sex nage

save "/Users/ryan/Documents/Papers/LE Trends_COVID/STMFinput_5_23_22/northern
ireland_1821 deaths.dta", replace



****************
*** Scotland ***
****************

import delimited "/Users/ryan/Documents/Papers/LE
Trends_COVID/STMFinput_5_23_22/GBR_SCOstmf.csv", encoding(ISO-8859-1) clear

keep year week sex age deaths

keep if year == 2018 | year == 2019 | year == 2020 | year == 2021

drop if age == "TOT"
drop if age == "UNK"

destring age, gen(age5)
drop age
rename age5 age

* Estimate Yearly Age-specific Death Count - Sum across the 52 weeks

sort sex year age

collapse (sum) deaths, by(sex year age)


gen age21 = .
replace age21 = age
replace age21 = 0 if age21 == 1
replace age21 = 90 if age21 == 95
```

```
sort sex year age21

collapse (sum) deaths, by(sex year age21)

rename age21 age

gen nage = .
replace nage = 1 if age ==0
replace nage = 2 if age ==5
replace nage = 3 if age ==10
replace nage = 4 if age ==15
replace nage = 5 if age ==20
replace nage = 6 if age ==25
replace nage = 7 if age ==30
replace nage = 8 if age ==35
replace nage = 9 if age ==40
replace nage = 10 if age ==45
replace nage = 11 if age ==50
replace nage = 12 if age ==55
replace nage = 13 if age ==60
replace nage = 14 if age ==65
replace nage = 15 if age ==70
replace nage = 16 if age ==75
replace nage = 17 if age ==80
replace nage = 18 if age ==85
replace nage = 19 if age ==90

gen country = "Scotland"

sort year sex nage

save "/Users/ryan/Documents/Papers/LE
Trends_COVID/STMFinput_5_23_22/scotland_1821 deaths.dta", replace



***********************
*** England & Wales ***
***********************

import delimited "/Users/ryan/Documents/Papers/LE
Trends_COVID/STMFinput_5_23_22/GBRTENWstmf.csv", encoding(ISO-8859-1) clear

keep year week sex age deaths

keep if year == 2018 | year == 2019 | year == 2020 | year == 2021

drop if age == "TOT"
drop if age == "UNK"

destring age, gen(age5)
drop age
rename age5 age


* Estimate Yearly Age-specific Death Count - Sum across the 52 weeks, but
2018/2019 and 2020/2021 have different age profiles
```

```
collapse (sum) deaths, by(sex year age)

gen age20 = .
replace age20 = age if year == 2018
replace age20 = age if year == 2019
replace age20 = 0 if age == 1 & year == 2018
replace age20 = 0 if age == 1 & year == 2019
replace age20 = 0  if age < 15 & year == 2020
replace age20 = 0  if age < 15 & year == 2021
replace age20 = 15 if age >= 15 & age < 45 & year == 2020
replace age20 = 15 if age >= 15 & age < 45 & year == 2021
replace age20 = 45 if age >= 45 & age < 65 & year == 2020
replace age20 = 45 if age >= 45 & age < 65 & year == 2021
replace age20 = 65 if age >= 65 & age < 75 & year == 2020
replace age20 = 65 if age >= 65 & age < 75 & year == 2021
replace age20 = 75 if age >= 75 & age < 85 & year == 2020
replace age20 = 75 if age >= 75 & age < 85 & year == 2021
replace age20 = 85 if age >= 85 & year == 2020
replace age20 = 85 if age >= 85 & year == 2021

sort sex year age20

collapse (sum) deaths, by(sex year age20)

rename age20 age

gen nage = .
replace nage = 1 if age ==0
replace nage = 2 if age ==15
replace nage = 3 if age ==45
replace nage = 4 if age ==65
replace nage = 5 if age ==75
replace nage = 6 if age ==85

gen country = "England & Wales"

sort year sex nage

save "/Users/ryan/Documents/Papers/LE
Trends_COVID/STMFinput_5_23_22/england_wales_1821 deaths.dta", replace
```

**Merge HMD-STMF Death Counts with Peer Population Counts: Austria, Australia, Belgium, Denmark, United Kingdom as Examples**

```
*** Merge Population 2018, 2019, 2020, 2021 with Death Counts from STMF
Original Source 2018, 2019, 2020, 2021 ***


***************
*** Austria ***
***************

import excel "/Users/ryan/Desktop/Dropbox_sync/Dropbox/My Mac (Ryan’s
MacBook)/Documents/Papers/LE Trends_COVID/Peer Pop_2021/pop
input/aut_pop_21.xlsx", sheet("Sheet1") firstrow clear

sort year sex nage

merge year sex nage using "/Users/ryan/Documents/Papers/LE
Trends_COVID/STMFinput_5_23_22/austria_1921 deaths.dta"

gen mx = (deaths/pop)*100000

drop _merge

* Mx Rate Ratio b/w 2020 and 2019 and b/w 2021 and 2019

tempfile a b c d

sort country

save `a'


keep if year == 2019

collapse (mean) mx_19=mx, by(sex nage)

save `b'



use `a', clear

sort sex nage

keep if year == 2020

collapse (mean) mx_20=mx, by(sex nage)

save `c'

merge using `b'

drop _merge

save `c', replace
```

```
use `a', clear

sort sex nage

keep if year == 2021

sort sex nage

save `d'

merge using `c'

drop _merge



* Estimate RR

gen rr21 = mx/mx_19
gen rr20 = mx_20/mx_19


save "/Users/ryan/Desktop/Dropbox_sync/Dropbox/My Mac (Ryan's
MacBook)/Documents/Papers/LE Trends_COVID/HMDB
data/peer_STMF/2021_2020_2019_2018 rate ratios_original source
data/austria_19.dta", replace


*****************
*** Australia ***
*****************

import excel "/Users/ryan/Desktop/Dropbox_sync/Dropbox/My Mac (Ryan's
MacBook)/Documents/Papers/LE Trends_COVID/Peer Pop_2021/pop
input/aus_pop_21.xlsx", sheet("Sheet1") firstrow clear

sort year sex nage

merge year sex nage using "/Users/ryan/Documents/Papers/LE
Trends_COVID/STMFinput_5_23_22/australia_1921 deaths.dta"

gen mx = (deaths/pop)*100000

drop _merge

* Mx Rate Ratio b/w 2020 and 2019 and b/w 2021 and 2019

tempfile a b c d

sort country

save `a'
```

```
keep if year == 2019

collapse (mean) mx_19=mx, by(sex nage)

save `b'


use `a', clear

sort sex nage

keep if year == 2020

collapse (mean) mx_20=mx, by(sex nage)

save `c'

merge using `b'

drop _merge

save `c', replace


use `a', clear

sort sex nage

keep if year == 2021

sort sex nage

save `d'

merge using `c'

drop _merge


* Estimate RR

gen rr21 = mx/mx_19
gen rr20 = mx_20/mx_19


save "/Users/ryan/Desktop/Dropbox_sync/Dropbox/My Mac (Ryan’s
MacBook)/Documents/Papers/LE Trends_COVID/HMDB
data/peer_STMF/2021_2020_2019_2018 rate ratios_original source
data/australia_19.dta", replace


***************
*** Belgium ***
***************
```

```
import excel "/Users/ryan/Desktop/Dropbox_sync/Dropbox/My Mac (Ryan’s MacBook)/Documents/Papers/LE Trends_COVID/Peer Pop_2021/pop input/bel_pop_21.xlsx", sheet("Sheet1") firstrow clear

sort year sex nage

merge year sex nage using "/Users/ryan/Documents/Papers/LE Trends_COVID/STMFinput_5_23_22/belgium_2021 deaths.dta"

gen mx = (deaths/pop)*100000

drop _merge

drop if year == .

* Mx Rate Ratio b/w 2021 and 2020

tempfile a b c

sort country

save `a'


keep if year == 2020

collapse (mean) mx_20=mx, by(sex nage)

save `b'


use `a', clear

sort sex nage

keep if year == 2021

sort sex nage

save `c'

merge using `b'

drop _merge


* Estimate RR

gen rr21 = mx/mx_20

save "/Users/ryan/Desktop/Dropbox_sync/Dropbox/My Mac (Ryan’s MacBook)/Documents/Papers/LE Trends_COVID/HMDB data/peer_STMF/2021_2020_2019_2018 rate ratios_original source data/belgium_20.dta", replace
```

```
***************
*** Denmark ***
***************

import excel "/Users/ryan/Desktop/Dropbox_sync/Dropbox/My Mac (Ryan’s
MacBook)/Documents/Papers/LE Trends_COVID/Peer Pop_2021/pop
input/den_pop_21.xlsx", sheet("Sheet1") firstrow clear

sort year sex nage

merge year sex nage using "/Users/ryan/Documents/Papers/LE
Trends_COVID/STMFinput_5_23_22/denmark_2021 deaths.dta"

gen mx = (deaths/pop)*100000

drop _merge

* Mx Rate Ratio b/w 2021 and 2020

tempfile a b c

sort country

save `a'


keep if year == 2020

collapse (mean) mx_20=mx, by(sex nage)

save `b'


use `a', clear

sort sex nage

keep if year == 2021

sort sex nage

save `c'

merge using `b'

drop _merge


* Estimate RR

gen rr21 = mx/mx_20

save "/Users/ryan/Desktop/Dropbox_sync/Dropbox/My Mac (Ryan’s
MacBook)/Documents/Papers/LE Trends_COVID/HMDB
data/peer_STMF/2021_2020_2019_2018 rate ratios_original source
data/denmark_20.dta", replace
```

```
***********************
*** England & Wales ***
***********************

import excel "/Users/ryan/Desktop/Dropbox_sync/Dropbox/My Mac (Ryan’s
MacBook)/Documents/Papers/LE Trends_COVID/Peer Pop_2021/pop
input/eng_w_pop_21.xlsx", sheet("Sheet1") firstrow clear

sort year sex nage

merge year sex nage using "/Users/ryan/Documents/Papers/LE
Trends_COVID/STMFinput_3_28_22/england_wales_1821 deaths.dta"

gen mx = (deaths/pop)*100000

drop _merge

* Mx Rate Ratio b/w 2021 and 2018 and b/w 2020 and 2018 and b/w 2019 and 2018

tempfile a b c d

sort country

save `a'


keep if year == 2018

collapse (mean) mx_18=mx, by(sex nage)

save `b'

use `a', clear

sort sex nage

keep if year == 2019

collapse (mean) mx_19=mx, by(sex nage)

save `c'

merge using `b'

drop _merge

save `c', replace


use `a', clear

sort sex nage

keep if year == 2020

collapse (mean) mx_20=mx, by(sex nage)
merge using `c'
```

```
drop _merge

save `d'


use `a', clear

sort sex nage

keep if year == 2021

merge using `d'


* Estimate RR

gen rr21 = mx/mx_18
gen rr20 = mx_20/mx_18
gen rr19 = mx_19/mx_18


save "/Users/ryan/Desktop/Dropbox_sync/Dropbox/My Mac (Ryan’s
MacBook)/Documents/Papers/LE Trends_COVID/HMDB
data/peer_STMF/2021_2020_2019_2018 rate ratios_original source
data/england_wales_18.dta", replace
```

**Import NCHS U.S. Death Counts, 2019-2020, and Merge with Census Population Estimates**

```
***** Load 2019 and 2020 Restricted-Access NCHS Mortality Files *****
***** Collapse into 0, 1-4, 5-9, ..., 95-99, 100+ Age Groups by sex and
race/eth *****
***** Merge with CDC Wonder Population Counts: Vintage 2020 NCHS Abridged-
Race, 2019 and 2020... 0, 1-4, 5-9, ..., 95-99, 100+ *****


****************
* Loading Data *
****************

cd "E:\Data\20211209.Masters\Ryan Masters_12-9-2021\MortAC2019"


*************
*    2019   *
* Sex, Race *
*************

infix str state 33-34 ra 450 mra 489-490 hisp 488 str sex 69 age 75-76 str
cod_icd10 146-149 cod113 154-156 countcond 341-342 str cond1 344-348 str
cond2 349-353 str cond3 354-358 str cond4 359-363 str cond5 364-368 using
"MULT2019US.AllCnty.txt", clear

compress

des

* gen mort

gen mort = 1

* drop foreign residents

drop if state == "YY"
drop if state == "ZZ"
drop if state == "VI"
drop if state == "PR"
drop if state == "MX"
drop if state == "GU"
drop if state == "CU"
drop if state == "AS"
drop if state == "CC"

gen female = 0
replace female = 1 if sex == "F"

* race/ethnicity
* 1=Hispanic
* 2=NH Black
* 3=NH white
* 4=NH AIAN
* 5=NH Asian/NH/PI
```

```
gen hispanic = .
replace hispanic = 1 if hisp >=1 & hisp <=5
replace hispanic = 0 if hisp >= 6 & hisp <= 8

gen race = .
replace race = 1 if hispanic == 1
replace race = 2 if hispanic == 0 & ra == 2
replace race = 3 if hispanic == 0 & ra == 1
replace race = 4 if hispanic == 0 & ra == 3
replace race = 5 if hispanic == 0 & ra == 4
replace race = 5 if hispanic == 0 & ra == 5

tab race hisp

gen agecat = .
replace agecat = 0  if age >= 1 & age <= 22
replace agecat = 1  if age >= 23 & age <= 26
replace agecat = 5  if age == 27
replace agecat = 10 if age == 28
replace agecat = 15 if age == 29
replace agecat = 20 if age == 30
replace agecat = 25 if age == 31
replace agecat = 30 if age == 32
replace agecat = 35 if age == 33
replace agecat = 40 if age == 34
replace agecat = 45 if age == 35
replace agecat = 50 if age == 36
replace agecat = 55 if age == 37
replace agecat = 60 if age == 38
replace agecat = 65 if age == 39
replace agecat = 70 if age == 40
replace agecat = 75 if age == 41
replace agecat = 80 if age == 42
replace agecat = 85 if age == 43
replace agecat = 90 if age == 44
replace agecat = 95 if age == 45
replace agecat = 100 if age >= 46 & age <=51


* Collapse Death Counts - by Sex, Race, Age Category

sort female race agecat

collapse (sum) death=mort, by(female race agecat)

rename agecat age
drop if race == .
drop if age == .

* Save 2019 Death Counts - By Sex, Race/Ethnicity, Age

gen year = 2019

sort year female race age

save "E:\Data\covid2020\data_nation\death_sex_race_2019.dta", replace
```

```
********
* 2019 *
* Race *
********

infix str state 33-34 ra 450 mra 489-490 hisp 488 str sex 69 age 75-76 str
cod_icd10 146-149 cod113 154-156 countcond 341-342 str cond1 344-348 str
cond2 349-353 str cond3 354-358 str cond4 359-363 str cond5 364-368 using
"MULT2019US.AllCnty.txt", clear

compress

des

* gen mort

gen mort = 1


* drop foreign residents

drop if state == "YY"
drop if state == "ZZ"
drop if state == "VI"
drop if state == "PR"
drop if state == "MX"
drop if state == "GU"
drop if state == "CU"
drop if state == "AS"
drop if state == "CC"

gen female = 0
replace female = 1 if sex == "F"

* race/ethnicity
* 1=Hispanic
* 2=NH Black
* 3=NH white
* 4=NH AIAN
* 5=NH Asian/NH/PI

gen hispanic = .
replace hispanic = 1 if hisp >=1 & hisp <=5
replace hispanic = 0 if hisp >= 6 & hisp <= 8

gen race = .
replace race = 1 if hispanic == 1
replace race = 2 if hispanic == 0 & ra == 2
replace race = 3 if hispanic == 0 & ra == 1
replace race = 4 if hispanic == 0 & ra == 3
replace race = 5 if hispanic == 0 & ra == 4
replace race = 5 if hispanic == 0 & ra == 5

tab race hisp

gen agecat = .
replace agecat = 0  if age >= 1 & age <= 22
```

```
replace agecat = 1  if age >= 23 & age <= 26
replace agecat = 5  if age == 27
replace agecat = 10 if age == 28
replace agecat = 15 if age == 29
replace agecat = 20 if age == 30
replace agecat = 25 if age == 31
replace agecat = 30 if age == 32
replace agecat = 35 if age == 33
replace agecat = 40 if age == 34
replace agecat = 45 if age == 35
replace agecat = 50 if age == 36
replace agecat = 55 if age == 37
replace agecat = 60 if age == 38
replace agecat = 65 if age == 39
replace agecat = 70 if age == 40
replace agecat = 75 if age == 41
replace agecat = 80 if age == 42
replace agecat = 85 if age == 43
replace agecat = 90 if age == 44
replace agecat = 95 if age == 45
replace agecat = 100 if age >= 46 & age <=51

* Collapse Death Counts - by Race, Age Category

sort race agecat

collapse (sum) death=mort, by(race agecat)

rename agecat age
drop if race == .
drop if age == .

* Save 2019 Death Counts - By Race/Ethnicity, Age

gen year = 2019

sort year race age

save "E:\Data\covid2020\data_nation\death_race_2019.dta", replace


********
* 2019 *
*  Sex *
********

infix str state 33-34 ra 450 mra 489-490 hisp 488 str sex 69 age 75-76 str
cod_icd10 146-149 cod113 154-156 countcond 341-342 str cond1 344-348 str
cond2 349-353 str cond3 354-358 str cond4 359-363 str cond5 364-368 using
"MULT2019US.AllCnty.txt", clear

compress

des

* gen mort
```

```
gen mort = 1


* drop foreign residents

drop if state == "YY"
drop if state == "ZZ"
drop if state == "VI"
drop if state == "PR"
drop if state == "MX"
drop if state == "GU"
drop if state == "CU"
drop if state == "AS"
drop if state == "CC"

gen female = 0
replace female = 1 if sex == "F"

gen agecat = .
replace agecat = 0  if age >= 1 & age <= 22
replace agecat = 1  if age >= 23 & age <= 26
replace agecat = 5  if age == 27
replace agecat = 10 if age == 28
replace agecat = 15 if age == 29
replace agecat = 20 if age == 30
replace agecat = 25 if age == 31
replace agecat = 30 if age == 32
replace agecat = 35 if age == 33
replace agecat = 40 if age == 34
replace agecat = 45 if age == 35
replace agecat = 50 if age == 36
replace agecat = 55 if age == 37
replace agecat = 60 if age == 38
replace agecat = 65 if age == 39
replace agecat = 70 if age == 40
replace agecat = 75 if age == 41
replace agecat = 80 if age == 42
replace agecat = 85 if age == 43
replace agecat = 90 if age == 44
replace agecat = 95 if age == 45
replace agecat = 100 if age >= 46 & age <=51


* Collapse Death Counts - by Sex, Age Category

sort sex agecat

collapse (sum) death=mort, by(female agecat)

rename agecat age
drop if age == .

* Save 2019 Death Counts - By Sex, Age

gen year = 2019

sort year female age
```

```
save "E:\Data\covid2020\data_nation\death_sex_2019.dta", replace


*********
* 2019  *
* Total *
*********

infix str state 33-34 ra 450 mra 489-490 hisp 488 str sex 69 age 75-76 str
cod_icd10 146-149 cod113 154-156 countcond 341-342 str cond1 344-348 str
cond2 349-353 str cond3 354-358 str cond4 359-363 str cond5 364-368 using
"MULT2019US.AllCnty.txt", clear

compress

des

* gen mort

gen mort = 1


* drop foreign residents

drop if state == "YY"
drop if state == "ZZ"
drop if state == "VI"
drop if state == "PR"
drop if state == "MX"
drop if state == "GU"
drop if state == "CU"
drop if state == "AS"
drop if state == "CC"

gen agecat = .
replace agecat = 0  if age >= 1 & age <= 22
replace agecat = 1  if age >= 23 & age <= 26
replace agecat = 5  if age == 27
replace agecat = 10 if age == 28
replace agecat = 15 if age == 29
replace agecat = 20 if age == 30
replace agecat = 25 if age == 31
replace agecat = 30 if age == 32
replace agecat = 35 if age == 33
replace agecat = 40 if age == 34
replace agecat = 45 if age == 35
replace agecat = 50 if age == 36
replace agecat = 55 if age == 37
replace agecat = 60 if age == 38
replace agecat = 65 if age == 39
replace agecat = 70 if age == 40
replace agecat = 75 if age == 41
replace agecat = 80 if age == 42
replace agecat = 85 if age == 43
replace agecat = 90 if age == 44
replace agecat = 95 if age == 45
```

```
replace agecat = 100 if age >= 46 & age <=51


* Collapse Death Counts - by Age Category

sort agecat

collapse (sum) death=mort, by(agecat)

rename agecat age
drop if age == .

* Save 2019 Death Counts - By Age

gen year = 2019

sort year age

save "E:\Data\covid2020\data_nation\death_2019.dta", replace



cd "E:\Data\20211209.Masters\Ryan Masters_12-9-2021\MULT2020.LimGeo"


*************
*    2020   *
* Sex, Race *
*************

infix str state 33-34 ra 450 mra 489-490 hisp 488 str sex 69 age 75-76 str
cod_icd10 146-149 cod113 154-156 countcond 341-342 str cond1 344-348 str
cond2 349-353 str cond3 354-358 str cond4 359-363 str cond5 364-368 using
"Mort2020US.LimGeo.txt", clear

compress

des

* gen mort

gen mort = 1

* drop foreign residents

drop if state == "YY"
drop if state == "ZZ"
drop if state == "VI"
drop if state == "PR"
drop if state == "MX"
drop if state == "GU"
drop if state == "CU"
drop if state == "AS"
drop if state == "CC"

gen female = 0
replace female = 1 if sex == "F"
```

```
* race/ethnicity
* 1=Hispanic
* 2=NH Black
* 3=NH white
* 4=NH AIAN
* 5=NH Asian/NH/PI

gen hispanic = .
replace hispanic = 1 if hisp >=1 & hisp <=5
replace hispanic = 0 if hisp >= 6 & hisp <= 8

gen race = .
replace race = 1 if hispanic == 1
replace race = 2 if hispanic == 0 & ra == 2
replace race = 3 if hispanic == 0 & ra == 1
replace race = 4 if hispanic == 0 & ra == 3
replace race = 5 if hispanic == 0 & ra == 4
replace race = 5 if hispanic == 0 & ra == 5

tab race hisp

gen agecat = .
replace agecat = 0  if age >= 1 & age <= 22
replace agecat = 1  if age >= 23 & age <= 26
replace agecat = 5  if age == 27
replace agecat = 10 if age == 28
replace agecat = 15 if age == 29
replace agecat = 20 if age == 30
replace agecat = 25 if age == 31
replace agecat = 30 if age == 32
replace agecat = 35 if age == 33
replace agecat = 40 if age == 34
replace agecat = 45 if age == 35
replace agecat = 50 if age == 36
replace agecat = 55 if age == 37
replace agecat = 60 if age == 38
replace agecat = 65 if age == 39
replace agecat = 70 if age == 40
replace agecat = 75 if age == 41
replace agecat = 80 if age == 42
replace agecat = 85 if age == 43
replace agecat = 90 if age == 44
replace agecat = 95 if age == 45
replace agecat = 100 if age >= 46 & age <=51


* Collapse Death Counts - by Sex, Race, Age Category

sort female race agecat

collapse (sum) death=mort, by(female race agecat)

rename agecat age
drop if race == .
drop if age == .
```

```
* Save 2020 Death Counts - By Sex, Race/Ethnicity, Age

gen year = 2020

sort year female race age

save "E:\Data\covid2020\data_nation\death_sex_race_2020.dta", replace


********
* 2020 *
* Race *
********

infix str state 33-34 ra 450 mra 489-490 hisp 488 str sex 69 age 75-76 str
cod_icd10 146-149 cod113 154-156 countcond 341-342 str cond1 344-348 str
cond2 349-353 str cond3 354-358 str cond4 359-363 str cond5 364-368 using
"Mort2020US.LimGeo.txt", clear

compress

des

* gen mort

gen mort = 1


* drop foreign residents

drop if state == "YY"
drop if state == "ZZ"
drop if state == "VI"
drop if state == "PR"
drop if state == "MX"
drop if state == "GU"
drop if state == "CU"
drop if state == "AS"
drop if state == "CC"

gen female = 0
replace female = 1 if sex == "F"

* race/ethnicity
* 1=Hispanic
* 2=NH Black
* 3=NH white
* 4=NH AIAN
* 5=NH Asian/NH/PI

gen hispanic = .
replace hispanic = 1 if hisp >=1 & hisp <=5
replace hispanic = 0 if hisp >= 6 & hisp <= 8

gen race = .
replace race = 1 if hispanic == 1
replace race = 2 if hispanic == 0 & ra == 2
```

```
replace race = 3 if hispanic == 0 & ra == 1
replace race = 4 if hispanic == 0 & ra == 3
replace race = 5 if hispanic == 0 & ra == 4
replace race = 5 if hispanic == 0 & ra == 5

tab race hisp

gen agecat = .
replace agecat = 0  if age >= 1 & age <= 22
replace agecat = 1  if age >= 23 & age <= 26
replace agecat = 5  if age == 27
replace agecat = 10 if age == 28
replace agecat = 15 if age == 29
replace agecat = 20 if age == 30
replace agecat = 25 if age == 31
replace agecat = 30 if age == 32
replace agecat = 35 if age == 33
replace agecat = 40 if age == 34
replace agecat = 45 if age == 35
replace agecat = 50 if age == 36
replace agecat = 55 if age == 37
replace agecat = 60 if age == 38
replace agecat = 65 if age == 39
replace agecat = 70 if age == 40
replace agecat = 75 if age == 41
replace agecat = 80 if age == 42
replace agecat = 85 if age == 43
replace agecat = 90 if age == 44
replace agecat = 95 if age == 45
replace agecat = 100 if age >= 46 & age <=51


* Collapse Death Counts - by Race, Age Category

sort race agecat

collapse (sum) death=mort, by(race agecat)

rename agecat age
drop if race == .
drop if age == .

* Save 2020 Death Counts - By Race/Ethnicity, Age

gen year = 2020

sort year race age

save "E:\Data\covid2020\data_nation\death_race_2020.dta", replace


********
* 2020 *
*  Sex *
********
```

```
infix str state 33-34 ra 450 mra 489-490 hisp 488 str sex 69 age 75-76 str
cod_icd10 146-149 cod113 154-156 countcond 341-342 str cond1 344-348 str
cond2 349-353 str cond3 354-358 str cond4 359-363 str cond5 364-368 using
"Mort2020US.LimGeo.txt", clear

compress

des

* gen mort

gen mort = 1


* drop foreign residents

drop if state == "YY"
drop if state == "ZZ"
drop if state == "VI"
drop if state == "PR"
drop if state == "MX"
drop if state == "GU"
drop if state == "CU"
drop if state == "AS"
drop if state == "CC"

gen female = 0
replace female = 1 if sex == "F"

gen agecat = .
replace agecat = 0  if age >= 1 & age <= 22
replace agecat = 1  if age >= 23 & age <= 26
replace agecat = 5  if age == 27
replace agecat = 10 if age == 28
replace agecat = 15 if age == 29
replace agecat = 20 if age == 30
replace agecat = 25 if age == 31
replace agecat = 30 if age == 32
replace agecat = 35 if age == 33
replace agecat = 40 if age == 34
replace agecat = 45 if age == 35
replace agecat = 50 if age == 36
replace agecat = 55 if age == 37
replace agecat = 60 if age == 38
replace agecat = 65 if age == 39
replace agecat = 70 if age == 40
replace agecat = 75 if age == 41
replace agecat = 80 if age == 42
replace agecat = 85 if age == 43
replace agecat = 90 if age == 44
replace agecat = 95 if age == 45
replace agecat = 100 if age >= 46 & age <=51


* Collapse Death Counts - by Sex, Age Category

sort sex agecat
```

```
collapse (sum) death=mort, by(female agecat)

rename agecat age
drop if age == .

* Save 2020 Death Counts - By Sex, Age

gen year = 2020

sort year female age

save "E:\Data\covid2020\data_nation\death_sex_2020.dta", replace


*********
* 2020  *
* Total *
*********

infix str state 33-34 ra 450 mra 489-490 hisp 488 str sex 69 age 75-76 str
cod_icd10 146-149 cod113 154-156 countcond 341-342 str cond1 344-348 str
cond2 349-353 str cond3 354-358 str cond4 359-363 str cond5 364-368 using
"Mort2020US.LimGeo.txt", clear

compress

des

* gen mort

gen mort = 1

* drop foreign residents

drop if state == "YY"
drop if state == "ZZ"
drop if state == "VI"
drop if state == "PR"
drop if state == "MX"
drop if state == "GU"
drop if state == "CU"
drop if state == "AS"
drop if state == "CC"

gen agecat = .
replace agecat = 0  if age >= 1 & age <= 22
replace agecat = 1  if age >= 23 & age <= 26
replace agecat = 5  if age == 27
replace agecat = 10 if age == 28
replace agecat = 15 if age == 29
replace agecat = 20 if age == 30
replace agecat = 25 if age == 31
replace agecat = 30 if age == 32
replace agecat = 35 if age == 33
replace agecat = 40 if age == 34
replace agecat = 45 if age == 35
```

```
replace agecat = 50 if age == 36
replace agecat = 55 if age == 37
replace agecat = 60 if age == 38
replace agecat = 65 if age == 39
replace agecat = 70 if age == 40
replace agecat = 75 if age == 41
replace agecat = 80 if age == 42
replace agecat = 85 if age == 43
replace agecat = 90 if age == 44
replace agecat = 95 if age == 45
replace agecat = 100 if age >= 46 & age <=51


* Collapse Death Counts - by Age Category

sort agecat

collapse (sum) death=mort, by(agecat)

rename agecat age
drop if age == .

* Save 2020 Death Counts - By Age

gen year = 2020

sort year age

save "E:\Data\covid2020\data_nation\death_2020.dta", replace


**************************************
*        Append 2019 & 2020          *
* Merge Death Counts with Pop Counts *
**************************************

use "E:\Data\covid2020\data_nation\death_2019.dta", replace
append using "E:\Data\covid2020\data_nation\death_2020.dta"

sort year age
save "E:\Data\covid2020\data_nation\total_death.dta", replace


use "E:\Data\covid2020\data_nation\death_sex_2019.dta", replace
append using "E:\Data\covid2020\data_nation\death_sex_2020.dta"

sort year female age
save "E:\Data\covid2020\data_nation\sex_death.dta", replace


use "E:\Data\covid2020\data_nation\death_race_2019.dta", replace
append using "E:\Data\covid2020\data_nation\death_race_2020.dta"

sort year race age
save "E:\Data\covid2020\data_nation\race_death.dta", replace
```

```
use "E:\Data\covid2020\data_nation\death_sex_race_2019.dta", replace
append using "E:\Data\covid2020\data_nation\death_sex_race_2020.dta"

sort year race female age
save "E:\Data\covid2020\data_nation\sex_race_death.dta", replace


* Merge Death + Pop

use "E:\Data\covid2020\data_nation\pop\sex race pop_1920.dta", replace
sort year race female age
merge year race female age using
"E:\Data\covid2020\data_nation\sex_race_death.dta"

gen mx = (death/pop)*100000

save "E:\Data\covid2020\data_nation\sex_race_mx.dta", replace


use "E:\Data\covid2020\data_nation\pop\sex pop_1920.dta", replace
sort year female age
merge year female age using "E:\Data\covid2020\data_nation\sex_death.dta"

gen mx = (death/pop)*100000

save "E:\Data\covid2020\data_nation\sex_mx.dta", replace


use "E:\Data\covid2020\data_nation\pop\race pop_1920.dta", replace
sort year race age
merge year race age using "E:\Data\covid2020\data_nation\race_death.dta"

gen mx = (death/pop)*100000

save "E:\Data\covid2020\data_nation\race_mx.dta", replace


use "E:\Data\covid2020\data_nation\pop\total pop_1920.dta", replace
sort year age
merge year age using "E:\Data\covid2020\data_nation\total_death.dta"

gen mx = (death/pop)*100000

save "E:\Data\covid2020\data_nation\total_mx.dta", replace
```

**Import NCHS Provisional Death Counts, 2021, and Merge with Census Population Estimates**

```
********* All US Pop ************

*** 2021 NCHS Mortality Data, by Week ***

import delimited "/Users/ryan/Desktop/Dropbox_sync/Dropbox/My Mac (Ryan's
MacBook)/Documents/Papers/LE Trends_COVID/NCHS
Data/AH_Excess_Deaths_by_Sex__Age__and_Race_5_25_22.csv", encoding(ISO-8859-
1) clear

drop numberaboveaverageweighted percentaboveaverageweighted
numberaboveaverageunweighted percentaboveaverageunweighted timeperiod
analysisdate weekending

keep if mmwryear == 2021

keep if raceethnicity == "All Race/Ethnicity Groups"

drop covid19weighted covid19unweighted averagenumberofdeathsweighted
averagenumberofdeathsunweighted

keep if sex == "All Sexes"

encode agegroup, gen(age)

drop if age == 17 | age == 18


*** Sum Deaths for Entire 2021 by Age ***

sort age

collapse (sum) mort = deathsunweighted, by(age)

save "/Users/ryan/Documents/Papers/LE Trends_COVID/NCHS Data/total pop_age
specific death counts_2021.dta", replace


****** Import 2021 Pop Estimates ******

* Linear Trend Approximations from Age-specific Populations, Census Vintage
2020 Bridged-Race: 2017-2020
* All US Pop, 0-14, 5-9, ... ,95-99, 100+

import excel "/Users/ryan/Documents/Papers/LE
Trends_COVID/pop_cdc/pop_21.xlsx", sheet("total") firstrow clear

gen agecat = age
replace agecat = 16 if agecat >=16

collapse (sum) pop=pop, by(agecat)

rename agecat age
sort age
```

```
merge using "/Users/ryan/Documents/Papers/LE Trends_COVID/NCHS Data/total pop_age specific death counts_2021.dta"

drop _merge

gen mx = mort/pop

save "/Users/ryan/Documents/Papers/LE Trends_COVID/NCHS Data/total pop_2021 mx.dta", replace


********* All Male and All Female ************

*** 2021 NCHS Mortality Data, by Week ***

import delimited "/Users/ryan/Desktop/Dropbox_sync/Dropbox/My Mac (Ryan's MacBook)/Documents/Papers/LE Trends_COVID/NCHS Data/AH_Excess_Deaths_by_Sex__Age__and_Race_5_25_22.csv", encoding(ISO-8859-1) clear

drop numberaboveaverageweighted percentaboveaverageweighted numberaboveaverageunweighted percentaboveaverageunweighted timeperiod analysisdate weekending

keep if mmwryear == 2021

keep if raceethnicity == "All Race/Ethnicity Groups"

drop covid19weighted covid19unweighted averagenumberofdeathsweighted averagenumberofdeathsunweighted

encode sex, gen(female)
encode agegroup, gen(age)

drop if female == 1
replace female = 1 if female == 2
replace female = 0 if female == 3

label define femalel 0 "male" 1 "female"
label values female femalel

drop if age == 17 | age == 18


*** Sum Deaths for Entire 2021 by Age for Men and Women ***

sort female age

collapse (sum) mort = deathsunweighted, by(female age)

sort female age

save "/Users/ryan/Documents/Papers/LE Trends_COVID/NCHS Data/total sex_age specific death counts_2021.dta", replace


****** Import 2021 Pop Estimates ******
```

```
* Linear Trend Approximations from Age-specific Populations, CDC Wonder Pop:
2017-2020
* All US Pop, 0-14, 5-9, ... ,80-84, 85+

import excel "/Users/ryan/Documents/Papers/LE
Trends_COVID/pop_cdc/pop_21.xlsx", sheet("sex") firstrow clear

gen agecat = age
replace agecat = 16 if agecat >=16

collapse (sum) pop=pop, by(female agecat)

rename agecat age
sort age

sort female age

merge using "/Users/ryan/Documents/Papers/LE Trends_COVID/NCHS Data/total
sex_age specific death counts_2021.dta"

drop _merge

gen mx = mort/pop

save "/Users/ryan/Documents/Papers/LE Trends_COVID/NCHS Data/total sex_2021
mx.dta", replace


*** 2021 NCHS Mortality Data, by Week ***

import delimited "/Users/ryan/Desktop/Dropbox_sync/Dropbox/My Mac (Ryan's
MacBook)/Documents/Papers/LE Trends_COVID/NCHS
Data/AH_Excess_Deaths_by_Sex__Age__and_Race_5_25_22.csv", encoding(ISO-8859-
1) clear

drop numberaboveaverageweighted percentaboveaverageweighted
numberaboveaverageunweighted percentaboveaverageunweighted timeperiod
analysisdate weekending

keep if mmwryear == 2021

drop covid19weighted covid19unweighted averagenumberofdeathsweighted
averagenumberofdeathsunweighted


keep if sex == "All Sexes"

encode agegroup, gen(age)
encode raceethnicity, gen(race_e)

drop if age == 17 | age == 18

tab race_e
tab race_e, nolabel

* Race/Eth
```

```
* 1=Hispanic
* 2=NH Black
* 3=NH White
* 4=NH AIAN
* 5=NH Asian

gen race = .
replace race = 1 if race_e == 2
replace race = 2 if race_e == 5
replace race = 3 if race_e == 7
replace race = 4 if race_e == 3
replace race = 5 if race_e == 4
replace race = 5 if race_e == 6

drop if race == .

label define racel 1 "Hispanic" 2 "NH Black" 3 "NH White" 4 "NH AIAN" 5 "NH
Asian"
label values race racel

*** Sum Deaths for Entire 2020 by Age for Race/Eth Pops Total ***

sort race  age

collapse (sum) mort = deathsunweighted, by(race age)

sort race age

save "/Users/ryan/Documents/Papers/LE Trends_COVID/NCHS Data/race_age
specific death counts_2021.dta", replace


****** Import 2021 Pop Estimates ******

* Linear Trend Approximations from Age-specific Populations, CDC Wonder Pop:
2017-2020
* All US Pop, 0-14, 5-9, ... ,80-84, 85+

* race
* 1=Hisp
* 2=NH B
* 3=NH W
* 4=NH AIAN
* 5=NH A

import excel "/Users/ryan/Documents/Papers/LE
Trends_COVID/pop_cdc/pop_21.xlsx", sheet("race") firstrow clear

gen agecat = age
replace agecat = 16 if agecat >=16

collapse (sum) pop=pop, by(race agecat)

rename agecat age

sort race age
```

```
merge using "/Users/ryan/Documents/Papers/LE Trends_COVID/NCHS Data/race_age specific death counts_2021.dta"

drop _merge

gen mx = mort/pop

save "/Users/ryan/Documents/Papers/LE Trends_COVID/NCHS Data/race_2021 mx.dta", replace


********* Sex X Race/Ethnicity ************

*** 2021 NCHS Mortality Data, by Week ***

import delimited "/Users/ryan/Desktop/Dropbox_sync/Dropbox/My Mac (Ryan’s MacBook)/Documents/Papers/LE Trends_COVID/NCHS Data/AH_Excess_Deaths_by_Sex__Age__and_Race_5_25_22.csv", encoding(ISO-8859-1) clear

drop numberaboveaverageweighted percentaboveaverageweighted numberaboveaverageunweighted percentaboveaverageunweighted timeperiod analysisdate weekending

keep if mmwryear == 2021

drop averagenumberofdeathsweighted averagenumberofdeathsunweighted

encode sex, gen(female)
encode agegroup, gen(age)
encode raceethnicity, gen(race_e)

drop if female == 1
replace female = 1 if female == 2
replace female = 0 if female == 3

label define femalel 0 "male" 1 "female"
label values female femalel

drop if age == 17 | age == 18


tab race_e
tab race_e, nolabel

* Race/Eth
* 1=Hispanic
* 2=NH Black
* 3=NH White
* 4=NH AIAN
* 5=NH Asian

gen race = .
replace race = 1 if race_e == 2
replace race = 2 if race_e == 5
replace race = 3 if race_e == 7
replace race = 4 if race_e == 3
```

```
replace race = 5 if race_e == 4
replace race = 5 if race_e == 6

drop if race == .

label define racel 1 "Hispanic" 2 "NH Black" 3 "NH White" 4 "NH AIAN" 5 "NH
Asian"
label values race racel


*** Sum Deaths for Entire 2021 by Age for NWB and NHW Men and Women ***

sort race female age

collapse (sum) mort = deathsunweighted, by(race female age)

sort female race age

save "/Users/ryan/Documents/Papers/LE Trends_COVID/NCHS Data/race_sex_age
specific death counts_2021.dta", replace


****** Import 2021 Pop Estimates ******

* Linear Trend Approximations from Age-specific Populations, CDC Wonder Pop:
2017-2020
* All US Pop, 0-14, 5-9, ... ,80-84, 85+

import excel "/Users/ryan/Documents/Papers/LE
Trends_COVID/pop_cdc/pop_21.xlsx", sheet("race_sex") firstrow clear


gen agecat = age
replace agecat = 16 if agecat >=16

collapse (sum) pop=pop, by(female race agecat)

rename agecat age

sort female race age

merge using "/Users/ryan/Documents/Papers/LE Trends_COVID/NCHS
Data/race_sex_age specific death counts_2021.dta"

drop _merge

gen mx = mort/pop

save "/Users/ryan/Documents/Papers/LE Trends_COVID/NCHS Data/race_sex_2021
mx.dta", replace
```

### 2021 Life Tables: U.S. Female Population, Germany Male Population, Spain Total Population

### U.S. Female Population, 2021

```
# -*- coding: utf-8 -*-
"""
Created May 26, 2022

Use AH Excess Death NCHS Mortality Data, 5/25 Release + restricted-access US 2020 Vintage Population Data + 2019 LT
Simulate 50K life tables with random uncertainty on qx and ax
Estimate mx for five age groups

@author: ryan masters


"""
#import packages
import random

# importing in the qx and error and ax
female_t = r"/Users/ryan/Documents/Papers/LE Trends_COVID/2021 Life Expectancy/simulations/us/female_t.txt"
# change as needed for input files

# read in the file
textFile = open(female_t,'r')
text = textFile.readlines()
# split into different age categories
a0=text[1]
a1=text[2]
a5=text[3]
a10=text[4]
a15=text[5]
a20=text[6]
a25=text[7]
a30=text[8]
a35=text[9]
a40=text[10]
a45=text[11]
a50=text[12]
a55=text[13]
a60=text[14]
a65=text[15]
a70=text[16]
a75=text[17]
a80=text[18]
a85=text[19]
a90=text[20]
a95=text[21]
a100=text[22]


a0_sp = a0.split(",")
a1_sp = a1.split(",")
a5_sp = a5.split(",")
a10_sp = a10.split(",")
a15_sp = a15.split(",")
a20_sp = a20.split(",")
a25_sp = a25.split(",")
a30_sp = a30.split(",")
```

```
a35_sp = a35.split(",")
a40_sp = a40.split(",")
a45_sp = a45.split(",")
a50_sp = a50.split(",")
a55_sp = a55.split(",")
a60_sp = a60.split(",")
a65_sp = a65.split(",")
a70_sp = a70.split(",")
a75_sp = a75.split(",")
a80_sp = a80.split(",")
a85_sp = a85.split(",")
a90_sp = a90.split(",")
a95_sp = a95.split(",")
a100_sp = a100.split(",")

# qx
a0_qx = float(a0_sp[1])
a1_qx = float(a1_sp[1])
a5_qx = float(a5_sp[1])
a10_qx = float(a10_sp[1])
a15_qx = float(a15_sp[1])
a20_qx = float(a20_sp[1])
a25_qx = float(a25_sp[1])
a30_qx = float(a30_sp[1])
a35_qx = float(a35_sp[1])
a40_qx = float(a40_sp[1])
a45_qx = float(a45_sp[1])
a50_qx = float(a50_sp[1])
a55_qx = float(a55_sp[1])
a60_qx = float(a60_sp[1])
a65_qx = float(a65_sp[1])
a70_qx = float(a70_sp[1])
a75_qx = float(a75_sp[1])
a80_qx = float(a80_sp[1])
a85_qx = float(a85_sp[1])
a90_qx = float(a90_sp[1])
a95_qx = float(a95_sp[1])
a100_qx = float(a100_sp[1])

# qx - LB
a0_qxl = float(a0_sp[2])
a1_qxl = float(a1_sp[2])
a5_qxl = float(a5_sp[2])
a10_qxl = float(a10_sp[2])
a15_qxl = float(a15_sp[2])
a20_qxl = float(a20_sp[2])
a25_qxl = float(a25_sp[2])
a30_qxl = float(a30_sp[2])
a35_qxl = float(a35_sp[2])
a40_qxl = float(a40_sp[2])
a45_qxl = float(a45_sp[2])
a50_qxl = float(a50_sp[2])
a55_qxl = float(a55_sp[2])
a60_qxl = float(a60_sp[2])
a65_qxl = float(a65_sp[2])
a70_qxl = float(a70_sp[2])
a75_qxl = float(a75_sp[2])
a80_qxl = float(a80_sp[2])
a85_qxl = float(a85_sp[2])
a90_qxl = float(a90_sp[2])
```

```
a95_qxl = float(a95_sp[2])
a100_qxl = float(a100_sp[2])

# qx - UB
a0_qxu = float(a0_sp[3])
a1_qxu = float(a1_sp[3])
a5_qxu = float(a5_sp[3])
a10_qxu = float(a10_sp[3])
a15_qxu = float(a15_sp[3])
a20_qxu = float(a20_sp[3])
a25_qxu = float(a25_sp[3])
a30_qxu = float(a30_sp[3])
a35_qxu = float(a35_sp[3])
a40_qxu = float(a40_sp[3])
a45_qxu = float(a45_sp[3])
a50_qxu = float(a50_sp[3])
a55_qxu = float(a55_sp[3])
a60_qxu = float(a60_sp[3])
a65_qxu = float(a65_sp[3])
a70_qxu = float(a70_sp[3])
a75_qxu = float(a75_sp[3])
a80_qxu = float(a80_sp[3])
a85_qxu = float(a85_sp[3])
a90_qxu = float(a90_sp[3])
a95_qxu = float(a95_sp[3])
a100_qxu = float(a100_sp[3])


# ax
a0_ax = float(a0_sp[4])
a1_ax = float(a1_sp[4])
a5_ax = float(a5_sp[4])
a10_ax = float(a10_sp[4])
a15_ax = float(a15_sp[4])
a20_ax = float(a20_sp[4])
a25_ax = float(a25_sp[4])
a30_ax = float(a30_sp[4])
a35_ax = float(a35_sp[4])
a40_ax = float(a40_sp[4])
a45_ax = float(a45_sp[4])
a50_ax = float(a50_sp[4])
a55_ax = float(a55_sp[4])
a60_ax = float(a60_sp[4])
a65_ax = float(a65_sp[4])
a70_ax = float(a70_sp[4])
a75_ax = float(a75_sp[4])
a80_ax = float(a80_sp[4])
a85_ax = float(a85_sp[4])
a90_ax = float(a90_sp[4])
a95_ax = float(a95_sp[4])
a100_ax = float(a100_sp[4])


# ax - LB
a0_axl = float(a0_sp[5])
a1_axl = float(a1_sp[5])
a5_axl = float(a5_sp[5])
a10_axl = float(a10_sp[5])
a15_axl = float(a15_sp[5])
a20_axl = float(a20_sp[5])
```

```
a25_axl = float(a25_sp[5])
a30_axl = float(a30_sp[5])
a35_axl = float(a35_sp[5])
a40_axl = float(a40_sp[5])
a45_axl = float(a45_sp[5])
a50_axl = float(a50_sp[5])
a55_axl = float(a55_sp[5])
a60_axl = float(a60_sp[5])
a65_axl = float(a65_sp[5])
a70_axl = float(a70_sp[5])
a75_axl = float(a75_sp[5])
a80_axl = float(a80_sp[5])
a85_axl = float(a85_sp[5])
a90_axl = float(a90_sp[5])
a95_axl = float(a95_sp[5])
a100_axl = float(a100_sp[5])


# ax - UB
a0_axu = float(a0_sp[6])
a1_axu = float(a1_sp[6])
a5_axu = float(a5_sp[6])
a10_axu = float(a10_sp[6])
a15_axu = float(a15_sp[6])
a20_axu = float(a20_sp[6])
a25_axu = float(a25_sp[6])
a30_axu = float(a30_sp[6])
a35_axu = float(a35_sp[6])
a40_axu = float(a40_sp[6])
a45_axu = float(a45_sp[6])
a50_axu = float(a50_sp[6])
a55_axu = float(a55_sp[6])
a60_axu = float(a60_sp[6])
a65_axu = float(a65_sp[6])
a70_axu = float(a70_sp[6])
a75_axu = float(a75_sp[6])
a80_axu = float(a80_sp[6])
a85_axu = float(a85_sp[6])
a90_axu = float(a90_sp[6])
a95_axu = float(a95_sp[6])
a100_axu = float(a100_sp[6])

# 2019 mx - 2019 LT
a0_m19 = float(a0_sp[7])
a1_m19 = float(a1_sp[7])
a5_m19 = float(a5_sp[7])
a10_m19 = float(a10_sp[7])
a15_m19 = float(a15_sp[7])
a20_m19 = float(a20_sp[7])
a25_m19 = float(a25_sp[7])
a30_m19 = float(a30_sp[7])
a35_m19 = float(a35_sp[7])
a40_m19 = float(a40_sp[7])
a45_m19 = float(a45_sp[7])
a50_m19 = float(a50_sp[7])
a55_m19 = float(a55_sp[7])
a60_m19 = float(a60_sp[7])
a65_m19 = float(a65_sp[7])
a70_m19 = float(a70_sp[7])
a75_m19 = float(a75_sp[7])
```

```
a80_m19 = float(a80_sp[7])
a85_m19 = float(a85_sp[7])
a90_m19 = float(a90_sp[7])
a95_m19 = float(a95_sp[7])
a100_m19 = float(a100_sp[7])


count = 0
while count < 50000:

# Random qx by age

#   uniform
    a0_rand_qx = random.uniform(a0_qxl,a0_qxu)
    a1_rand_qx = random.uniform(a1_qxl,a1_qxu)
    a5_rand_qx = random.uniform(a5_qxl,a5_qxu)
    a10_rand_qx = random.uniform(a10_qxl,a10_qxu)
    a15_rand_qx = random.uniform(a15_qxl,a15_qxu)
    a20_rand_qx = random.uniform(a20_qxl,a20_qxu)
    a25_rand_qx = random.uniform(a25_qxl,a25_qxu)
    a30_rand_qx = random.uniform(a30_qxl,a30_qxu)
    a35_rand_qx = random.uniform(a35_qxl,a35_qxu)
    a40_rand_qx = random.uniform(a40_qxl,a40_qxu)
    a45_rand_qx = random.uniform(a45_qxl,a45_qxu)
    a50_rand_qx = random.uniform(a50_qxl,a50_qxu)
    a55_rand_qx = random.uniform(a55_qxl,a55_qxu)
    a60_rand_qx = random.uniform(a60_qxl,a60_qxu)
    a65_rand_qx = random.uniform(a65_qxl,a65_qxu)
    a70_rand_qx = random.uniform(a70_qxl,a70_qxu)
    a75_rand_qx = random.uniform(a75_qxl,a75_qxu)
    a80_rand_qx = random.uniform(a80_qxl,a80_qxu)
    a85_rand_qx = random.uniform(a85_qxl,a85_qxu)
    a90_rand_qx = random.uniform(a90_qxl,a90_qxu)
    a95_rand_qx = random.uniform(a95_qxl,a95_qxu)
    a100_rand_qx = 1


# Random ax by age

#   uniform
    a0_rand_ax = random.uniform(a0_axl,a0_axu)
    a1_rand_ax = random.uniform(a1_axl,a1_axu)
    a5_rand_ax = random.uniform(a5_axl,a5_axu)
    a10_rand_ax = random.uniform(a10_axl,a10_axu)
    a15_rand_ax = random.uniform(a15_axl,a15_axu)
    a20_rand_ax = random.uniform(a20_axl,a20_axu)
    a25_rand_ax = random.uniform(a25_axl,a25_axu)
    a30_rand_ax = random.uniform(a30_axl,a30_axu)
    a35_rand_ax = random.uniform(a35_axl,a35_axu)
    a40_rand_ax = random.uniform(a40_axl,a40_axu)
    a45_rand_ax = random.uniform(a45_axl,a45_axu)
    a50_rand_ax = random.uniform(a50_axl,a50_axu)
    a55_rand_ax = random.uniform(a55_axl,a55_axu)
    a60_rand_ax = random.uniform(a60_axl,a60_axu)
    a65_rand_ax = random.uniform(a65_axl,a65_axu)
    a70_rand_ax = random.uniform(a70_axl,a70_axu)
    a75_rand_ax = random.uniform(a75_axl,a75_axu)
    a80_rand_ax = random.uniform(a80_axl,a80_axu)
    a85_rand_ax = random.uniform(a85_axl,a85_axu)
    a90_rand_ax = random.uniform(a90_axl,a90_axu)
```

```
    a95_rand_ax = random.uniform(a95_axl,a95_axu)
    a100_rand_ax = random.uniform(a100_axl,a100_axu)


### calculate life table variables
    radix = 1000000.0000000

    # calculate the number of deaths age0
    a0_dx = a0_rand_qx*radix
    # calculate survivors
    a0_lx=radix
    a0_sx=a0_lx/radix # this is 1?
    a1_lx=(radix-a0_dx)
    a1_sx = a1_lx/radix

    # calculate the number of deaths age1
    a1_dx = a1_rand_qx*a1_lx
    # calculate survivors
    a5_lx=(a1_lx-a1_dx)
    a5_sx = a5_lx/radix

    # calculate the number of deaths age5
    a5_dx = a5_rand_qx*a5_lx
    # calculate survivors
    a10_lx=(a5_lx-a5_dx)
    a10_sx = a10_lx/radix

    # calculate the number of deaths age10
    a10_dx = a10_rand_qx*a10_lx
    # calculate survivors
    a15_lx=(a10_lx-a10_dx)
    a15_sx = a15_lx/radix

    # calculate the number of deaths age15
    a15_dx = a15_rand_qx*a15_lx
    # calculate survivors
    a20_lx=(a15_lx-a15_dx)
    a20_sx = a20_lx/radix

    # calculate the number of deaths age20
    a20_dx = a20_rand_qx*a20_lx
    # calculate survivors
    a25_lx=(a20_lx-a20_dx)
    a25_sx = a25_lx/radix

    # calculate the number of deaths age25
    a25_dx = a25_rand_qx*a25_lx
    # calculate survivors
    a30_lx=(a25_lx-a25_dx)
    a30_sx = a30_lx/radix

    # calculate the number of deaths age30
    a30_dx = a30_rand_qx*a30_lx
    # calculate survivors
    a35_lx=(a30_lx-a30_dx)
    a35_sx = a35_lx/radix

    # calculate the number of deaths age35
    a35_dx = a35_rand_qx*a35_lx
    # calculate survivors
```

```
a40_lx=(a35_lx-a35_dx)
a40_sx = a40_lx/radix

# calculate the number of deaths age40
a40_dx = a40_rand_qx*a40_lx
# calculate survivors
a45_lx=(a40_lx-a40_dx)
a45_sx = a45_lx/radix

# calculate the number of deaths age45
a45_dx = a45_rand_qx*a45_lx
# calculate survivors
a50_lx=(a45_lx-a45_dx)
a50_sx = a50_lx/radix

# calculate the number of deaths age50
a50_dx = a50_rand_qx*a50_lx
# calculate survivors
a55_lx=(a50_lx-a50_dx)
a55_sx = a55_lx/radix

# calculate the number of deaths age55
a55_dx = a55_rand_qx*a55_lx
# calculate survivors
a60_lx=(a55_lx-a55_dx)
a60_sx = a60_lx/radix

# calculate the number of deaths age60
a60_dx = a60_rand_qx*a60_lx
# calculate survivors
a65_lx=(a60_lx-a60_dx)
a65_sx = a65_lx/radix

# calculate the number of deaths age65
a65_dx = a65_rand_qx*a65_lx
# calculate survivors
a70_lx=(a65_lx-a65_dx)
a70_sx = a70_lx/radix

# calculate the number of deaths age70
a70_dx = a70_rand_qx*a70_lx
# calculate survivors
a75_lx=(a70_lx-a70_dx)
a75_sx = a75_lx/radix

# calculate the number of deaths age75
a75_dx = a75_rand_qx*a75_lx
# calculate survivors
a80_lx=(a75_lx-a75_dx)
a80_sx = a80_lx/radix

# calculate the number of deaths age80
a80_dx = a80_rand_qx*a80_lx
# calculate survivors
a85_lx=(a80_lx-a80_dx)
a85_sx = a85_lx/radix

# calculate the number of deaths age85
a85_dx = a85_rand_qx*a85_lx
# calculate survivors
```

```
a90_lx=(a85_lx-a85_dx)
a90_sx = a90_lx/radix

# calculate the number of deaths age90
a90_dx = a90_rand_qx*a90_lx
# calculate survivors
a95_lx=(a90_lx-a90_dx)
a95_sx = a95_lx/radix

# calculate the number of deaths age95
a95_dx = a95_rand_qx*a95_lx
# calculate survivors
a100_lx=(a95_lx-a95_dx)
a100_sx = a100_lx/radix

# calculate the number of deaths age100
a100_dx = a100_rand_qx*a100_lx
# No Survivors - top-coded

#calculate Lx
a0_Lx = (a1_lx*1)+(a0_dx*a0_rand_ax)
a1_Lx = (a5_lx*4)+(a1_dx*a1_rand_ax)
a5_Lx = (a10_lx*5)+(a5_dx*a5_rand_ax)
a10_Lx = (a15_lx*5)+(a10_dx*a10_rand_ax)
a15_Lx = (a20_lx*5)+(a15_dx*a15_rand_ax)
a20_Lx = (a25_lx*5)+(a20_dx*a20_rand_ax)
a25_Lx = (a30_lx*5)+(a25_dx*a25_rand_ax)
a30_Lx = (a35_lx*5)+(a30_dx*a30_rand_ax)
a35_Lx = (a40_lx*5)+(a35_dx*a35_rand_ax)
a40_Lx = (a45_lx*5)+(a40_dx*a40_rand_ax)
a45_Lx = (a50_lx*5)+(a45_dx*a45_rand_ax)
a50_Lx = (a55_lx*5)+(a50_dx*a50_rand_ax)
a55_Lx = (a60_lx*5)+(a55_dx*a55_rand_ax)
a60_Lx = (a65_lx*5)+(a60_dx*a60_rand_ax)
a65_Lx = (a70_lx*5)+(a65_dx*a65_rand_ax)
a70_Lx = (a75_lx*5)+(a70_dx*a70_rand_ax)
a75_Lx = (a80_lx*5)+(a75_dx*a75_rand_ax)
a80_Lx = (a85_lx*5)+(a80_dx*a80_rand_ax)
a85_Lx = (a90_lx*5)+(a85_dx*a85_rand_ax)
a90_Lx = (a95_lx*5)+(a90_dx*a90_rand_ax)
a95_Lx = (a100_lx*5)+(a95_dx*a95_rand_ax)
a100_Lx = (a100_dx*a100_rand_ax)


####
# calculate Tx
a0_Tx =
a0_Lx+a1_Lx+a5_Lx+a10_Lx+a15_Lx+a20_Lx+a25_Lx+a30_Lx+a35_Lx+a40_Lx+a45_Lx+a50_Lx+a55_Lx+a60_Lx+a65_Lx+a70_L
x+a75_Lx+a80_Lx+a85_Lx+a90_Lx+a95_Lx+a100_Lx
a1_Tx =
a1_Lx+a5_Lx+a10_Lx+a15_Lx+a20_Lx+a25_Lx+a30_Lx+a35_Lx+a40_Lx+a45_Lx+a50_Lx+a55_Lx+a60_Lx+a65_Lx+a70_Lx+a75_
Lx+a80_Lx+a85_Lx+a90_Lx+a95_Lx+a100_Lx
a5_Tx =
a5_Lx+a10_Lx+a15_Lx+a20_Lx+a25_Lx+a30_Lx+a35_Lx+a40_Lx+a45_Lx+a50_Lx+a55_Lx+a60_Lx+a65_Lx+a70_Lx+a75_Lx+a80
_Lx+a85_Lx+a90_Lx+a95_Lx+a100_Lx
a10_Tx =
a10_Lx+a15_Lx+a20_Lx+a25_Lx+a30_Lx+a35_Lx+a40_Lx+a45_Lx+a50_Lx+a55_Lx+a60_Lx+a65_Lx+a70_Lx+a75_Lx+a80_Lx+a8
5_Lx+a90_Lx+a95_Lx+a100_Lx
```

```
   a15_Tx =
a15_Lx+a20_Lx+a25_Lx+a30_Lx+a35_Lx+a40_Lx+a45_Lx+a50_Lx+a55_Lx+a60_Lx+a65_Lx+a70_Lx+a75_Lx+a80_Lx+a85_Lx+a9
0_Lx+a95_Lx+a100_Lx
   a20_Tx =
a20_Lx+a25_Lx+a30_Lx+a35_Lx+a40_Lx+a45_Lx+a50_Lx+a55_Lx+a60_Lx+a65_Lx+a70_Lx+a75_Lx+a80_Lx+a85_Lx+a90_Lx+a9
5_Lx+a100_Lx
   a25_Tx =
a25_Lx+a30_Lx+a35_Lx+a40_Lx+a45_Lx+a50_Lx+a55_Lx+a60_Lx+a65_Lx+a70_Lx+a75_Lx+a80_Lx+a85_Lx+a90_Lx+a95_Lx+a1
00_Lx
   a30_Tx =
a30_Lx+a35_Lx+a40_Lx+a45_Lx+a50_Lx+a55_Lx+a60_Lx+a65_Lx+a70_Lx+a75_Lx+a80_Lx+a85_Lx+a90_Lx+a95_Lx+a100_Lx
   a35_Tx =
a35_Lx+a40_Lx+a45_Lx+a50_Lx+a55_Lx+a60_Lx+a65_Lx+a70_Lx+a75_Lx+a80_Lx+a85_Lx+a90_Lx+a95_Lx+a100_Lx
   a40_Tx = a40_Lx+a45_Lx+a50_Lx+a55_Lx+a60_Lx+a65_Lx+a70_Lx+a75_Lx+a80_Lx+a85_Lx+a90_Lx+a95_Lx+a100_Lx
   a45_Tx = a45_Lx+a50_Lx+a55_Lx+a60_Lx+a65_Lx+a70_Lx+a75_Lx+a80_Lx+a85_Lx+a90_Lx+a95_Lx+a100_Lx
   a50_Tx = a50_Lx+a55_Lx+a60_Lx+a65_Lx+a70_Lx+a75_Lx+a80_Lx+a85_Lx+a90_Lx+a95_Lx+a100_Lx
   a55_Tx = a55_Lx+a60_Lx+a65_Lx+a70_Lx+a75_Lx+a80_Lx+a85_Lx+a90_Lx+a95_Lx+a100_Lx
   a60_Tx = a60_Lx+a65_Lx+a70_Lx+a75_Lx+a80_Lx+a85_Lx+a90_Lx+a95_Lx+a100_Lx
   a65_Tx = a65_Lx+a70_Lx+a75_Lx+a80_Lx+a85_Lx+a90_Lx+a95_Lx+a100_Lx
   a70_Tx = a70_Lx+a75_Lx+a80_Lx+a85_Lx+a90_Lx+a95_Lx+a100_Lx
   a75_Tx = a75_Lx+a80_Lx+a85_Lx+a90_Lx+a95_Lx+a100_Lx
   a80_Tx = a80_Lx+a85_Lx+a90_Lx+a95_Lx+a100_Lx
   a85_Tx = a85_Lx+a90_Lx+a95_Lx+a100_Lx
   a90_Tx = a90_Lx+a95_Lx+a100_Lx
   a95_Tx = a95_Lx+a100_Lx
   a100_Tx = a100_Lx


   #calculate mx
   a0_mx = (a0_dx)/a0_Lx
   a1_mx = (a1_dx)/a1_Lx
   a5_mx = (a5_dx)/a5_Lx
   a10_mx = (a10_dx)/a10_Lx
   a15_mx = (a15_dx)/a15_Lx
   a20_mx = (a20_dx)/a20_Lx
   a25_mx = (a25_dx)/a25_Lx
   a30_mx = (a30_dx)/a30_Lx
   a35_mx = (a35_dx)/a35_Lx
   a40_mx = (a40_dx)/a40_Lx
   a45_mx = (a45_dx)/a45_Lx
   a50_mx = (a50_dx)/a50_Lx
   a55_mx = (a55_dx)/a55_Lx
   a60_mx = (a60_dx)/a60_Lx
   a65_mx = (a65_dx)/a65_Lx
   a70_mx = (a70_dx)/a70_Lx
   a75_mx = (a75_dx)/a75_Lx
   a80_mx = (a80_dx)/a80_Lx
   a85_mx = (a85_dx)/a85_Lx
   a90_mx = (a90_dx)/a90_Lx
   a95_mx = (a95_dx)/a95_Lx
   a100_mx = (a100_dx)/a100_Lx


   #calculate Lx 0-14, 15-39, 40-64, 65-84, 85+

   a015_tx = (a0_Lx+a1_Lx+a5_Lx+a10_Lx)
   a1539_tx = (a15_Lx+a20_Lx+a25_Lx+a30_Lx+a35_Lx)
   a4064_tx = (a40_Lx+a45_Lx+a50_Lx+a55_Lx+a60_Lx)
   a6584_tx = (a65_Lx+a70_Lx+a75_Lx+a80_Lx)
   a85_tx = a85_Tx
```

```
# simulation of weighted-averages of 2020:2019 mx ratio:0-15, 15-40, 40-65, 65-85, 85+

    a015_rr  =
(a0_mx/a0_m19)*(a0_Lx/a015_tx)+(a1_mx/a1_m19)*(a1_Lx/a015_tx)+(a5_mx/a5_m19)*(a5_Lx/a015_tx)+(a10_mx/a10_m19)*(a10_
Lx/a015_tx)
    a1539_rr =
(a15_mx/a15_m19)*(a15_Lx/a1539_tx)+(a20_mx/a20_m19)*(a20_Lx/a1539_tx)+(a25_mx/a25_m19)*(a25_Lx/a1539_tx)+(a30_mx/a
30_m19)*(a30_Lx/a1539_tx)+(a35_mx/a35_m19)*(a35_Lx/a1539_tx)
    a4064_rr =
(a40_mx/a40_m19)*(a40_Lx/a4064_tx)+(a45_mx/a45_m19)*(a45_Lx/a4064_tx)+(a50_mx/a50_m19)*(a50_Lx/a4064_tx)+(a55_mx/a
55_m19)*(a55_Lx/a4064_tx)+(a60_mx/a60_m19)*(a60_Lx/a4064_tx)
    a6584_rr =
(a65_mx/a65_m19)*(a65_Lx/a6584_tx)+(a70_mx/a70_m19)*(a70_Lx/a6584_tx)+(a75_mx/a75_m19)*(a75_Lx/a6584_tx)+(a80_mx/a
80_m19)*(a80_Lx/a6584_tx)
    a85100_rr   =
(a85_mx/a85_m19)*(a85_Lx/a85_tx)+(a90_mx/a90_m19)*(a90_Lx/a85_tx)+(a95_mx/a95_m19)*(a95_Lx/a85_tx)+(a100_mx/a100_m
19)*(a100_Lx/a85_tx)


# output the mortality rate ratios by age groups
    mxgrp = r"/Users/ryan/Documents/Papers/LE Trends_COVID/2021 Life Expectancy/simulations/us/female_t_rrgrp.txt"
    opened_file = open(mxgrp, 'a')

    if count==0:
        opened_file.write('{0} {1} {2} {3} {4} {5}\n'.format("sim_num","rr015","rr1539","rr4064","rr6584","rr85"))

    else:
        opened_file.write('{0} {1} {2} {3} {4} {5}\n'.format(count,a015_rr,a1539_rr,a4064_rr,a6584_rr,a85100_rr))


# simulation of 2020:2019 mx ratio

    a0_rr = a0_mx/a0_m19
    a1_rr = a1_mx/a1_m19
    a5_rr = a5_mx/a5_m19
    a10_rr = a10_mx/a10_m19
    a15_rr = a15_mx/a15_m19
    a20_rr = a20_mx/a20_m19
    a25_rr = a25_mx/a25_m19
    a30_rr = a30_mx/a30_m19
    a35_rr = a35_mx/a35_m19
    a40_rr = a40_mx/a40_m19
    a45_rr = a45_mx/a45_m19
    a50_rr = a50_mx/a50_m19
    a55_rr = a55_mx/a55_m19
    a60_rr = a60_mx/a60_m19
    a65_rr = a65_mx/a65_m19
    a70_rr = a70_mx/a70_m19
    a75_rr = a75_mx/a75_m19
    a80_rr = a80_mx/a80_m19
    a85_rr = a85_mx/a85_m19
    a90_rr = a90_mx/a90_m19
    a95_rr = a95_mx/a95_m19
    a100_rr = a100_mx/a100_m19


# output the mortality rate ratios for each sim
    rr = r"/Users/ryan/Documents/Papers/LE Trends_COVID/2021 Life Expectancy/simulations/us/female_t_rr.txt"
    opened_file = open(rr, 'a')
```

```
    if count==0:
        opened_file.write('{0} {1} {2} {3} {4} {5} {6} {7} {8} {9} {10} {11} {12} {13} {14} {15} {16} {17} {18} {19} {20} {21}
{22}\n'.format("sim_num","a0_rr","a1_rr","a5_rr","a10_rr","a15_rr","a20_rr","a25_rr","a30_rr","a35_rr","a40_rr","a45_rr","a50_rr","a55_r
r","a60_rr","a65_rr","a70_rr","a75_rr","a80_rr","a85_rr","a90_rr","a95_rr","a100_rr"))

    else:
        opened_file.write('{0} {1} {2} {3} {4} {5} {6} {7} {8} {9} {10} {11} {12} {13} {14} {15} {16} {17} {18} {19} {20} {21}
{22}\n'.format(count,a0_rr,a1_rr,a5_rr,a10_rr,a15_rr,a20_rr,a25_rr,a30_rr,a35_rr,a40_rr,a45_rr,a50_rr,a55_rr,a60_rr,a65_rr,a70_rr,a7
5_rr,a80_rr,a85_rr,a90_rr,a95_rr,a100_rr))

# output the mortality rates for each sim
    mx = r"/Users/ryan/Documents/Papers/LE Trends_COVID/2021 Life Expectancy/simulations/us/female_t_mx.txt"
    opened_file = open(mx, 'a')

    if count==0:
        opened_file.write('{0} {1} {2} {3} {4} {5} {6} {7} {8} {9} {10} {11} {12} {13} {14} {15} {16} {17} {18} {19} {20} {21}
{22}\n'.format("sim_num","a0_mx","a1_mx","a5_mx","a10_mx","a15_mx","a20_mx","a25_mx","a30_mx","a35_mx","a40_mx","a45_mx
","a50_mx","a55_mx","a60_mx","a65_mx","a70_mx","a75_mx","a80_mx","a85_mx","a90_mx","a95_mx","a100_mx"))

    else:
        opened_file.write('{0} {1} {2} {3} {4} {5} {6} {7} {8} {9} {10} {11} {12} {13} {14} {15} {16} {17} {18} {19} {20} {21}
{22}\n'.format(count,a0_mx,a1_mx,a5_mx,a10_mx,a15_mx,a20_mx,a25_mx,a30_mx,a35_mx,a40_mx,a45_mx,a50_mx,a55_mx,a60
_mx,a65_mx,a70_mx,a75_mx,a80_mx,a85_mx,a90_mx,a95_mx,a100_mx))



    ###### estimate life expectancy

    #calculate ex
    a0_ex = a0_Tx/radix
    a1_ex = a1_Tx/a1_lx
    a5_ex = a5_Tx/a5_lx
    a10_ex = a10_Tx/a10_lx
    a15_ex = a15_Tx/a15_lx
    a20_ex = a20_Tx/a20_lx
    a25_ex = a25_Tx/a25_lx
    a30_ex = a30_Tx/a30_lx
    a35_ex = a35_Tx/a35_lx
    a40_ex = a40_Tx/a40_lx
    a45_ex = a45_Tx/a45_lx
    a50_ex = a50_Tx/a50_lx
    a55_ex = a55_Tx/a55_lx
    a60_ex = a60_Tx/a60_lx
    a65_ex = a65_Tx/a65_lx
    a70_ex = a70_Tx/a70_lx
    a75_ex = a75_Tx/a75_lx
    a80_ex = a80_Tx/a80_lx
    a85_ex = a85_Tx/a85_lx
    a90_ex = a90_Tx/a90_lx
    a95_ex = a95_Tx/a95_lx
    a100_ex = a100_Tx/a100_lx

    # save data

    tot_file_name = r"/Users/ryan/Documents/Papers/LE Trends_COVID/2021 Life Expectancy/simulations/us/female_t_ex.txt"
    tot_opened_file = open(tot_file_name, 'a')
    #opened_file.write("%r\n" %age45_ex_total)
    if count==0:
        tot_opened_file.write('{0} {1} {2}\n'.format("sim_num","age", "ex"))
```

```
        tot_opened_file.write('{0} {1} {2}\n'.format(count,"0",a0_ex))
        tot_opened_file.write('{0} {1} {2}\n'.format(count,"1",a1_ex))
        tot_opened_file.write('{0} {1} {2}\n'.format(count,"5",a5_ex))
        tot_opened_file.write('{0} {1} {2}\n'.format(count,"10",a10_ex))
        tot_opened_file.write('{0} {1} {2}\n'.format(count,"15",a15_ex))
        tot_opened_file.write('{0} {1} {2}\n'.format(count,"20",a20_ex))
        tot_opened_file.write('{0} {1} {2}\n'.format(count,"25",a25_ex))
        tot_opened_file.write('{0} {1} {2}\n'.format(count,"30",a30_ex))
        tot_opened_file.write('{0} {1} {2}\n'.format(count,"35",a35_ex))
        tot_opened_file.write('{0} {1} {2}\n'.format(count,"40",a40_ex))
        tot_opened_file.write('{0} {1} {2}\n'.format(count,"45",a45_ex))
        tot_opened_file.write('{0} {1} {2}\n'.format(count,"50",a50_ex))
        tot_opened_file.write('{0} {1} {2}\n'.format(count,"55",a55_ex))
        tot_opened_file.write('{0} {1} {2}\n'.format(count,"60",a60_ex))
        tot_opened_file.write('{0} {1} {2}\n'.format(count,"65",a65_ex))
        tot_opened_file.write('{0} {1} {2}\n'.format(count,"70",a70_ex))
        tot_opened_file.write('{0} {1} {2}\n'.format(count,"75",a75_ex))
        tot_opened_file.write('{0} {1} {2}\n'.format(count,"80",a80_ex))
        tot_opened_file.write('{0} {1} {2}\n'.format(count,"85",a85_ex))
        tot_opened_file.write('{0} {1} {2}\n'.format(count,"90",a90_ex))
        tot_opened_file.write('{0} {1} {2}\n'.format(count,"95",a95_ex))
        tot_opened_file.write('{0} {1} {2}\n'.format(count,"100",a100_ex))


    else:
        tot_opened_file.write('{0} {1} {2}\n'.format(count,"0",a0_ex))
        tot_opened_file.write('{0} {1} {2}\n'.format(count,"1",a1_ex))
        tot_opened_file.write('{0} {1} {2}\n'.format(count,"5",a5_ex))
        tot_opened_file.write('{0} {1} {2}\n'.format(count,"10",a10_ex))
        tot_opened_file.write('{0} {1} {2}\n'.format(count,"15",a15_ex))
        tot_opened_file.write('{0} {1} {2}\n'.format(count,"20",a20_ex))
        tot_opened_file.write('{0} {1} {2}\n'.format(count,"25",a25_ex))
        tot_opened_file.write('{0} {1} {2}\n'.format(count,"30",a30_ex))
        tot_opened_file.write('{0} {1} {2}\n'.format(count,"35",a35_ex))
        tot_opened_file.write('{0} {1} {2}\n'.format(count,"40",a40_ex))
        tot_opened_file.write('{0} {1} {2}\n'.format(count,"45",a45_ex))
        tot_opened_file.write('{0} {1} {2}\n'.format(count,"50",a50_ex))
        tot_opened_file.write('{0} {1} {2}\n'.format(count,"55",a55_ex))
        tot_opened_file.write('{0} {1} {2}\n'.format(count,"60",a60_ex))
        tot_opened_file.write('{0} {1} {2}\n'.format(count,"65",a65_ex))
        tot_opened_file.write('{0} {1} {2}\n'.format(count,"70",a70_ex))
        tot_opened_file.write('{0} {1} {2}\n'.format(count,"75",a75_ex))
        tot_opened_file.write('{0} {1} {2}\n'.format(count,"80",a80_ex))
        tot_opened_file.write('{0} {1} {2}\n'.format(count,"85",a85_ex))
        tot_opened_file.write('{0} {1} {2}\n'.format(count,"90",a90_ex))
        tot_opened_file.write('{0} {1} {2}\n'.format(count,"95",a95_ex))
        tot_opened_file.write('{0} {1} {2}\n'.format(count,"100",a100_ex))


    print(count)
    count += 1  # This is the same as count = count + 1

tot_opened_file.close()

print("simulation completed")
```

### Germany Male Population, 2021

```
# -*- coding: utf-8 -*-
"""
Created 5/23

Peer 2021 Life Tables from 2018/2019/2020 qx*2020:2018RR and 2018 ax

@author: ryan masters


"""
#import packages
import random

# importing in the qx and error and ax
deu_m = r"/Users/ryan/Documents/Papers/LE Trends_COVID/2021 Life Expectancy/simulations/peer/male/deu_m.txt"
# change as needed for input files

# read in the file
textFile = open(deu_m,'r')
text = textFile.readlines()
# split into different age categories
a0=text[1]
a1=text[2]
a5=text[3]
a10=text[4]
a15=text[5]
a20=text[6]
a25=text[7]
a30=text[8]
a35=text[9]
a40=text[10]
a45=text[11]
a50=text[12]
a55=text[13]
a60=text[14]
a65=text[15]
a70=text[16]
a75=text[17]
a80=text[18]
a85=text[19]
a90=text[20]
a95=text[21]
a100=text[22]


a0_sp = a0.split(",")
a1_sp = a1.split(",")
a5_sp = a5.split(",")
a10_sp = a10.split(",")
a15_sp = a15.split(",")
a20_sp = a20.split(",")
a25_sp = a25.split(",")
a30_sp = a30.split(",")
a35_sp = a35.split(",")
a40_sp = a40.split(",")
a45_sp = a45.split(",")
a50_sp = a50.split(",")
a55_sp = a55.split(",")
a60_sp = a60.split(",")
a65_sp = a65.split(",")
```

```
a70_sp = a70.split(",")
a75_sp = a75.split(",")
a80_sp = a80.split(",")
a85_sp = a85.split(",")
a90_sp = a90.split(",")
a95_sp = a95.split(",")
a100_sp = a100.split(",")


# qx
a0_qx = float(a0_sp[1])
a1_qx = float(a1_sp[1])
a5_qx = float(a5_sp[1])
a10_qx = float(a10_sp[1])
a15_qx = float(a15_sp[1])
a20_qx = float(a20_sp[1])
a25_qx = float(a25_sp[1])
a30_qx = float(a30_sp[1])
a35_qx = float(a35_sp[1])
a40_qx = float(a40_sp[1])
a45_qx = float(a45_sp[1])
a50_qx = float(a50_sp[1])
a55_qx = float(a55_sp[1])
a60_qx = float(a60_sp[1])
a65_qx = float(a65_sp[1])
a70_qx = float(a70_sp[1])
a75_qx = float(a75_sp[1])
a80_qx = float(a80_sp[1])
a85_qx = float(a85_sp[1])
a90_qx = float(a90_sp[1])
a95_qx = float(a95_sp[1])
a100_qx = float(a100_sp[1])


# qx - lower bound
a0_qxl = float(a0_sp[2])
a1_qxl = float(a1_sp[2])
a5_qxl = float(a5_sp[2])
a10_qxl = float(a10_sp[2])
a15_qxl = float(a15_sp[2])
a20_qxl = float(a20_sp[2])
a25_qxl = float(a25_sp[2])
a30_qxl = float(a30_sp[2])
a35_qxl = float(a35_sp[2])
a40_qxl = float(a40_sp[2])
a45_qxl = float(a45_sp[2])
a50_qxl = float(a50_sp[2])
a55_qxl = float(a55_sp[2])
a60_qxl = float(a60_sp[2])
a65_qxl = float(a65_sp[2])
a70_qxl = float(a70_sp[2])
a75_qxl = float(a75_sp[2])
a80_qxl = float(a80_sp[2])
a85_qxl = float(a85_sp[2])
a90_qxl = float(a90_sp[2])
a95_qxl = float(a95_sp[2])
a100_qxl = float(a100_sp[2])

# qx - Upper bound
a0_qxu = float(a0_sp[3])
```

```
a1_qxu = float(a1_sp[3])
a5_qxu = float(a5_sp[3])
a10_qxu = float(a10_sp[3])
a15_qxu = float(a15_sp[3])
a20_qxu = float(a20_sp[3])
a25_qxu = float(a25_sp[3])
a30_qxu = float(a30_sp[3])
a35_qxu = float(a35_sp[3])
a40_qxu = float(a40_sp[3])
a45_qxu = float(a45_sp[3])
a50_qxu = float(a50_sp[3])
a55_qxu = float(a55_sp[3])
a60_qxu = float(a60_sp[3])
a65_qxu = float(a65_sp[3])
a70_qxu = float(a70_sp[3])
a75_qxu = float(a75_sp[3])
a80_qxu = float(a80_sp[3])
a85_qxu = float(a85_sp[3])
a90_qxu = float(a90_sp[3])
a95_qxu = float(a95_sp[3])
a100_qxu = float(a100_sp[3])


# ax
a0_ax = float(a0_sp[4])
a1_ax = float(a1_sp[4])
a5_ax = float(a5_sp[4])
a10_ax = float(a10_sp[4])
a15_ax = float(a15_sp[4])
a20_ax = float(a20_sp[4])
a25_ax = float(a25_sp[4])
a30_ax = float(a30_sp[4])
a35_ax = float(a35_sp[4])
a40_ax = float(a40_sp[4])
a45_ax = float(a45_sp[4])
a50_ax = float(a50_sp[4])
a55_ax = float(a55_sp[4])
a60_ax = float(a60_sp[4])
a65_ax = float(a65_sp[4])
a70_ax = float(a70_sp[4])
a75_ax = float(a75_sp[4])
a80_ax = float(a80_sp[4])
a85_ax = float(a85_sp[4])
a90_ax = float(a90_sp[4])
a95_ax = float(a95_sp[4])
a100_ax = float(a100_sp[4])

# ax - LB
a0_axl = float(a0_sp[5])
a1_axl = float(a1_sp[5])
a5_axl = float(a5_sp[5])
a10_axl = float(a10_sp[5])
a15_axl = float(a15_sp[5])
a20_axl = float(a20_sp[5])
a25_axl = float(a25_sp[5])
a30_axl = float(a30_sp[5])
a35_axl = float(a35_sp[5])
a40_axl = float(a40_sp[5])
a45_axl = float(a45_sp[5])
a50_axl = float(a50_sp[5])
```

```
a55_axl = float(a55_sp[5])
a60_axl = float(a60_sp[5])
a65_axl = float(a65_sp[5])
a70_axl = float(a70_sp[5])
a75_axl = float(a75_sp[5])
a80_axl = float(a80_sp[5])
a85_axl = float(a85_sp[5])
a90_axl = float(a90_sp[5])
a95_axl = float(a95_sp[5])
a100_axl = float(a100_sp[5])


# ax - UB
a0_axu = float(a0_sp[6])
a1_axu = float(a1_sp[6])
a5_axu = float(a5_sp[6])
a10_axu = float(a10_sp[6])
a15_axu = float(a15_sp[6])
a20_axu = float(a20_sp[6])
a25_axu = float(a25_sp[6])
a30_axu = float(a30_sp[6])
a35_axu = float(a35_sp[6])
a40_axu = float(a40_sp[6])
a45_axu = float(a45_sp[6])
a50_axu = float(a50_sp[6])
a55_axu = float(a55_sp[6])
a60_axu = float(a60_sp[6])
a65_axu = float(a65_sp[6])
a70_axu = float(a70_sp[6])
a75_axu = float(a75_sp[6])
a80_axu = float(a80_sp[6])
a85_axu = float(a85_sp[6])
a90_axu = float(a90_sp[6])
a95_axu = float(a95_sp[6])
a100_axu = float(a100_sp[6])


# 2019 mx - 2019 LT
a0_m19 = float(a0_sp[7])
a1_m19 = float(a1_sp[7])
a5_m19 = float(a5_sp[7])
a10_m19 = float(a10_sp[7])
a15_m19 = float(a15_sp[7])
a20_m19 = float(a20_sp[7])
a25_m19 = float(a25_sp[7])
a30_m19 = float(a30_sp[7])
a35_m19 = float(a35_sp[7])
a40_m19 = float(a40_sp[7])
a45_m19 = float(a45_sp[7])
a50_m19 = float(a50_sp[7])
a55_m19 = float(a55_sp[7])
a60_m19 = float(a60_sp[7])
a65_m19 = float(a65_sp[7])
a70_m19 = float(a70_sp[7])
a75_m19 = float(a75_sp[7])
a80_m19 = float(a80_sp[7])
a85_m19 = float(a85_sp[7])
a90_m19 = float(a90_sp[7])
a95_m19 = float(a95_sp[7])
a100_m19 = float(a100_sp[7])
```

```
count = 0
while count < 50000: #5000: #50000

# Random qx by age - need to check we are not getting any values below zero

#    make this uniform
    a0_rand_qx = random.uniform(a0_qxl,a0_qxu)
    a1_rand_qx = random.uniform(a1_qxl,a1_qxu)
    a5_rand_qx = random.uniform(a5_qxl,a5_qxu)
    a10_rand_qx = random.uniform(a10_qxl,a10_qxu)
    a15_rand_qx = random.uniform(a15_qxl,a15_qxu)
    a20_rand_qx = random.uniform(a20_qxl,a20_qxu)
    a25_rand_qx = random.uniform(a25_qxl,a25_qxu)
    a30_rand_qx = random.uniform(a30_qxl,a30_qxu)
    a35_rand_qx = random.uniform(a35_qxl,a35_qxu)
    a40_rand_qx = random.uniform(a40_qxl,a40_qxu)
    a45_rand_qx = random.uniform(a45_qxl,a45_qxu)
    a50_rand_qx = random.uniform(a50_qxl,a50_qxu)
    a55_rand_qx = random.uniform(a55_qxl,a55_qxu)
    a60_rand_qx = random.uniform(a60_qxl,a60_qxu)
    a65_rand_qx = random.uniform(a65_qxl,a65_qxu)
    a70_rand_qx = random.uniform(a70_qxl,a70_qxu)
    a75_rand_qx = random.uniform(a75_qxl,a75_qxu)
    a80_rand_qx = random.uniform(a80_qxl,a80_qxu)
    a85_rand_qx = random.uniform(a85_qxl,a85_qxu)
    a90_rand_qx = random.uniform(a90_qxl,a90_qxu)
    a95_rand_qx = random.uniform(a95_qxl,a95_qxu)
    a100_rand_qx =  1

# Random ax by age - need to check we are not getting any values below zero

#    make this uniform
    a0_rand_ax = random.uniform(a0_axl,a0_axu)
    a1_rand_ax = random.uniform(a1_axl,a1_axu)
    a5_rand_ax = random.uniform(a5_axl,a5_axu)
    a10_rand_ax = random.uniform(a10_axl,a10_axu)
    a15_rand_ax = random.uniform(a15_axl,a15_axu)
    a20_rand_ax = random.uniform(a20_axl,a20_axu)
    a25_rand_ax = random.uniform(a25_axl,a25_axu)
    a30_rand_ax = random.uniform(a30_axl,a30_axu)
    a35_rand_ax = random.uniform(a35_axl,a35_axu)
    a40_rand_ax = random.uniform(a40_axl,a40_axu)
    a45_rand_ax = random.uniform(a45_axl,a45_axu)
    a50_rand_ax = random.uniform(a50_axl,a50_axu)
    a55_rand_ax = random.uniform(a55_axl,a55_axu)
    a60_rand_ax = random.uniform(a60_axl,a60_axu)
    a65_rand_ax = random.uniform(a65_axl,a65_axu)
    a70_rand_ax = random.uniform(a70_axl,a70_axu)
    a75_rand_ax = random.uniform(a75_axl,a75_axu)
    a80_rand_ax = random.uniform(a80_axl,a80_axu)
    a85_rand_ax = random.uniform(a85_axl,a85_axu)
    a90_rand_ax = random.uniform(a90_axl,a90_axu)
    a95_rand_ax = random.uniform(a95_axl,a95_axu)
    a100_rand_ax = random.uniform(a100_axl,a100_axu)
```

```
### calculate life table variables
    radix = 1000000.0000000

    # calculate the number of deaths age0
    a0_dx = a0_rand_qx*radix
    # calculate survivors
    a0_lx=radix
    a0_sx=a0_lx/radix # this is 1?
    a1_lx=(radix-a0_dx)
    a1_sx = a1_lx/radix

    # calculate the number of deaths age1
    a1_dx = a1_rand_qx*a1_lx
    # calculate survivors
    a5_lx=(a1_lx-a1_dx)
    a5_sx = a5_lx/radix

    # calculate the number of deaths age5
    a5_dx = a5_rand_qx*a5_lx
    # calculate survivors
    a10_lx=(a5_lx-a5_dx)
    a10_sx = a10_lx/radix

    # calculate the number of deaths age10
    a10_dx = a10_rand_qx*a10_lx
    # calculate survivors
    a15_lx=(a10_lx-a10_dx)
    a15_sx = a15_lx/radix

    # calculate the number of deaths age15
    a15_dx = a15_rand_qx*a15_lx
    # calculate survivors
    a20_lx=(a15_lx-a15_dx)
    a20_sx = a20_lx/radix

    # calculate the number of deaths age20
    a20_dx = a20_rand_qx*a20_lx
    # calculate survivors
    a25_lx=(a20_lx-a20_dx)
    a25_sx = a25_lx/radix

    # calculate the number of deaths age25
    a25_dx = a25_rand_qx*a25_lx
    # calculate survivors
    a30_lx=(a25_lx-a25_dx)
    a30_sx = a30_lx/radix

    # calculate the number of deaths age30
    a30_dx = a30_rand_qx*a30_lx
    # calculate survivors
    a35_lx=(a30_lx-a30_dx)
    a35_sx = a35_lx/radix

    # calculate the number of deaths age35
    a35_dx = a35_rand_qx*a35_lx
    # calculate survivors
    a40_lx=(a35_lx-a35_dx)
```

```
a40_sx = a40_lx/radix

# calculate the number of deaths age40
a40_dx = a40_rand_qx*a40_lx
# calculate survivors
a45_lx=(a40_lx-a40_dx)
a45_sx = a45_lx/radix

# calculate the number of deaths age45
a45_dx = a45_rand_qx*a45_lx
# calculate survivors
a50_lx=(a45_lx-a45_dx)
a50_sx = a50_lx/radix

# calculate the number of deaths age50
a50_dx = a50_rand_qx*a50_lx
# calculate survivors
a55_lx=(a50_lx-a50_dx)
a55_sx = a55_lx/radix

# calculate the number of deaths age55
a55_dx = a55_rand_qx*a55_lx
# calculate survivors
a60_lx=(a55_lx-a55_dx)
a60_sx = a60_lx/radix

# calculate the number of deaths age60
a60_dx = a60_rand_qx*a60_lx
# calculate survivors
a65_lx=(a60_lx-a60_dx)
a65_sx = a65_lx/radix

# calculate the number of deaths age65
a65_dx = a65_rand_qx*a65_lx
# calculate survivors
a70_lx=(a65_lx-a65_dx)
a70_sx = a70_lx/radix

# calculate the number of deaths age70
a70_dx = a70_rand_qx*a70_lx
# calculate survivors
a75_lx=(a70_lx-a70_dx)
a75_sx = a75_lx/radix

# calculate the number of deaths age75
a75_dx = a75_rand_qx*a75_lx
# calculate survivors
a80_lx=(a75_lx-a75_dx)
a80_sx = a80_lx/radix

# calculate the number of deaths age80
a80_dx = a80_rand_qx*a80_lx
# calculate survivors
a85_lx=(a80_lx-a80_dx)
a85_sx = a85_lx/radix

# calculate the number of deaths age85
a85_dx = a85_rand_qx*a85_lx
# calculate survivors
a90_lx=(a85_lx-a85_dx)
```

```
    a90_sx = a90_lx/radix

    # calculate the number of deaths age90
    a90_dx = a90_rand_qx*a90_lx
    # calculate survivors
    a95_lx=(a90_lx-a90_dx)
    a95_sx = a95_lx/radix

    # calculate the number of deaths age95
    a95_dx = a95_rand_qx*a95_lx
    # calculate survivors
    a100_lx=(a95_lx-a95_dx)
    a100_sx = a100_lx/radix

    # calculate the number of deaths age100
    a100_dx = a100_rand_qx*a100_lx
    # No Survivors - top-coded

    #calculate Lx
    a0_Lx = (a1_lx*1)+(a0_dx*a0_rand_ax)
    a1_Lx = (a5_lx*4)+(a1_dx*a1_rand_ax)
    a5_Lx = (a10_lx*5)+(a5_dx*a5_rand_ax)
    a10_Lx = (a15_lx*5)+(a10_dx*a10_rand_ax)
    a15_Lx = (a20_lx*5)+(a15_dx*a15_rand_ax)
    a20_Lx = (a25_lx*5)+(a20_dx*a20_rand_ax)
    a25_Lx = (a30_lx*5)+(a25_dx*a25_rand_ax)
    a30_Lx = (a35_lx*5)+(a30_dx*a30_rand_ax)
    a35_Lx = (a40_lx*5)+(a35_dx*a35_rand_ax)
    a40_Lx = (a45_lx*5)+(a40_dx*a40_rand_ax)
    a45_Lx = (a50_lx*5)+(a45_dx*a45_rand_ax)
    a50_Lx = (a55_lx*5)+(a50_dx*a50_rand_ax)
    a55_Lx = (a60_lx*5)+(a55_dx*a55_rand_ax)
    a60_Lx = (a65_lx*5)+(a60_dx*a60_rand_ax)
    a65_Lx = (a70_lx*5)+(a65_dx*a65_rand_ax)
    a70_Lx = (a75_lx*5)+(a70_dx*a70_rand_ax)
    a75_Lx = (a80_lx*5)+(a75_dx*a75_rand_ax)
    a80_Lx = (a85_lx*5)+(a80_dx*a80_rand_ax)
    a85_Lx = (a90_lx*5)+(a85_dx*a85_rand_ax)
    a90_Lx = (a95_lx*5)+(a90_dx*a90_rand_ax)
    a95_Lx = (a100_lx*5)+(a95_dx*a95_rand_ax)
    a100_Lx = (a100_dx*a100_rand_ax)


    ####
    # calculate Tx
    a0_Tx =
a0_Lx+a1_Lx+a5_Lx+a10_Lx+a15_Lx+a20_Lx+a25_Lx+a30_Lx+a35_Lx+a40_Lx+a45_Lx+a50_Lx+a55_Lx+a60_Lx+a65_Lx+a70_L
x+a75_Lx+a80_Lx+a85_Lx+a90_Lx+a95_Lx+a100_Lx
    a1_Tx =
a1_Lx+a5_Lx+a10_Lx+a15_Lx+a20_Lx+a25_Lx+a30_Lx+a35_Lx+a40_Lx+a45_Lx+a50_Lx+a55_Lx+a60_Lx+a65_Lx+a70_Lx+a75_
Lx+a80_Lx+a85_Lx+a90_Lx+a95_Lx+a100_Lx
    a5_Tx =
a5_Lx+a10_Lx+a15_Lx+a20_Lx+a25_Lx+a30_Lx+a35_Lx+a40_Lx+a45_Lx+a50_Lx+a55_Lx+a60_Lx+a65_Lx+a70_Lx+a75_Lx+a80
_Lx+a85_Lx+a90_Lx+a95_Lx+a100_Lx
    a10_Tx =
a10_Lx+a15_Lx+a20_Lx+a25_Lx+a30_Lx+a35_Lx+a40_Lx+a45_Lx+a50_Lx+a55_Lx+a60_Lx+a65_Lx+a70_Lx+a75_Lx+a80_Lx+a8
5_Lx+a90_Lx+a95_Lx+a100_Lx
    a15_Tx =
a15_Lx+a20_Lx+a25_Lx+a30_Lx+a35_Lx+a40_Lx+a45_Lx+a50_Lx+a55_Lx+a60_Lx+a65_Lx+a70_Lx+a75_Lx+a80_Lx+a85_Lx+a9
0_Lx+a95_Lx+a100_Lx
```

```
    a20_Tx =
a20_Lx+a25_Lx+a30_Lx+a35_Lx+a40_Lx+a45_Lx+a50_Lx+a55_Lx+a60_Lx+a65_Lx+a70_Lx+a75_Lx+a80_Lx+a85_Lx+a90_Lx+a9
5_Lx+a100_Lx
    a25_Tx =
a25_Lx+a30_Lx+a35_Lx+a40_Lx+a45_Lx+a50_Lx+a55_Lx+a60_Lx+a65_Lx+a70_Lx+a75_Lx+a80_Lx+a85_Lx+a90_Lx+a95_Lx+a1
00_Lx
    a30_Tx =
a30_Lx+a35_Lx+a40_Lx+a45_Lx+a50_Lx+a55_Lx+a60_Lx+a65_Lx+a70_Lx+a75_Lx+a80_Lx+a85_Lx+a90_Lx+a95_Lx+a100_Lx
    a35_Tx =
a35_Lx+a40_Lx+a45_Lx+a50_Lx+a55_Lx+a60_Lx+a65_Lx+a70_Lx+a75_Lx+a80_Lx+a85_Lx+a90_Lx+a95_Lx+a100_Lx
    a40_Tx = a40_Lx+a45_Lx+a50_Lx+a55_Lx+a60_Lx+a65_Lx+a70_Lx+a75_Lx+a80_Lx+a85_Lx+a90_Lx+a95_Lx+a100_Lx
    a45_Tx = a45_Lx+a50_Lx+a55_Lx+a60_Lx+a65_Lx+a70_Lx+a75_Lx+a80_Lx+a85_Lx+a90_Lx+a95_Lx+a100_Lx
    a50_Tx = a50_Lx+a55_Lx+a60_Lx+a65_Lx+a70_Lx+a75_Lx+a80_Lx+a85_Lx+a90_Lx+a95_Lx+a100_Lx
    a55_Tx = a55_Lx+a60_Lx+a65_Lx+a70_Lx+a75_Lx+a80_Lx+a85_Lx+a90_Lx+a95_Lx+a100_Lx
    a60_Tx = a60_Lx+a65_Lx+a70_Lx+a75_Lx+a80_Lx+a85_Lx+a90_Lx+a95_Lx+a100_Lx
    a65_Tx = a65_Lx+a70_Lx+a75_Lx+a80_Lx+a85_Lx+a90_Lx+a95_Lx+a100_Lx
    a70_Tx = a70_Lx+a75_Lx+a80_Lx+a85_Lx+a90_Lx+a95_Lx+a100_Lx
    a75_Tx = a75_Lx+a80_Lx+a85_Lx+a90_Lx+a95_Lx+a100_Lx
    a80_Tx = a80_Lx+a85_Lx+a90_Lx+a95_Lx+a100_Lx
    a85_Tx = a85_Lx+a90_Lx+a95_Lx+a100_Lx
    a90_Tx = a90_Lx+a95_Lx+a100_Lx
    a95_Tx = a95_Lx+a100_Lx
    a100_Tx = a100_Lx



#calculate mx
    a0_mx = (a0_dx)/a0_Lx
    a1_mx = (a1_dx)/a1_Lx
    a5_mx = (a5_dx)/a5_Lx
    a10_mx = (a10_dx)/a10_Lx
    a15_mx = (a15_dx)/a15_Lx
    a20_mx = (a20_dx)/a20_Lx
    a25_mx = (a25_dx)/a25_Lx
    a30_mx = (a30_dx)/a30_Lx
    a35_mx = (a35_dx)/a35_Lx
    a40_mx = (a40_dx)/a40_Lx
    a45_mx = (a45_dx)/a45_Lx
    a50_mx = (a50_dx)/a50_Lx
    a55_mx = (a55_dx)/a55_Lx
    a60_mx = (a60_dx)/a60_Lx
    a65_mx = (a65_dx)/a65_Lx
    a70_mx = (a70_dx)/a70_Lx
    a75_mx = (a75_dx)/a75_Lx
    a80_mx = (a80_dx)/a80_Lx
    a85_mx = (a85_dx)/a85_Lx
    a90_mx = (a90_dx)/a90_Lx
    a95_mx = (a95_dx)/a95_Lx
    a100_mx = (a100_dx)/a100_Lx



    #calculate Lx 0-14, 15-39, 40-64, 65-84, 85+

    a015_tx = (a0_Lx+a1_Lx+a5_Lx+a10_Lx)
    a1539_tx = (a15_Lx+a20_Lx+a25_Lx+a30_Lx+a35_Lx)
    a4064_tx = (a40_Lx+a45_Lx+a50_Lx+a55_Lx+a60_Lx)
    a6584_tx = (a65_Lx+a70_Lx+a75_Lx+a80_Lx)
    a85_tx = a85_Tx
```

```
# simulation of weighted-averages of 2020:2019 mx ratio:0-15, 15-40, 40-65, 65-85, 85+

    a015_rr  =
(a0_mx/a0_m19)*(a0_Lx/a015_tx)+(a1_mx/a1_m19)*(a1_Lx/a015_tx)+(a5_mx/a5_m19)*(a5_Lx/a015_tx)+(a10_mx/a10_m19)*(a10_
Lx/a015_tx)
    a1539_rr =
(a15_mx/a15_m19)*(a15_Lx/a1539_tx)+(a20_mx/a20_m19)*(a20_Lx/a1539_tx)+(a25_mx/a25_m19)*(a25_Lx/a1539_tx)+(a30_mx/a
30_m19)*(a30_Lx/a1539_tx)+(a35_mx/a35_m19)*(a35_Lx/a1539_tx)
    a4064_rr =
(a40_mx/a40_m19)*(a40_Lx/a4064_tx)+(a45_mx/a45_m19)*(a45_Lx/a4064_tx)+(a50_mx/a50_m19)*(a50_Lx/a4064_tx)+(a55_mx/a
55_m19)*(a55_Lx/a4064_tx)+(a60_mx/a60_m19)*(a60_Lx/a4064_tx)
    a6584_rr =
(a65_mx/a65_m19)*(a65_Lx/a6584_tx)+(a70_mx/a70_m19)*(a70_Lx/a6584_tx)+(a75_mx/a75_m19)*(a75_Lx/a6584_tx)+(a80_mx/a
80_m19)*(a80_Lx/a6584_tx)
    a85110_rr =
(a85_mx/a85_m19)*(a85_Lx/a85_tx)+(a90_mx/a90_m19)*(a90_Lx/a85_tx)+(a95_mx/a95_m19)*(a95_Lx/a85_tx)+(a100_mx/a100_m
19)*(a100_Lx/a85_tx)


# this outputs the mortality rates for each sim
    mxgrp = r"/Users/ryan/Documents/Papers/LE Trends_COVID/2021 Life Expectancy/simulations/peer/male/deu_m_mxgrp.txt"
    opened_file = open(mxgrp, 'a')

    if count==0:
        opened_file.write('{0} {1} {2} {3} {4} {5}\n'.format("sim_num","rr015","rr1539","rr4064","rr6584","rr85"))

    else:
        opened_file.write('{0} {1} {2} {3} {4} {5}\n'.format(count,a015_rr,a1539_rr,a4064_rr,a6584_rr,a85110_rr))


    ###### estimate life expectancy

    #calculate ex
    a0_ex = a0_Tx/radix
    a1_ex = a1_Tx/a1_lx
    a5_ex = a5_Tx/a5_lx
    a10_ex = a10_Tx/a10_lx
    a15_ex = a15_Tx/a15_lx
    a20_ex = a20_Tx/a20_lx
    a25_ex = a25_Tx/a25_lx
    a30_ex = a30_Tx/a30_lx
    a35_ex = a35_Tx/a35_lx
    a40_ex = a40_Tx/a40_lx
    a45_ex = a45_Tx/a45_lx
    a50_ex = a50_Tx/a50_lx
    a55_ex = a55_Tx/a55_lx
    a60_ex = a60_Tx/a60_lx
    a65_ex = a65_Tx/a65_lx
    a70_ex = a70_Tx/a70_lx
    a75_ex = a75_Tx/a75_lx
    a80_ex = a80_Tx/a80_lx
    a85_ex = a85_Tx/a85_lx
    a90_ex = a90_Tx/a90_lx
    a95_ex = a95_Tx/a95_lx
    a100_ex = a100_Tx/a100_lx




    # save data
```

```
    tot_file_name = r"/Users/ryan/Documents/Papers/LE Trends_COVID/2021 Life Expectancy/simulations/peer/male/deu_m_ex.txt"

    tot_opened_file = open(tot_file_name, 'a')

    if count==0:
        tot_opened_file.write('{0} {1} {2} {3}\n'.format("sim_num","age","sx", "ex"))
        tot_opened_file.write('{0} {1} {2} {3}\n'.format(count,"0",a0_sx,a0_ex))
        tot_opened_file.write('{0} {1} {2} {3}\n'.format(count,"1",a1_sx,a1_ex))
        tot_opened_file.write('{0} {1} {2} {3}\n'.format(count,"5",a5_sx,a5_ex))
        tot_opened_file.write('{0} {1} {2} {3}\n'.format(count,"10",a10_sx,a10_ex))
        tot_opened_file.write('{0} {1} {2} {3}\n'.format(count,"15",a15_sx,a15_ex))
        tot_opened_file.write('{0} {1} {2} {3}\n'.format(count,"20",a20_sx,a20_ex))
        tot_opened_file.write('{0} {1} {2} {3}\n'.format(count,"25",a25_sx,a25_ex))
        tot_opened_file.write('{0} {1} {2} {3}\n'.format(count,"30",a30_sx,a30_ex))
        tot_opened_file.write('{0} {1} {2} {3}\n'.format(count,"35",a35_sx,a35_ex))
        tot_opened_file.write('{0} {1} {2} {3}\n'.format(count,"40",a40_sx,a40_ex))
        tot_opened_file.write('{0} {1} {2} {3}\n'.format(count,"45",a45_sx,a45_ex))
        tot_opened_file.write('{0} {1} {2} {3}\n'.format(count,"50",a50_sx,a50_ex))
        tot_opened_file.write('{0} {1} {2} {3}\n'.format(count,"55",a55_sx,a55_ex))
        tot_opened_file.write('{0} {1} {2} {3}\n'.format(count,"60",a60_sx,a60_ex))
        tot_opened_file.write('{0} {1} {2} {3}\n'.format(count,"65",a65_sx,a65_ex))
        tot_opened_file.write('{0} {1} {2} {3}\n'.format(count,"70",a70_sx,a70_ex))
        tot_opened_file.write('{0} {1} {2} {3}\n'.format(count,"75",a75_sx,a75_ex))
        tot_opened_file.write('{0} {1} {2} {3}\n'.format(count,"80",a80_sx,a80_ex))
        tot_opened_file.write('{0} {1} {2} {3}\n'.format(count,"85",a85_sx,a85_ex))
        tot_opened_file.write('{0} {1} {2} {3}\n'.format(count,"90",a90_sx,a90_ex))
        tot_opened_file.write('{0} {1} {2} {3}\n'.format(count,"95",a95_sx,a95_ex))
        tot_opened_file.write('{0} {1} {2} {3}\n'.format(count,"100",a100_sx,a100_ex))


    else:
        tot_opened_file.write('{0} {1} {2} {3}\n'.format(count,"0",a0_sx,a0_ex))
        tot_opened_file.write('{0} {1} {2} {3}\n'.format(count,"1",a1_sx,a1_ex))
        tot_opened_file.write('{0} {1} {2} {3}\n'.format(count,"5",a5_sx,a5_ex))
        tot_opened_file.write('{0} {1} {2} {3}\n'.format(count,"10",a10_sx,a10_ex))
        tot_opened_file.write('{0} {1} {2} {3}\n'.format(count,"15",a15_sx,a15_ex))
        tot_opened_file.write('{0} {1} {2} {3}\n'.format(count,"20",a20_sx,a20_ex))
        tot_opened_file.write('{0} {1} {2} {3}\n'.format(count,"25",a25_sx,a25_ex))
        tot_opened_file.write('{0} {1} {2} {3}\n'.format(count,"30",a30_sx,a30_ex))
        tot_opened_file.write('{0} {1} {2} {3}\n'.format(count,"35",a35_sx,a35_ex))
        tot_opened_file.write('{0} {1} {2} {3}\n'.format(count,"40",a40_sx,a40_ex))
        tot_opened_file.write('{0} {1} {2} {3}\n'.format(count,"45",a45_sx,a45_ex))
        tot_opened_file.write('{0} {1} {2} {3}\n'.format(count,"50",a50_sx,a50_ex))
        tot_opened_file.write('{0} {1} {2} {3}\n'.format(count,"55",a55_sx,a55_ex))
        tot_opened_file.write('{0} {1} {2} {3}\n'.format(count,"60",a60_sx,a60_ex))
        tot_opened_file.write('{0} {1} {2} {3}\n'.format(count,"65",a65_sx,a65_ex))
        tot_opened_file.write('{0} {1} {2} {3}\n'.format(count,"70",a70_sx,a70_ex))
        tot_opened_file.write('{0} {1} {2} {3}\n'.format(count,"75",a75_sx,a75_ex))
        tot_opened_file.write('{0} {1} {2} {3}\n'.format(count,"80",a80_sx,a80_ex))
        tot_opened_file.write('{0} {1} {2} {3}\n'.format(count,"85",a85_sx,a85_ex))
        tot_opened_file.write('{0} {1} {2} {3}\n'.format(count,"90",a90_sx,a90_ex))
        tot_opened_file.write('{0} {1} {2} {3}\n'.format(count,"95",a95_sx,a95_ex))
        tot_opened_file.write('{0} {1} {2} {3}\n'.format(count,"100",a100_sx,a100_ex))


    print(count)
    count += 1  # This is the same as count = count + 1

tot_opened_file.close()
```

```
print("simulation completed")
```

### Spain Total Population, 2021

```
# -*- coding: utf-8 -*-
"""
Created 5/25

Peer 2021 Life Tables from 2018/2019/2020 qx*RR

@author: ryan masters


"""
#import packages
import random

# importing in the qx and error and ax
esp_t = r"/Users/ryan/Documents/Papers/LE Trends_COVID/2021 Life Expectancy/simulations/peer/total/esp_t.txt"
# change as needed for input files

# read in the file
textFile = open(esp_t,'r')
text = textFile.readlines()
# split into different age categories
a0=text[1]
a1=text[2]
a5=text[3]
a10=text[4]
a15=text[5]
a20=text[6]
a25=text[7]
a30=text[8]
a35=text[9]
a40=text[10]
a45=text[11]
a50=text[12]
a55=text[13]
a60=text[14]
a65=text[15]
a70=text[16]
a75=text[17]
a80=text[18]
a85=text[19]
a90=text[20]
a95=text[21]
a100=text[22]
a105=text[23]
a110=text[24]


a0_sp = a0.split(",")
a1_sp = a1.split(",")
a5_sp = a5.split(",")
a10_sp = a10.split(",")
a15_sp = a15.split(",")
```

```
a20_sp = a20.split(",")
a25_sp = a25.split(",")
a30_sp = a30.split(",")
a35_sp = a35.split(",")
a40_sp = a40.split(",")
a45_sp = a45.split(",")
a50_sp = a50.split(",")
a55_sp = a55.split(",")
a60_sp = a60.split(",")
a65_sp = a65.split(",")
a70_sp = a70.split(",")
a75_sp = a75.split(",")
a80_sp = a80.split(",")
a85_sp = a85.split(",")
a90_sp = a90.split(",")
a95_sp = a95.split(",")
a100_sp = a100.split(",")
a105_sp = a105.split(",")
a110_sp = a110.split(",")

# qx
a0_qx = float(a0_sp[1])
a1_qx = float(a1_sp[1])
a5_qx = float(a5_sp[1])
a10_qx = float(a10_sp[1])
a15_qx = float(a15_sp[1])
a20_qx = float(a20_sp[1])
a25_qx = float(a25_sp[1])
a30_qx = float(a30_sp[1])
a35_qx = float(a35_sp[1])
a40_qx = float(a40_sp[1])
a45_qx = float(a45_sp[1])
a50_qx = float(a50_sp[1])
a55_qx = float(a55_sp[1])
a60_qx = float(a60_sp[1])
a65_qx = float(a65_sp[1])
a70_qx = float(a70_sp[1])
a75_qx = float(a75_sp[1])
a80_qx = float(a80_sp[1])
a85_qx = float(a85_sp[1])
a90_qx = float(a90_sp[1])
a95_qx = float(a95_sp[1])
a100_qx = float(a100_sp[1])
a105_qx = float(a105_sp[1])
a110_qx = float(a110_sp[1])

# qx - lower bound
a0_qxl = float(a0_sp[2])
a1_qxl = float(a1_sp[2])
a5_qxl = float(a5_sp[2])
a10_qxl = float(a10_sp[2])
a15_qxl = float(a15_sp[2])
a20_qxl = float(a20_sp[2])
a25_qxl = float(a25_sp[2])
a30_qxl = float(a30_sp[2])
a35_qxl = float(a35_sp[2])
a40_qxl = float(a40_sp[2])
a45_qxl = float(a45_sp[2])
a50_qxl = float(a50_sp[2])
a55_qxl = float(a55_sp[2])
```

```
a60_qxl = float(a60_sp[2])
a65_qxl = float(a65_sp[2])
a70_qxl = float(a70_sp[2])
a75_qxl = float(a75_sp[2])
a80_qxl = float(a80_sp[2])
a85_qxl = float(a85_sp[2])
a90_qxl = float(a90_sp[2])
a95_qxl = float(a95_sp[2])
a100_qxl = float(a100_sp[2])
a105_qxl = float(a105_sp[2])
a110_qxl = float(a110_sp[2])

# qx - upper bound
a0_qxu = float(a0_sp[3])
a1_qxu = float(a1_sp[3])
a5_qxu = float(a5_sp[3])
a10_qxu = float(a10_sp[3])
a15_qxu = float(a15_sp[3])
a20_qxu = float(a20_sp[3])
a25_qxu = float(a25_sp[3])
a30_qxu = float(a30_sp[3])
a35_qxu = float(a35_sp[3])
a40_qxu = float(a40_sp[3])
a45_qxu = float(a45_sp[3])
a50_qxu = float(a50_sp[3])
a55_qxu = float(a55_sp[3])
a60_qxu = float(a60_sp[3])
a65_qxu = float(a65_sp[3])
a70_qxu = float(a70_sp[3])
a75_qxu = float(a75_sp[3])
a80_qxu = float(a80_sp[3])
a85_qxu = float(a85_sp[3])
a90_qxu = float(a90_sp[3])
a95_qxu = float(a95_sp[3])
a100_qxu = float(a100_sp[3])
a105_qxu = float(a105_sp[3])
a110_qxu = float(a110_sp[3])

# ax
a0_ax = float(a0_sp[4])
a1_ax = float(a1_sp[4])
a5_ax = float(a5_sp[4])
a10_ax = float(a10_sp[4])
a15_ax = float(a15_sp[4])
a20_ax = float(a20_sp[4])
a25_ax = float(a25_sp[4])
a30_ax = float(a30_sp[4])
a35_ax = float(a35_sp[4])
a40_ax = float(a40_sp[4])
a45_ax = float(a45_sp[4])
a50_ax = float(a50_sp[4])
a55_ax = float(a55_sp[4])
a60_ax = float(a60_sp[4])
a65_ax = float(a65_sp[4])
a70_ax = float(a70_sp[4])
a75_ax = float(a75_sp[4])
a80_ax = float(a80_sp[4])
a85_ax = float(a85_sp[4])
a90_ax = float(a90_sp[4])
a95_ax = float(a95_sp[4])
```

```
a100_ax = float(a100_sp[4])
a105_ax = float(a105_sp[4])
a110_ax = float(a110_sp[4])

# ax - LB
a0_axl = float(a0_sp[5])
a1_axl = float(a1_sp[5])
a5_axl = float(a5_sp[5])
a10_axl = float(a10_sp[5])
a15_axl = float(a15_sp[5])
a20_axl = float(a20_sp[5])
a25_axl = float(a25_sp[5])
a30_axl = float(a30_sp[5])
a35_axl = float(a35_sp[5])
a40_axl = float(a40_sp[5])
a45_axl = float(a45_sp[5])
a50_axl = float(a50_sp[5])
a55_axl = float(a55_sp[5])
a60_axl = float(a60_sp[5])
a65_axl = float(a65_sp[5])
a70_axl = float(a70_sp[5])
a75_axl = float(a75_sp[5])
a80_axl = float(a80_sp[5])
a85_axl = float(a85_sp[5])
a90_axl = float(a90_sp[5])
a95_axl = float(a95_sp[5])
a100_axl = float(a100_sp[5])
a105_axl = float(a105_sp[5])
a110_axl = float(a110_sp[5])

# ax - UB
a0_axu = float(a0_sp[6])
a1_axu = float(a1_sp[6])
a5_axu = float(a5_sp[6])
a10_axu = float(a10_sp[6])
a15_axu = float(a15_sp[6])
a20_axu = float(a20_sp[6])
a25_axu = float(a25_sp[6])
a30_axu = float(a30_sp[6])
a35_axu = float(a35_sp[6])
a40_axu = float(a40_sp[6])
a45_axu = float(a45_sp[6])
a50_axu = float(a50_sp[6])
a55_axu = float(a55_sp[6])
a60_axu = float(a60_sp[6])
a65_axu = float(a65_sp[6])
a70_axu = float(a70_sp[6])
a75_axu = float(a75_sp[6])
a80_axu = float(a80_sp[6])
a85_axu = float(a85_sp[6])
a90_axu = float(a90_sp[6])
a95_axu = float(a95_sp[6])
a100_axu = float(a100_sp[6])
a105_axu = float(a105_sp[6])
a110_axu = float(a110_sp[6])

# 2019 mx - 2019 LT
a0_m19 = float(a0_sp[7])
a1_m19 = float(a1_sp[7])
a5_m19 = float(a5_sp[7])
```

```
a10_m19 = float(a10_sp[7])
a15_m19 = float(a15_sp[7])
a20_m19 = float(a20_sp[7])
a25_m19 = float(a25_sp[7])
a30_m19 = float(a30_sp[7])
a35_m19 = float(a35_sp[7])
a40_m19 = float(a40_sp[7])
a45_m19 = float(a45_sp[7])
a50_m19 = float(a50_sp[7])
a55_m19 = float(a55_sp[7])
a60_m19 = float(a60_sp[7])
a65_m19 = float(a65_sp[7])
a70_m19 = float(a70_sp[7])
a75_m19 = float(a75_sp[7])
a80_m19 = float(a80_sp[7])
a85_m19 = float(a85_sp[7])
a90_m19 = float(a90_sp[7])
a95_m19 = float(a95_sp[7])
a100_m19 = float(a100_sp[7])
a105_m19 = float(a105_sp[7])
a110_m19 = float(a110_sp[7])


count = 0
while count < 50000: #5000: #50000

# Random qx by age - need to check we are not getting any values below zero

#    make this uniform
    a0_rand_qx = random.uniform(a0_qxl,a0_qxu)
    a1_rand_qx = random.uniform(a1_qxl,a1_qxu)
    a5_rand_qx = random.uniform(a5_qxl,a5_qxu)
    a10_rand_qx = random.uniform(a10_qxl,a10_qxu)
    a15_rand_qx = random.uniform(a15_qxl,a15_qxu)
    a20_rand_qx = random.uniform(a20_qxl,a20_qxu)
    a25_rand_qx = random.uniform(a25_qxl,a25_qxu)
    a30_rand_qx = random.uniform(a30_qxl,a30_qxu)
    a35_rand_qx = random.uniform(a35_qxl,a35_qxu)
    a40_rand_qx = random.uniform(a40_qxl,a40_qxu)
    a45_rand_qx = random.uniform(a45_qxl,a45_qxu)
    a50_rand_qx = random.uniform(a50_qxl,a50_qxu)
    a55_rand_qx = random.uniform(a55_qxl,a55_qxu)
    a60_rand_qx = random.uniform(a60_qxl,a60_qxu)
    a65_rand_qx = random.uniform(a65_qxl,a65_qxu)
    a70_rand_qx = random.uniform(a70_qxl,a70_qxu)
    a75_rand_qx = random.uniform(a75_qxl,a75_qxu)
    a80_rand_qx = random.uniform(a80_qxl,a80_qxu)
    a85_rand_qx = random.uniform(a85_qxl,a85_qxu)
    a90_rand_qx = random.uniform(a90_qxl,a90_qxu)
    a95_rand_qx = random.uniform(a95_qxl,a95_qxu)
    a100_rand_qx = random.uniform(a100_qxl,a100_qxu)
    a105_rand_qx = random.uniform(a105_qxl,a105_qxu)
    a110_rand_qx = 1

# Random ax by age - need to check we are not getting any values below zero

    #    make this uniform
    a0_rand_ax = random.uniform(a0_axl,a0_axu)
    a1_rand_ax = random.uniform(a1_axl,a1_axu)
    a5_rand_ax = random.uniform(a5_axl,a5_axu)
```

```
    a10_rand_ax = random.uniform(a10_axl,a10_axu)
    a15_rand_ax = random.uniform(a15_axl,a15_axu)
    a20_rand_ax = random.uniform(a20_axl,a20_axu)
    a25_rand_ax = random.uniform(a25_axl,a25_axu)
    a30_rand_ax = random.uniform(a30_axl,a30_axu)
    a35_rand_ax = random.uniform(a35_axl,a35_axu)
    a40_rand_ax = random.uniform(a40_axl,a40_axu)
    a45_rand_ax = random.uniform(a45_axl,a45_axu)
    a50_rand_ax = random.uniform(a50_axl,a50_axu)
    a55_rand_ax = random.uniform(a55_axl,a55_axu)
    a60_rand_ax = random.uniform(a60_axl,a60_axu)
    a65_rand_ax = random.uniform(a65_axl,a65_axu)
    a70_rand_ax = random.uniform(a70_axl,a70_axu)
    a75_rand_ax = random.uniform(a75_axl,a75_axu)
    a80_rand_ax = random.uniform(a80_axl,a80_axu)
    a85_rand_ax = random.uniform(a85_axl,a85_axu)
    a90_rand_ax = random.uniform(a90_axl,a90_axu)
    a95_rand_ax = random.uniform(a95_axl,a95_axu)
    a100_rand_ax = random.uniform(a100_axl,a100_axu)
    a105_rand_ax = random.uniform(a105_axl,a105_axu)
    a110_rand_ax = random.uniform(a105_axl,a105_axu)



### calculate life table variables
    radix = 1000000.0000000

    # calculate the number of deaths age0
    a0_dx = a0_rand_qx*radix
    # calculate survivors
    a0_lx=radix
    a0_sx=a0_lx/radix # this is 1?
    a1_lx=(radix-a0_dx)
    a1_sx = a1_lx/radix

    # calculate the number of deaths age1
    a1_dx = a1_rand_qx*a1_lx
    # calculate survivors
    a5_lx=(a1_lx-a1_dx)
    a5_sx = a5_lx/radix

    # calculate the number of deaths age5
    a5_dx = a5_rand_qx*a5_lx
    # calculate survivors
    a10_lx=(a5_lx-a5_dx)
    a10_sx = a10_lx/radix

    # calculate the number of deaths age10
    a10_dx = a10_rand_qx*a10_lx
    # calculate survivors
    a15_lx=(a10_lx-a10_dx)
    a15_sx = a15_lx/radix

    # calculate the number of deaths age15
    a15_dx = a15_rand_qx*a15_lx
    # calculate survivors
    a20_lx=(a15_lx-a15_dx)
    a20_sx = a20_lx/radix

    # calculate the number of deaths age20
```

```
a20_dx = a20_rand_qx*a20_lx
# calculate survivors
a25_lx=(a20_lx-a20_dx)
a25_sx = a25_lx/radix

# calculate the number of deaths age25
a25_dx = a25_rand_qx*a25_lx
# calculate survivors
a30_lx=(a25_lx-a25_dx)
a30_sx = a30_lx/radix

# calculate the number of deaths age30
a30_dx = a30_rand_qx*a30_lx
# calculate survivors
a35_lx=(a30_lx-a30_dx)
a35_sx = a35_lx/radix

# calculate the number of deaths age35
a35_dx = a35_rand_qx*a35_lx
# calculate survivors
a40_lx=(a35_lx-a35_dx)
a40_sx = a40_lx/radix

# calculate the number of deaths age40
a40_dx = a40_rand_qx*a40_lx
# calculate survivors
a45_lx=(a40_lx-a40_dx)
a45_sx = a45_lx/radix

# calculate the number of deaths age45
a45_dx = a45_rand_qx*a45_lx
# calculate survivors
a50_lx=(a45_lx-a45_dx)
a50_sx = a50_lx/radix

# calculate the number of deaths age50
a50_dx = a50_rand_qx*a50_lx
# calculate survivors
a55_lx=(a50_lx-a50_dx)
a55_sx = a55_lx/radix

# calculate the number of deaths age55
a55_dx = a55_rand_qx*a55_lx
# calculate survivors
a60_lx=(a55_lx-a55_dx)
a60_sx = a60_lx/radix

# calculate the number of deaths age60
a60_dx = a60_rand_qx*a60_lx
# calculate survivors
a65_lx=(a60_lx-a60_dx)
a65_sx = a65_lx/radix

# calculate the number of deaths age65
a65_dx = a65_rand_qx*a65_lx
# calculate survivors
a70_lx=(a65_lx-a65_dx)
a70_sx = a70_lx/radix

# calculate the number of deaths age70
```

```
a70_dx = a70_rand_qx*a70_lx
# calculate survivors
a75_lx=(a70_lx-a70_dx)
a75_sx = a75_lx/radix

# calculate the number of deaths age75
a75_dx = a75_rand_qx*a75_lx
# calculate survivors
a80_lx=(a75_lx-a75_dx)
a80_sx = a80_lx/radix

# calculate the number of deaths age80
a80_dx = a80_rand_qx*a80_lx
# calculate survivors
a85_lx=(a80_lx-a80_dx)
a85_sx = a85_lx/radix

# calculate the number of deaths age85
a85_dx = a85_rand_qx*a85_lx
# calculate survivors
a90_lx=(a85_lx-a85_dx)
a90_sx = a90_lx/radix

# calculate the number of deaths age90
a90_dx = a90_rand_qx*a90_lx
# calculate survivors
a95_lx=(a90_lx-a90_dx)
a95_sx = a95_lx/radix

# calculate the number of deaths age95
a95_dx = a95_rand_qx*a95_lx
# calculate survivors
a100_lx=(a95_lx-a95_dx)
a100_sx = a100_lx/radix

# calculate the number of deaths age100
a100_dx = a100_rand_qx*a100_lx
# calculate survivors
a105_lx=(a100_lx-a100_dx)
a105_sx = a105_lx/radix

# calculate the number of deaths age105
a105_dx = a105_rand_qx*a105_lx
# calculate survivors
a110_lx=(a105_lx-a105_dx)
a110_sx = a110_lx/radix

# calculate the number of deaths age110
a110_dx = a110_rand_qx*a110_lx
# No Survivors - top-coded

#calculate Lx
a0_Lx = (a1_lx*1)+(a0_dx*a0_rand_ax)
a1_Lx = (a5_lx*4)+(a1_dx*a1_rand_ax)
a5_Lx = (a10_lx*5)+(a5_dx*a5_rand_ax)
a10_Lx = (a15_lx*5)+(a10_dx*a10_rand_ax)
a15_Lx = (a20_lx*5)+(a15_dx*a15_rand_ax)
a20_Lx = (a25_lx*5)+(a20_dx*a20_rand_ax)
a25_Lx = (a30_lx*5)+(a25_dx*a25_rand_ax)
a30_Lx = (a35_lx*5)+(a30_dx*a30_rand_ax)
```

```
  a35_Lx = (a40_lx*5)+(a35_dx*a35_rand_ax)
  a40_Lx = (a45_lx*5)+(a40_dx*a40_rand_ax)
  a45_Lx = (a50_lx*5)+(a45_dx*a45_rand_ax)
  a50_Lx = (a55_lx*5)+(a50_dx*a50_rand_ax)
  a55_Lx = (a60_lx*5)+(a55_dx*a55_rand_ax)
  a60_Lx = (a65_lx*5)+(a60_dx*a60_rand_ax)
  a65_Lx = (a70_lx*5)+(a65_dx*a65_rand_ax)
  a70_Lx = (a75_lx*5)+(a70_dx*a70_rand_ax)
  a75_Lx = (a80_lx*5)+(a75_dx*a75_rand_ax)
  a80_Lx = (a85_lx*5)+(a80_dx*a80_rand_ax)
  a85_Lx = (a90_lx*5)+(a85_dx*a85_rand_ax)
  a90_Lx = (a95_lx*5)+(a90_dx*a90_rand_ax)
  a95_Lx = (a100_lx*5)+(a95_dx*a95_rand_ax)
  a100_Lx = (a105_lx*5)+(a100_dx*a100_rand_ax)
  a105_Lx = (a110_lx*5)+(a105_dx*a105_rand_ax)
  a110_Lx = (a110_dx*a110_rand_ax)

  ####
  # calculate Tx
  a0_Tx =
a0_Lx+a1_Lx+a5_Lx+a10_Lx+a15_Lx+a20_Lx+a25_Lx+a30_Lx+a35_Lx+a40_Lx+a45_Lx+a50_Lx+a55_Lx+a60_Lx+a65_Lx+a70_L
x+a75_Lx+a80_Lx+a85_Lx+a90_Lx+a95_Lx+a100_Lx+a105_Lx+a110_Lx
  a1_Tx =
a1_Lx+a5_Lx+a10_Lx+a15_Lx+a20_Lx+a25_Lx+a30_Lx+a35_Lx+a40_Lx+a45_Lx+a50_Lx+a55_Lx+a60_Lx+a65_Lx+a70_Lx+a75_
Lx+a80_Lx+a85_Lx+a90_Lx+a95_Lx+a100_Lx+a105_Lx+a110_Lx
  a5_Tx =
a5_Lx+a10_Lx+a15_Lx+a20_Lx+a25_Lx+a30_Lx+a35_Lx+a40_Lx+a45_Lx+a50_Lx+a55_Lx+a60_Lx+a65_Lx+a70_Lx+a75_Lx+a80
_Lx+a85_Lx+a90_Lx+a95_Lx+a100_Lx+a105_Lx+a110_Lx
  a10_Tx =
a10_Lx+a15_Lx+a20_Lx+a25_Lx+a30_Lx+a35_Lx+a40_Lx+a45_Lx+a50_Lx+a55_Lx+a60_Lx+a65_Lx+a70_Lx+a75_Lx+a80_Lx+a8
5_Lx+a90_Lx+a95_Lx+a100_Lx+a105_Lx+a110_Lx
  a15_Tx =
a15_Lx+a20_Lx+a25_Lx+a30_Lx+a35_Lx+a40_Lx+a45_Lx+a50_Lx+a55_Lx+a60_Lx+a65_Lx+a70_Lx+a75_Lx+a80_Lx+a85_Lx+a9
0_Lx+a95_Lx+a100_Lx+a105_Lx+a110_Lx
  a20_Tx =
a20_Lx+a25_Lx+a30_Lx+a35_Lx+a40_Lx+a45_Lx+a50_Lx+a55_Lx+a60_Lx+a65_Lx+a70_Lx+a75_Lx+a80_Lx+a85_Lx+a90_Lx+a9
5_Lx+a100_Lx+a105_Lx+a110_Lx
  a25_Tx =
a25_Lx+a30_Lx+a35_Lx+a40_Lx+a45_Lx+a50_Lx+a55_Lx+a60_Lx+a65_Lx+a70_Lx+a75_Lx+a80_Lx+a85_Lx+a90_Lx+a95_Lx+a1
00_Lx+a105_Lx+a110_Lx
  a30_Tx =
a30_Lx+a35_Lx+a40_Lx+a45_Lx+a50_Lx+a55_Lx+a60_Lx+a65_Lx+a70_Lx+a75_Lx+a80_Lx+a85_Lx+a90_Lx+a95_Lx+a100_Lx+a
105_Lx+a110_Lx
  a35_Tx =
a35_Lx+a40_Lx+a45_Lx+a50_Lx+a55_Lx+a60_Lx+a65_Lx+a70_Lx+a75_Lx+a80_Lx+a85_Lx+a90_Lx+a95_Lx+a100_Lx+a105_Lx+
a110_Lx
  a40_Tx =
a40_Lx+a45_Lx+a50_Lx+a55_Lx+a60_Lx+a65_Lx+a70_Lx+a75_Lx+a80_Lx+a85_Lx+a90_Lx+a95_Lx+a100_Lx+a105_Lx+a110_Lx
  a45_Tx =
a45_Lx+a50_Lx+a55_Lx+a60_Lx+a65_Lx+a70_Lx+a75_Lx+a80_Lx+a85_Lx+a90_Lx+a95_Lx+a100_Lx+a105_Lx+a110_Lx
  a50_Tx = a50_Lx+a55_Lx+a60_Lx+a65_Lx+a70_Lx+a75_Lx+a80_Lx+a85_Lx+a90_Lx+a95_Lx+a100_Lx+a105_Lx+a110_Lx
  a55_Tx = a55_Lx+a60_Lx+a65_Lx+a70_Lx+a75_Lx+a80_Lx+a85_Lx+a90_Lx+a95_Lx+a100_Lx+a105_Lx+a110_Lx
  a60_Tx = a60_Lx+a65_Lx+a70_Lx+a75_Lx+a80_Lx+a85_Lx+a90_Lx+a95_Lx+a100_Lx+a105_Lx+a110_Lx
  a65_Tx = a65_Lx+a70_Lx+a75_Lx+a80_Lx+a85_Lx+a90_Lx+a95_Lx+a100_Lx+a105_Lx+a110_Lx
  a70_Tx = a70_Lx+a75_Lx+a80_Lx+a85_Lx+a90_Lx+a95_Lx+a100_Lx+a105_Lx+a110_Lx
  a75_Tx = a75_Lx+a80_Lx+a85_Lx+a90_Lx+a95_Lx+a100_Lx+a105_Lx+a110_Lx
  a80_Tx = a80_Lx+a85_Lx+a90_Lx+a95_Lx+a100_Lx+a105_Lx+a110_Lx
  a85_Tx = a85_Lx+a90_Lx+a95_Lx+a100_Lx+a105_Lx+a110_Lx
  a90_Tx = a90_Lx+a95_Lx+a100_Lx+a105_Lx+a110_Lx
  a95_Tx = a95_Lx+a100_Lx+a105_Lx+a110_Lx
```

```
    a100_Tx = a100_Lx+a105_Lx+a110_Lx
    a105_Tx = a105_Lx+a110_Lx
    a110_Tx = a110_Lx

#calculate mx
    a0_mx = (a0_dx)/a0_Lx
    a1_mx = (a1_dx)/a1_Lx
    a5_mx = (a5_dx)/a5_Lx
    a10_mx = (a10_dx)/a10_Lx
    a15_mx = (a15_dx)/a15_Lx
    a20_mx = (a20_dx)/a20_Lx
    a25_mx = (a25_dx)/a25_Lx
    a30_mx = (a30_dx)/a30_Lx
    a35_mx = (a35_dx)/a35_Lx
    a40_mx = (a40_dx)/a40_Lx
    a45_mx = (a45_dx)/a45_Lx
    a50_mx = (a50_dx)/a50_Lx
    a55_mx = (a55_dx)/a55_Lx
    a60_mx = (a60_dx)/a60_Lx
    a65_mx = (a65_dx)/a65_Lx
    a70_mx = (a70_dx)/a70_Lx
    a75_mx = (a75_dx)/a75_Lx
    a80_mx = (a80_dx)/a80_Lx
    a85_mx = (a85_dx)/a85_Lx
    a90_mx = (a90_dx)/a90_Lx
    a95_mx = (a95_dx)/a95_Lx
    a100_mx = (a100_dx)/a100_Lx
    a105_mx = (a105_dx)/a105_Lx
    a110_mx = (a110_dx)/a110_Lx


    #calculate Lx 0-14, 15-39, 40-64, 65-84, 85+

    a015_tx = (a0_Lx+a1_Lx+a5_Lx+a10_Lx)
    a1539_tx = (a15_Lx+a20_Lx+a25_Lx+a30_Lx+a35_Lx)
    a4064_tx = (a40_Lx+a45_Lx+a50_Lx+a55_Lx+a60_Lx)
    a6584_tx = (a65_Lx+a70_Lx+a75_Lx+a80_Lx)
    a85_tx = a85_Tx

# simulation of weighted-averages of 2020:2019 mx ratio:0-15, 15-40, 40-65, 65-85, 85+

    a015_rr  =
(a0_mx/a0_m19)*(a0_Lx/a015_tx)+(a1_mx/a1_m19)*(a1_Lx/a015_tx)+(a5_mx/a5_m19)*(a5_Lx/a015_tx)+(a10_mx/a10_m19)*(a10_
Lx/a015_tx)
    a1539_rr =
(a15_mx/a15_m19)*(a15_Lx/a1539_tx)+(a20_mx/a20_m19)*(a20_Lx/a1539_tx)+(a25_mx/a25_m19)*(a25_Lx/a1539_tx)+(a30_mx/a
30_m19)*(a30_Lx/a1539_tx)+(a35_mx/a35_m19)*(a35_Lx/a1539_tx)
    a4064_rr =
(a40_mx/a40_m19)*(a40_Lx/a4064_tx)+(a45_mx/a45_m19)*(a45_Lx/a4064_tx)+(a50_mx/a50_m19)*(a50_Lx/a4064_tx)+(a55_mx/a
55_m19)*(a55_Lx/a4064_tx)+(a60_mx/a60_m19)*(a60_Lx/a4064_tx)
    a6584_rr =
(a65_mx/a65_m19)*(a65_Lx/a6584_tx)+(a70_mx/a70_m19)*(a70_Lx/a6584_tx)+(a75_mx/a75_m19)*(a75_Lx/a6584_tx)+(a80_mx/a
80_m19)*(a80_Lx/a6584_tx)
    a85110_rr =
(a85_mx/a85_m19)*(a85_Lx/a85_tx)+(a90_mx/a90_m19)*(a90_Lx/a85_tx)+(a95_mx/a95_m19)*(a95_Lx/a85_tx)+(a100_mx/a100_m
19)*(a100_Lx/a85_tx)+(a105_mx/a105_m19)*(a105_Lx/a85_tx)+(a110_mx/a110_m19)*(a110_Lx/a85_tx)


# this outputs the mortality rates for each sim
    mxgrp = r"/Users/ryan/Documents/Papers/LE Trends_COVID/2021 Life Expectancy/simulations/peer/total/esp_t_mxgrp.txt"
```

```
opened_file = open(mxgrp, 'a')

if count==0:
    opened_file.write('{0} {1} {2} {3} {4} {5}\n'.format("sim_num","rr015","rr1539","rr4064","rr6584","rr85"))

else:
    opened_file.write('{0} {1} {2} {3} {4} {5}\n'.format(count,a015_rr,a1539_rr,a4064_rr,a6584_rr,a85110_rr))


###### estimate life expectancy

#calculate ex
a0_ex = a0_Tx/radix
a1_ex = a1_Tx/a1_lx
a5_ex = a5_Tx/a5_lx
a10_ex = a10_Tx/a10_lx
a15_ex = a15_Tx/a15_lx
a20_ex = a20_Tx/a20_lx
a25_ex = a25_Tx/a25_lx
a30_ex = a30_Tx/a30_lx
a35_ex = a35_Tx/a35_lx
a40_ex = a40_Tx/a40_lx
a45_ex = a45_Tx/a45_lx
a50_ex = a50_Tx/a50_lx
a55_ex = a55_Tx/a55_lx
a60_ex = a60_Tx/a60_lx
a65_ex = a65_Tx/a65_lx
a70_ex = a70_Tx/a70_lx
a75_ex = a75_Tx/a75_lx
a80_ex = a80_Tx/a80_lx
a85_ex = a85_Tx/a85_lx
a90_ex = a90_Tx/a90_lx
a95_ex = a95_Tx/a95_lx
a100_ex = a100_Tx/a100_lx
a105_ex = a105_Tx/a105_lx
a110_ex = a110_Tx/a110_lx


# save data

tot_file_name = r"/Users/ryan/Documents/Papers/LE Trends_COVID/2021 Life Expectancy/simulations/peer/total/esp_t_ex.txt"

tot_opened_file = open(tot_file_name, 'a')

if count==0:
    tot_opened_file.write('{0} {1} {2} {3}\n'.format("sim_num","age","sx", "ex"))
    tot_opened_file.write('{0} {1} {2} {3}\n'.format(count,"0",a0_sx,a0_ex))
    tot_opened_file.write('{0} {1} {2} {3}\n'.format(count,"1",a1_sx,a1_ex))
    tot_opened_file.write('{0} {1} {2} {3}\n'.format(count,"5",a5_sx,a5_ex))
    tot_opened_file.write('{0} {1} {2} {3}\n'.format(count,"10",a10_sx,a10_ex))
    tot_opened_file.write('{0} {1} {2} {3}\n'.format(count,"15",a15_sx,a15_ex))
    tot_opened_file.write('{0} {1} {2} {3}\n'.format(count,"20",a20_sx,a20_ex))
    tot_opened_file.write('{0} {1} {2} {3}\n'.format(count,"25",a25_sx,a25_ex))
    tot_opened_file.write('{0} {1} {2} {3}\n'.format(count,"30",a30_sx,a30_ex))
    tot_opened_file.write('{0} {1} {2} {3}\n'.format(count,"35",a35_sx,a35_ex))
    tot_opened_file.write('{0} {1} {2} {3}\n'.format(count,"40",a40_sx,a40_ex))
    tot_opened_file.write('{0} {1} {2} {3}\n'.format(count,"45",a45_sx,a45_ex))
    tot_opened_file.write('{0} {1} {2} {3}\n'.format(count,"50",a50_sx,a50_ex))
    tot_opened_file.write('{0} {1} {2} {3}\n'.format(count,"55",a55_sx,a55_ex))
    tot_opened_file.write('{0} {1} {2} {3}\n'.format(count,"60",a60_sx,a60_ex))
```

```
        tot_opened_file.write('{0} {1} {2} {3}\n'.format(count,"65",a65_sx,a65_ex))
        tot_opened_file.write('{0} {1} {2} {3}\n'.format(count,"70",a70_sx,a70_ex))
        tot_opened_file.write('{0} {1} {2} {3}\n'.format(count,"75",a75_sx,a75_ex))
        tot_opened_file.write('{0} {1} {2} {3}\n'.format(count,"80",a80_sx,a80_ex))
        tot_opened_file.write('{0} {1} {2} {3}\n'.format(count,"85",a85_sx,a85_ex))
        tot_opened_file.write('{0} {1} {2} {3}\n'.format(count,"90",a90_sx,a90_ex))
        tot_opened_file.write('{0} {1} {2} {3}\n'.format(count,"95",a95_sx,a95_ex))
        tot_opened_file.write('{0} {1} {2} {3}\n'.format(count,"100",a100_sx,a100_ex))
        tot_opened_file.write('{0} {1} {2} {3}\n'.format(count,"105",a105_sx,a105_ex))
        tot_opened_file.write('{0} {1} {2} {3}\n'.format(count,"110",a110_sx,a110_ex))


    else:
        tot_opened_file.write('{0} {1} {2} {3}\n'.format(count,"0",a0_sx,a0_ex))
        tot_opened_file.write('{0} {1} {2} {3}\n'.format(count,"1",a1_sx,a1_ex))
        tot_opened_file.write('{0} {1} {2} {3}\n'.format(count,"5",a5_sx,a5_ex))
        tot_opened_file.write('{0} {1} {2} {3}\n'.format(count,"10",a10_sx,a10_ex))
        tot_opened_file.write('{0} {1} {2} {3}\n'.format(count,"15",a15_sx,a15_ex))
        tot_opened_file.write('{0} {1} {2} {3}\n'.format(count,"20",a20_sx,a20_ex))
        tot_opened_file.write('{0} {1} {2} {3}\n'.format(count,"25",a25_sx,a25_ex))
        tot_opened_file.write('{0} {1} {2} {3}\n'.format(count,"30",a30_sx,a30_ex))
        tot_opened_file.write('{0} {1} {2} {3}\n'.format(count,"35",a35_sx,a35_ex))
        tot_opened_file.write('{0} {1} {2} {3}\n'.format(count,"40",a40_sx,a40_ex))
        tot_opened_file.write('{0} {1} {2} {3}\n'.format(count,"45",a45_sx,a45_ex))
        tot_opened_file.write('{0} {1} {2} {3}\n'.format(count,"50",a50_sx,a50_ex))
        tot_opened_file.write('{0} {1} {2} {3}\n'.format(count,"55",a55_sx,a55_ex))
        tot_opened_file.write('{0} {1} {2} {3}\n'.format(count,"60",a60_sx,a60_ex))
        tot_opened_file.write('{0} {1} {2} {3}\n'.format(count,"65",a65_sx,a65_ex))
        tot_opened_file.write('{0} {1} {2} {3}\n'.format(count,"70",a70_sx,a70_ex))
        tot_opened_file.write('{0} {1} {2} {3}\n'.format(count,"75",a75_sx,a75_ex))
        tot_opened_file.write('{0} {1} {2} {3}\n'.format(count,"80",a80_sx,a80_ex))
        tot_opened_file.write('{0} {1} {2} {3}\n'.format(count,"85",a85_sx,a85_ex))
        tot_opened_file.write('{0} {1} {2} {3}\n'.format(count,"90",a90_sx,a90_ex))
        tot_opened_file.write('{0} {1} {2} {3}\n'.format(count,"95",a95_sx,a95_ex))
        tot_opened_file.write('{0} {1} {2} {3}\n'.format(count,"100",a100_sx,a100_ex))
        tot_opened_file.write('{0} {1} {2} {3}\n'.format(count,"105",a105_sx,a105_ex))
        tot_opened_file.write('{0} {1} {2} {3}\n'.format(count,"110",a110_sx,a110_ex))

    print(count)
    count += 1  # This is the same as count = count + 1

tot_opened_file.close()

print("simulation completed")
```

**Estimate Median and Credible Range from 2021 Life Expectancy Distributions, Peer Total**

```
*****************************************************************
********************* Append all Countries *********************
***** Distributions of Sim LE at birth for 21 peer countries *****
*****************************************************************

* 21 Country Peer Comparison Group, 5/23/22 HMDB-STMF data
* Input Simulation Results into Stata

* Australia, Total Pop

import delimited "/Users/ryan/Documents/Papers/LE Trends_COVID/2021 Life
Expectancy/simulations/peer/total/aus_t_ex.txt", delimiter(space) varnames(1)
encoding(ISO-8859-1)clear

gen country="Australia"
keep if age == 0

save "/Users/ryan/Documents/Papers/LE Trends_COVID/2021 Life
Expectancy/simulations/peer/total/aus_t.dta", replace


* Austria, Total Pop

import delimited "/Users/ryan/Documents/Papers/LE Trends_COVID/2021 Life
Expectancy/simulations/peer/total/aut_t_ex.txt", delimiter(space) varnames(1)
encoding(ISO-8859-1)clear

gen country="Austria"
keep if age == 0

save "/Users/ryan/Documents/Papers/LE Trends_COVID/2021 Life
Expectancy/simulations/peer/total/aut_t.dta", replace


* Belgium, Total Pop

import delimited "/Users/ryan/Documents/Papers/LE Trends_COVID/2021 Life
Expectancy/simulations/peer/total/bel_t_ex.txt", delimiter(space) encoding(ISO-8859-1)
clear

gen country="Belgium"
keep if age == 0

save "/Users/ryan/Documents/Papers/LE Trends_COVID/2021 Life
Expectancy/simulations/peer/total/bel_t.dta", replace


* Canada, Total Pop

import delimited "/Users/ryan/Documents/Papers/LE Trends_COVID/2021 Life
Expectancy/simulations/peer/total/can_t_ex.txt", delimiter(space) encoding(ISO-8859-1)
clear

gen country="Canada"
keep if age == 0

save "/Users/ryan/Documents/Papers/LE Trends_COVID/2021 Life
Expectancy/simulations/peer/total/can_t.dta", replace
```

```
* Denmark, Total Pop

import delimited "/Users/ryan/Documents/Papers/LE Trends_COVID/2021 Life
Expectancy/simulations/peer/total/dnk_t_ex.txt", delimiter(space) encoding(ISO-8859-1)
clear

gen country="Denmark"
keep if age == 0

save "/Users/ryan/Documents/Papers/LE Trends_COVID/2021 Life
Expectancy/simulations/peer/total/dnk_t.dta", replace


* Finland, Total Pop

import delimited "/Users/ryan/Documents/Papers/LE Trends_COVID/2021 Life
Expectancy/simulations/peer/total/fin_t_ex.txt", delimiter(space) encoding(ISO-8859-1)
clear

gen country="Finland"
keep if age == 0

save "/Users/ryan/Documents/Papers/LE Trends_COVID/2021 Life
Expectancy/simulations/peer/total/fin_t.dta", replace


* Germany, Total Pop

import delimited "/Users/ryan/Documents/Papers/LE Trends_COVID/2021 Life
Expectancy/simulations/peer/total/deu_t_ex.txt", delimiter(space) encoding(ISO-8859-1)
clear

gen country="Germany"
keep if age == 0

save "/Users/ryan/Documents/Papers/LE Trends_COVID/2021 Life
Expectancy/simulations/peer/total/deu_t.dta", replace


* England, Total Pop

import delimited "/Users/ryan/Documents/Papers/LE Trends_COVID/2021 Life
Expectancy/simulations/peer/total/eng_t_ex.txt", delimiter(space) encoding(ISO-8859-1)
clear

gen country="England & Wales"
keep if age == 0

save "/Users/ryan/Documents/Papers/LE Trends_COVID/2021 Life
Expectancy/simulations/peer/total/eng_t.dta", replace


* Spain, Total Pop

import delimited "/Users/ryan/Documents/Papers/LE Trends_COVID/2021 Life
Expectancy/simulations/peer/total/esp_t_ex.txt", delimiter(space) encoding(ISO-8859-1)
clear

gen country="Spain"
keep if age == 0
```

```
save "/Users/ryan/Documents/Papers/LE Trends_COVID/2021 Life
Expectancy/simulations/peer/total/esp_t.dta", replace

* France, Total Pop

import delimited "/Users/ryan/Documents/Papers/LE Trends_COVID/2021 Life
Expectancy/simulations/peer/total/fra_t_ex.txt", delimiter(space) encoding(ISO-8859-1)
clear

gen country="France"
keep if age == 0

save "/Users/ryan/Documents/Papers/LE Trends_COVID/2021 Life
Expectancy/simulations/peer/total/fra_t.dta", replace


* Israel, Total Pop

import delimited "/Users/ryan/Documents/Papers/LE Trends_COVID/2021 Life
Expectancy/simulations/peer/total/isr_t_ex.txt", delimiter(space) encoding(ISO-8859-1)
clear

gen country="Israel"
keep if age == 0

save "/Users/ryan/Documents/Papers/LE Trends_COVID/2021 Life
Expectancy/simulations/peer/total/isr_t.dta", replace


* Italy, Total Pop

import delimited "/Users/ryan/Documents/Papers/LE Trends_COVID/2021 Life
Expectancy/simulations/peer/total/ita_t_ex.txt", delimiter(space) encoding(ISO-8859-1)
clear

gen country="Italy"
keep if age == 0

save "/Users/ryan/Documents/Papers/LE Trends_COVID/2021 Life
Expectancy/simulations/peer/total/ita_t.dta", replace


* Northern Ireland, Total Pop

import delimited "/Users/ryan/Documents/Papers/LE Trends_COVID/2021 Life
Expectancy/simulations/peer/total/nir_t_ex.txt", delimiter(space) encoding(ISO-8859-1)
clear

gen country="Northern Ireland"
keep if age == 0

save "/Users/ryan/Documents/Papers/LE Trends_COVID/2021 Life
Expectancy/simulations/peer/total/nir_t.dta", replace


* Netherlands, Total Pop

import delimited "/Users/ryan/Documents/Papers/LE Trends_COVID/2021 Life
Expectancy/simulations/peer/total/nld_t_ex.txt", delimiter(space) encoding(ISO-8859-1)
clear

gen country="Netherlands"
```

```
keep if age == 0

save "/Users/ryan/Documents/Papers/LE Trends_COVID/2021 Life
Expectancy/simulations/peer/total/nld_t.dta", replace


* Norway, Total Pop

import delimited "/Users/ryan/Documents/Papers/LE Trends_COVID/2021 Life
Expectancy/simulations/peer/total/nor_t_ex.txt", delimiter(space) encoding(ISO-8859-1)
clear

gen country="Norway"
keep if age == 0

save "/Users/ryan/Documents/Papers/LE Trends_COVID/2021 Life
Expectancy/simulations/peer/total/nor_t.dta", replace


* Portugal, Total Pop

import delimited "/Users/ryan/Documents/Papers/LE Trends_COVID/2021 Life
Expectancy/simulations/peer/total/por_t_ex.txt", delimiter(space) encoding(ISO-8859-1)
clear

gen country="Portugal"
keep if age == 0

save "/Users/ryan/Documents/Papers/LE Trends_COVID/2021 Life
Expectancy/simulations/peer/total/por_t.dta", replace


* Scotland, Total Pop

import delimited "/Users/ryan/Documents/Papers/LE Trends_COVID/2021 Life
Expectancy/simulations/peer/total/sco_t_ex.txt", delimiter(space) encoding(ISO-8859-1)
clear

gen country="Scotland"
keep if age == 0

save "/Users/ryan/Documents/Papers/LE Trends_COVID/2021 Life
Expectancy/simulations/peer/total/sco_t.dta", replace


* Sweden, Total Pop

import delimited "/Users/ryan/Documents/Papers/LE Trends_COVID/2021 Life
Expectancy/simulations/peer/total/swe_t_ex.txt", delimiter(space) encoding(ISO-8859-1)
clear

gen country="Sweden"
keep if age == 0

save "/Users/ryan/Documents/Papers/LE Trends_COVID/2021 Life
Expectancy/simulations/peer/total/swe_t.dta", replace


* Switzerland, Total Pop

import delimited "/Users/ryan/Documents/Papers/LE Trends_COVID/2021 Life
Expectancy/simulations/peer/total/che_t_ex.txt", delimiter(space) encoding(ISO-8859-1)
clear
```

```
gen country="Switzerland"
keep if age == 0

save "/Users/ryan/Documents/Papers/LE Trends_COVID/2021 Life
Expectancy/simulations/peer/total/che_t.dta", replace


* New Zealand, Total Pop

import delimited "/Users/ryan/Documents/Papers/LE Trends_COVID/2021 Life
Expectancy/simulations/peer/total/nzl_t_ex.txt", delimiter(space) encoding(ISO-8859-1)
clear

gen country="New Zealand"
keep if age == 0

save "/Users/ryan/Documents/Papers/LE Trends_COVID/2021 Life
Expectancy/simulations/peer/total/nzl_t.dta", replace


* South Korea, Total Pop

import delimited "/Users/ryan/Documents/Papers/LE Trends_COVID/2021 Life
Expectancy/simulations/peer/total/kor_t_ex.txt", delimiter(space) encoding(ISO-8859-1)
clear

gen country="South Korea"
keep if age == 0

save "/Users/ryan/Documents/Papers/LE Trends_COVID/2021 Life
Expectancy/simulations/peer/total/kor_t.dta", replace


*****************************************************************
********************** Append all Countries *********************
***** Distributions of Sim LE at birth for 21 peer countries *****
*****************************************************************

use "/Users/ryan/Documents/Papers/LE Trends_COVID/2021 Life
Expectancy/simulations/peer/total/aut_t.dta", clear

append using "/Users/ryan/Documents/Papers/LE Trends_COVID/2021 Life
Expectancy/simulations/peer/total/bel_t.dta"

append using "/Users/ryan/Documents/Papers/LE Trends_COVID/2021 Life
Expectancy/simulations/peer/total/dnk_t.dta"

append using "/Users/ryan/Documents/Papers/LE Trends_COVID/2021 Life
Expectancy/simulations/peer/total/fin_t.dta"

append using "/Users/ryan/Documents/Papers/LE Trends_COVID/2021 Life
Expectancy/simulations/peer/total/deu_t.dta"

append using "/Users/ryan/Documents/Papers/LE Trends_COVID/2021 Life
Expectancy/simulations/peer/total/eng_t.dta"

append using "/Users/ryan/Documents/Papers/LE Trends_COVID/2021 Life
Expectancy/simulations/peer/total/esp_t.dta"

append using "/Users/ryan/Documents/Papers/LE Trends_COVID/2021 Life
Expectancy/simulations/peer/total/fra_t.dta"
```

```
append using "/Users/ryan/Documents/Papers/LE Trends_COVID/2021 Life
Expectancy/simulations/peer/total/isr_t.dta"

append using "/Users/ryan/Documents/Papers/LE Trends_COVID/2021 Life
Expectancy/simulations/peer/total/nir_t.dta"

append using "/Users/ryan/Documents/Papers/LE Trends_COVID/2021 Life
Expectancy/simulations/peer/total/nld_t.dta"

append using "/Users/ryan/Documents/Papers/LE Trends_COVID/2021 Life
Expectancy/simulations/peer/total/nor_t.dta"

append using "/Users/ryan/Documents/Papers/LE Trends_COVID/2021 Life
Expectancy/simulations/peer/total/por_t.dta"

append using "/Users/ryan/Documents/Papers/LE Trends_COVID/2021 Life
Expectancy/simulations/peer/total/sco_t.dta"

append using "/Users/ryan/Documents/Papers/LE Trends_COVID/2021 Life
Expectancy/simulations/peer/total/swe_t.dta"

append using "/Users/ryan/Documents/Papers/LE Trends_COVID/2021 Life
Expectancy/simulations/peer/total/che_t.dta"

append using "/Users/ryan/Documents/Papers/LE Trends_COVID/2021 Life
Expectancy/simulations/peer/total/nzl_t.dta"

append using "/Users/ryan/Documents/Papers/LE Trends_COVID/2021 Life
Expectancy/simulations/peer/total/kor_t.dta"

append using "/Users/ryan/Documents/Papers/LE Trends_COVID/2021 Life
Expectancy/simulations/peer/total/ita_t.dta"

append using "/Users/ryan/Documents/Papers/LE Trends_COVID/2021 Life
Expectancy/simulations/peer/total/can_t.dta"

append using "/Users/ryan/Documents/Papers/LE Trends_COVID/2021 Life
Expectancy/simulations/peer/total/aus_t.dta"


save "/Users/ryan/Documents/Papers/LE Trends_COVID/2021 Life
Expectancy/simulations/peer/total_2021ex_sim.dta", replace

tabstat ex if age == 0, statistics( p50 p5 p95 ) by(country)
```
